# Exposure contrasts of pregnant women during the Household Air Pollution Intervention Network randomized controlled trial

**DOI:** 10.1101/2021.11.04.21265938

**Authors:** Michael Johnson, Ajay Pillarisetti, Ricardo Piedrahita, Kalpana Balakrishnan, Jennifer L. Peel, Kyle Steenland, Lindsay J. Underhill, Ghislaine Rosa, Miles A. Kirby, Anaité Díaz-Artiga, John McCracken, Maggie L. Clark, Lance Waller, Howard Chang, Jiantong Wang, Ephrem Dusabimana, Florien Ndagijimana, Sankar Sambandam, Krishnendu Mukhopadhyay, Katherine Kearns, Devan Campbell, Jacob Kremer, Joshua Rosenthal, William Checkley, Thomas Clasen, Luke Naeher, on behalf of the Household Air Pollution Intervention Network (HAPIN) trial Investigators

## Abstract

**Background:** Exposure to PM_2.5_ arising from solid fuel combustion is estimated to result in approximately 2.3 million premature deaths and 90 million lost disability-adjusted life years annually. ‘Clean’ cooking interventions attempting to mitigate this burden have had limited success in reducing exposures to levels that may yield improved health outcomes.

**Objectives:** This paper reports exposure reductions achieved by a liquified petroleum gas (LPG) stove and fuel intervention for pregnant mothers in the Household Air Pollution Intervention Network (HAPIN) randomized controlled trial.

**Methods:** The HAPIN trial included 3,195 households primarily using biomass for cooking in Guatemala, India, Peru, and Rwanda. 24-hour exposures to PM_2.5_, carbon monoxide (CO), and black carbon (BC) were measured for pregnant women once before randomization into control (n=1605) and LPG arms (n=1590) and twice thereafter (aligned with trimester). Changes in exposure were estimated by directly comparing exposures between intervention and control arms and by using linear mixed-effect models to estimate the impact of the intervention on exposure levels.

**Results:** Median exposures of PM_2.5_, BC, and CO post-randomization in the intervention arm were lower by 66% (70.7 versus 24.0 μg/m^3^), 71% (9.6 versus 2.8 μg/m^3^), and 83% (1.2 versus ppm), respectively, compared to the control arm. Exposure reductions were similar across research locations. Post-intervention PM_2.5_ exposures in the intervention arm were at the lower end of what has been reported for LPG and other clean fuel interventions, with 69% of PM_2.5_ samples falling below the WHO Annual Interim Target 1 of 35 μg/m^3^.

**Discussion:** This study indicates that an LPG intervention with high displacement of traditional cooking can reduce exposures to levels thought to be associated with health benefits. Success in reducing exposures was likely due to strong performance of, and high adherence to the intervention.

## 1. Introduction

Household air pollution (HAP) from the incomplete combustion of solid fuels—including wood, dung, and crop residues—results in exposure to particulate matter with an aerodynamic diameter of ≤ 2.5 μm (PM2.5), carbon monoxide (CO), black carbon (BC), and other emissions that are hazardous to human health. Exposure to PM2.5 arising from solid fuel combustion is estimated to result in approximately 2.3 million premature deaths and 90 million lost disability-adjusted life years annually (Institute for Health Metrics and Evaluation). Approximately half of the world’s population relies on these fuels for cooking (Health Effects Institute 2020), predominantly in low- and middle-income countries.

Many studies have documented associations between HAP exposure and health endpoints, including cardiopulmonary outcomes, cancer, and pneumonia.(Lee et al. 2020) Few are randomized controlled trials of cleaner cooking interventions, such as LPG or ethanol (Alexander et al. 2017; Checkley et al. 2020; Chillrud et al. 2021; Katz et al. 2020; Kirby et al. 2019; Mortimer et al. 2020; Romieu et al. 2009; Smith et al. 2011). Fewer yet have coupled a randomized study design with the rigorous measurement needed to demonstrate that an intervention results in reduced exposures that may in turn yield health benefits. Many previously evaluated clean cooking interventions either (1) were not assessed rigorously enough to evaluate impacts on changes in exposure (Clark et al. 2013; Hanna et al. 2016) or (2) insufficiently reduced exposures [e.g., were used alongside traditional stoves, resulting in attenuated exposure reductions or were conducted in locations with elevated ambient air pollution concentrations (Mortimer et al. 2020)]. Better characterization of exposure contrasts achieved by clean fuel interventions is important for understanding their health implications.

As part of the Household Air Pollution Intervention Network (HAPIN) study, we undertook extensive personal air pollution exposure assessment at baseline (prior to randomization) and at multiple timepoints during pregnancy and after. We report the impact of HAPIN’s LPG intervention on personal PM_2.5_, CO, and BC exposures among pregnant participants.

## 2. Methods

### a. Study setting

The HAPIN study is a randomized controlled liquefied petroleum gas (LPG) fuel and stove intervention trial underway in four international research centers (IRC) in Guatemala, India, Peru, and Rwanda. Biomass use is common as a primary cooking fuel in each of the IRCs. Using a common protocol across each site, the trial aims to assess the health effects among pregnant women (n = 3200), children (n = 3200) and non-pregnant adult women living in the same household (n = 444). Participants were randomly assigned to receive LPG stoves, continuous fuel delivery, and regular behavioral messaging versus continued use of a biomass-burning stove. Primary health outcomes are birth weight, incidence of severe pediatric pneumonia, stunted growth in the children, and blood pressure in the non-pregnant adult women.

The study protocol has been reviewed and approved by institutional review boards (IRBs) or Ethics Committees at Emory University (00089799), Johns Hopkins University (00007403), Sri Ramachandra Institute of Higher Education and Research (IEC-N1/16/JUL/54/49) and the Indian Council of Medical Research – Health Ministry Screening Committee (5/8/4-30/(Env)/Indo-US/2016-NCD-I), Universidad del Valle de Guatemala (146-08-2016) and Guatemalan Ministry of Health National Ethics Committee (11-2016), Asociación Benefica PRISMA (CE2981.17), the London School of Hygiene and Tropical Medicine (11664-5) and the Rwandan National Ethics Committee (No.357/RNEC/2018), and Washington University in St. Louis (201611159). The study has been registered with ClinicalTrials.gov (Identifier NCT02944682).

### b. Site and participants information

Personal exposures to PM_2.5_, BC, and CO reported in this manuscript were based on measurements from pregnant participants over three visits at each of the four HAPIN IRCs, with the approach and site descriptions described in detail previously (Clasen et al. 2020; Johnson et al. 2020). Briefly, sites (in Guatemala, India, Peru, and Rwanda) are characterized as rural, with low background air pollution concentrations. Households typically use traditional biomass stoves to fulfill their cooking energy needs.

Inclusion criteria were as follows: women aged 18-35 years who were pregnant between 9 and 20 weeks gestation with a viable singleton pregnancy, who used primarily biomass fuel for cooking, and who agreed to participate via informed consent. Exclusion criteria included tobacco use, plans to move outside the study area, and plans to switch to clean fuels.

Additional information describing the study sites, participants, and recruitment can be found in the supporting information (SI Tables S1 and S2) and in Clasen et al. 2020.

### c. Air pollutant sampling instrumentation

We used the RTI Enhanced Children’s MicroPEM™ (ECM, RTI International, Research Triangle Park, USA) to measure exposure to PM_2.5_ (Chartier 2017). The ECM utilizes a 2.5 micron size-cut impactor at a flow rate of 0.3 liters per minute and measures continuous PM_2.5_ concentrations using a nephelometer. It simultaneously collects integrated gravimetric samples on 15mm polytetrafluoroethylene filters (Measurement Technology Laboratories, USA). The ECM is lightweight (approximately 150 g), small (2.5 × 6.5 × 12.5cm), and nearly silent during use. It logs temperature, relative humidity, pressure drop across the filter, and triaxial accelerometry. BC was estimated during post-sampling processing via transmissometry (see below).

We logged 1-minute interval CO concentrations using the Lascar EL-USB-300 (Lascar Electronics, USA), which is the size of a large pen (125 × 26.4 × 26.4mm, 42g), runs on ½ AA batteries, and has a sensing range between 0 and 300 ppm. The Lascar CO device has been used extensively in HAP assessment (Chillrud et al. 2021; Fandiño-Del-Rio et al. 2017; Johnson et al. 2021).

### d. Sampling strategy

PM_2.5_, BC, and CO were measured at three 24-hour periods during pregnancy. Baseline measurements were made at greater than 9 and less than 20 weeks of gestation, prior to randomization. Follow-up, post-randomization measurements were made at 24–28 weeks of gestation and 32–36 weeks of gestation. At each monitoring period, pregnant participants were asked to wear a customized garment (Johnson et al. 2020) with instrumentation situated in the breathing zone (Balakrishnan et al. 2018; Delapena et al. 2018), and asked to keep them nearby (within 1-2m) when sleeping, bathing, or when conducting other activities for which the equipment could not be safely worn.

## Measurements

### *Determining* PM_2.5_ *mass concentrations*

At each visit, 24-hour gravimetric filter-based samples were collected for each participant. Changes in filter mass pre- and post-sampling were assessed using 1-μg resolution microbalances (Sartorius Cubis, MSA6.6s-000-DF, Göttingen, Germany) at the University of Georgia (filters for Guatemala, Rwanda, and Peru) and at the Sri Ramachandra Institute for Higher Education and Research (for India).

We assessed gravimetric data validity using a three stage process: 1) Field technicians evaluated pre- and post-sample flow rates with a primary flowmeter at the field office, enabling implementation of validation criteria to flag and remove samples outside of expected ranges; 2) laboratory technicians invalidated samples with damaged filters; 3) data analysts removed data that did not meet criteria for sample duration (24 ± 4 hours), flow rate (300 ± 100 ml/min, as measured by the internal flow sensor), and inlet pressure (95th percentile < 5 inches H2O) (additional details provided in SI Table S2).

For cases in which the gravimetric sample was invalidated (e.g., due to a missing or damaged filter or flow faults), the nephelometer data from the instrument were used to estimate personal exposure, requiring additional data validity checks. We evaluated the relationship between the nephelometer and gravimetric samples for each ECM monitor (n = 431) used in the study, deeming acceptable performance as follows: R^2^ values greater than 0.65; ≥ 3 available pairs of valid filter and nephelometric samples; and slopes between 0.5 and 2.5. Additional details available in the SI. For the ECM samplers that met these criteria (57.8%), regression models were applied to the adjusted 24-h average nephelometer values for those samples with missing or invalid gravimetric samples, resulting in instrument-specific nephelometric PM_2.5_ concentrations normalized to field-based filter samples.

### Quality control and assurance

Field blanks were collected at a rate of 4 per 100 sample filters. 393 field blanks, an average of 98 samples (SD 38) per IRC, were collected in total to perform median blank corrections by IRC (Table S1). The limit of detection (LoD) was calculated separately for each IRC as three times the standard deviation of the blank mass depositions, after removing unrealistic blank values (outliers with greater than 10 μg or less than -10 μg mass deposition). Sample depositions below the LoD were replaced with LoD/(2^0.5^) (Hornung and Reed 1990). Duplicate ECMs were deployed on a subset of samples (n = 253) to assess between-monitor performance (SI Table S3; SI Figure S1).

### Wearing compliance

Compliance, as measured by the ECM’s accelerometer, was not used for data exclusion, due to differences in wearing patterns by country and the difficulty in discerning whether stillness of the monitor was truly indicative of non-compliance or if the pregnant women were actually adjacent to the ECMs. Daytime compliance was calculated from the ECM files as per the algorithm presented in Johnson et al. 2020. Compliance summary statistics and distributions are presented in SI Table S4 and SI Figure S2.

### Black Carbon

Black carbon concentrations were estimated for PM_2.5_ filter samples using SootScan Model OT21 Optical Transmissometers (Magee Scientific, USA), either at the University of Georgia (UGA, Athens, GA, USA for samples collected in Guatemala, Peru, and Rwanda) or at Sri Ramachandra Institute for Higher Education and Research (SRIHER, Chennai, India) for samples collected in India. BC depositions were estimated per Garland et al. 2017, using the BC attenuation cross-section values for similar Teflon filters (σ_ATN_ = 13.7 μg/cm^2^) collected from similar source types. Most filters collected for the Guatemala, Peru, and Rwanda samples used both a pre- and post-scan (2672 (99.2%), 2232 (97.1%), and 2181 (98.9%), respectively), while India had 2443 (100%) without pre-scans due to equipment unavailability. For India, the average of blank filter post-scan values was substituted for pre-scan values. LoD was calculated as it was for gravimetric mass (three times the blank standard deviation) after filtering out outliers above 5 μg and below -5 μg deposition. Values below the LoD were replaced with LoD/(2^0.5^). Data exclusion details are presented in Table S5.

### Carbon Monoxide

CO data quality assurance procedures included calibrations with zero air and CO span gas (ranging between 40 and 80ppm by IRC); automated, server-based quality assurance checks at regular intervals; and a visual rating system similar to that applied in the Ghana Randomized Air Pollution and Health Study (Chillrud et al. 2021). CO loggers were to be calibrated every 1-3 months, as per Johnson et al., 2020. CO monitors were calibrated using the temporally closest calibration coefficient. Data were then checked for sampling duration (24h ± 4h) and visually rated to remove files (3.4%) which displayed response artifacts (described in the SI). Duplicate monitors were deployed for a subset of samples to assess monitor performance.

### Surveys

Oral surveys and observations were conducted at the end of each exposure period to collect information on other sources of exposure, compliance, and stove/fuel use patterns. Additional household, stove, and kitchen characteristics data were also collected at baseline.

### e. Statistical analysis

All analyses were performed in R (versions 3.6 and 4.0, R Foundation for Statistical Computing, Vienna, Austria). We first calculated pollutant-specific descriptive statistics for valid measurements in control and intervention groups by study phase (baseline versus post-intervention rounds) and IRC. We evaluated Spearman correlations between measurements for the same pollutants collected at baseline and post-intervention and correlations between pollutants at each measurement point. These were evaluated overall and stratified by assigned stove/fuel type. Differences in pollutant levels between control and intervention groups by period (i.e., at baseline, post-intervention visits 1 and 2) were evaluated using non-parametric tests (Wilcoxon Rank Sum, Kruskal-Wallis, and Dunn’s tests). We also evaluated the proportion of samples that were less than or equal to the annual WHO Interim Target 1 (WHO-IT1) guideline value of 35 μg/m^3^ for PM_2.5_ (WHO 2006) and the WHO 24-hour guideline value for of 7 mg/m^3^ (∼6.1 ppm) for carbon monoxide (WHO 2010).

Following approaches described in McCracken et al (2009) and Chillrud et al (2021), we used statistical methods that leverage our study design and repeat measurements to assess the impact of intervening with LPG on exposure to PM_2.5_, CO, and BC during gestation. Given the right-skewed distribution of measured data, pollutant concentrations were natural log transformed prior to regression analyses. We used linear mixed effects models to assess the impact of the intervention on log-transformed personal exposures and included a random intercept to account for correlation among repeated measurements made on the same participants (i.e., at baseline and post-intervention visits 1 and 2). We also evaluated non-transformed models to estimate the absolute change in exposures.

We fit four models, offering distinct comparisons for the association between the LPG intervention and exposures. A summary of models --including equations, data utilized, and rationale --is in SI Table S8. Model 1 estimates the effect of the intervention by comparing exposures in the treatment arms during the post-intervention period (“between groups”). The main parameter of interest from this model is the fixed effect for the treatment arm. Model 2 estimates the difference in exposure between post-intervention and baseline periods (“before and after”) separately for each treatment arm. In these models, the parameter of interest is the difference between baseline and post-baseline measurements for the intervention arm, and the same difference for the control arm. Model 3 estimates changes in exposure in the intervention arm, pre-versus post-intervention, relative to any changes experienced in the control arm over the same period (“comparison-of-changes”). In this model, the parameter of interest is the “treatment arm x period” interaction term, where period is either pre- or post-intervention, and which controls for potential differences at baseline. Model 4 estimates comparison-of-changes by study visit (the same as model 3, treating each post-intervention visit as its own time point). The parameter of interest is the “treatment arm x visit” interaction term. This approach enables evaluation of the stability of changes in exposure over time. Parameters of interest were exponentiated, subtracted from 1, and multiplied by 100 to estimate the percent reduction in personal exposure due to the intervention.

## 3. Results

### a. Household characteristics

Table 1 shows household and participant characteristics for control and intervention arms at the four IRCs. Additional covariates are in SI Tables S1 and S2. Balance was evident as expected between the arms within each IRC for the age of the pregnant women, as well as educational attainment and occupational status. Households typically cooked indoors; there was heterogeneity in fuel types between countries, with wood dominant in Guatemala and India, dung dominant in Peru, and wood and charcoal in use in Rwanda.

**Table 1:** Household and maternal characteristics at HAPIN baseline, by study site and intervention arm

|  | Guatemala |  | India |  | Peru |  | Rwanda |  |
| --- | --- | --- | --- | --- | --- | --- | --- | --- |
| Variable | Control<br>(N=400) | Intervention<br>(N=400) | Control<br>(N=399) | Intervention<br>(N=400) | Control<br>(N=402) | Intervention<br>(N=396) | Control<br>(N=404) | Intervention<br>(N=394) |
| **Household and kitchen characteristics** |  |  |  |  |  |  |  |  |
| **Household size** |  |  |  |  |  |  |  |  |
| Mean ± SD | 5.1 ± 2.6 | 5.3 ± 2.7 | 3.8 ± 1.5 | 3.7 ± 1.6 | 4.7 ± 1.8 | 4.5 ± 1.7 | 3.5 ± 1.5 | 3.5 ± 1.5 |
| Range | 2-18 | 2-17 | 2-9 | 1-10 | 2-12 | 2-11 | 1-10 | 1-10 |
| Missing | 0 | 0 | 0 | 0 | 0 | 1 | 0 | 0 |
| **Household wealth at national quintiles** |  |  |  |  |  |  |  |  |
| Lowest | 235 (59%) | 236 (59%) | 82 (21%) | 97 (24%) | 210 (52%) | 202 (51%) | 21 (5%) | 11 (3%) |
| Second lowest | 103 (26%) | 102 (26%) | 206 (52%) | 196 (49%) | 117 (29%) | 103 (26%) | 67 (17%) | 57 (14%) |
| Medium | 55 (14%) | 47 (12%) | 90 (23%) | 85 (21%) | 62 (15%) | 85 (21%) | 117 (29%) | 86 (22%) |
| Second highest | 7 (2%) | 15 (4%) | 21 (5%) | 22 (6%) | 13 (3%) | 6 (2%) | 149 (37%) | 142 (36%) |
| Highest | 0 (0%) | 0 (0%) | 0 (0%) | 0 (0%) | 0 (0%) | 0 (0%) | 50 (12%) | 98 (25%) |
| Missing | 0 | 0 | 0 | 0 | 0 | 0 | 0 | 0 |
| **Access to electricity** |  |  |  |  |  |  |  |  |
| No | 40 (10%) | 46 (12%) | 16 (4%) | 12 (3%) | 22 (5%) | 24 (6%) | 272 (72%) | 213 (58%) |
| Yes | 360 (90%) | 354 (89%) | 383 (96%) | 388 (97%) | 380 (95%) | 372 (94%) | 104 (28%) | 156 (42%) |
| Missing | 0 | 0 | 0 | 0 | 0 | 0 | 28 | 25 |
| **Kitchen volume (m<sup>3</sup>)<sup>a</sup>** |  |  |  |  |  |  |  |  |
| Mean (SD) | 33.4 ± 16.4 | 34.0 ± 17.2 | 20.1 ± 13.3 | 21.9 ± 15.9 | 19.9 ± 12.5 | 18.8 ± 11.7 | 12.3 ± 7.6 | 13.3 ± 7.0 |
| Range | 7-100 | 7-126 | 0-86 | 2-133 | 2-116 | 2-82 | 0-83 | 0-53 |
| N | 378 | 379 | 386 | 391 | 296 | 290 | 272 | 270 |
| Missing | 14 | 15 | 0 | 1 | 14 | 14 | 2 | 2 |
| **Roof in the kitchen** |  |  |  |  |  |  |  |  |
| No | 1 (<1%) | 1 (<1%) | 1 (<1%) | 3 ( 1%) | 88 (22%) | 91 (23%) | 124 (31%) | 119 (30%) |
| Yes | 395 (100%) | 398 (100%) | 398 (100%) | 397 (99%) | 312 (78%) | 305 (77%) | 279 (69%) | 274 (70%) |
| Missing | 4 | 1 | 0 | 0 | 2 | 0 | 1 | 1 |
| **Number of stoves** |  |  |  |  |  |  |  |  |
| None | 0 (0%) | 0 (0%) | 0 (0%) | 0 (0%) | 0 (0%) | 1 (<1%) | 1 (<1%) | 0 (0%) |
| One | 138 (35%) | 135 (34%) | 271 (68%) | 282 (71%) | 105 (26%) | 90 (23%) | 248 (62%) | 228 (58%) |
| Two or more | 262 (66%) | 265 (66%) | 128 (32%) | 118 (30%) | 295 (74%) | 304 (77%) | 154 (38%) | 165 (42%) |
| Missing | 0 | 0 | 0 | 0 | 2 | 1 | 1 | 1 |
| **Primary stove has a chimney** |  |  |  |  |  |  |  |  |
| No | 310 (78%) | 313 (78%) | 397 (99%) | 398 (100%) | 249 (62%) | 252 (64%) | 382 (95%) | 379 (96%) |
| Yes | 90 (23%) | 87 (22%) | 2 ( 1%) | 2 (1%) | 152 (38%) | 144 (36%) | 21 (5%) | 14 (4%) |
| Missing | 0 | 0 | 0 | 0 | 1 | 0 | 1 | 1 |
| **Primary fuel type** |  |  |  |  |  |  |  |  |
| Cow dung | 0 ( 0%) | 0 ( 0%) | 0 ( 0%) | 0 ( 0%) | 351 (88%) | 346 (87%) | 0 ( 0%) | 0 ( 0%) |
| Wood | 394 (99%) | 399 (100%) | 399 (100%) | 400 (100%) | 48 (12%) | 42 (11%) | 323 (80%) | 257 (65%) |
| Charcoal | 0 ( 0%) | 0 ( 0%) | 0 ( 0%) | 0 ( 0%) | 0 ( 0%) | 0 ( 0%) | 72 (18%) | 125 (32%) |
| Other | 2 ( 1%) | 1 (<1%) | 0 ( 0%) | 0 ( 0%) | 2 (<1%) | 8 ( 2%) | 8 ( 2%) | 11 ( 3%) |
| Missing | 4 | 0 | 0 | 0 | 1 | 0 | 1 | 1 |
| **Primary Stove Location** |  |  |  |  |  |  |  |  |
| In participant's bedroom | 18 (5%) | 18 (5%) | 78 (20%) | 88 (22%) | 5 ( 1%) | 4 ( 1%) | 6 ( 2%) | 2 ( 1%) |
| Room immediately adjacent to the participant's bedroom | 194 (49%) | 186 (47%) | 143 (36%) | 122 (31%) | 31 ( 8%) | 32 ( 8%) | 9 ( 2%) | 13 ( 3%) |
| Separated from the participant's bedroom but inside the house | 103 (26%) | 122 (31%) | 75 (19%) | 73 (18%) | 72 (18%) | 74 (19%) | 24 ( 6%) | 19 ( 5%) |
| Outside the house (outdoors) | 4 ( 1%) | 7 ( 2%) | 1 (<1%) | 4 ( 1%) | 86 (22%) | 79 (20%) | 125 (31%) | 130 (34%) |
| In a separate building detached from the bedroom-main home | 77 (19%) | 64 (16%) | 97 (25%) | 109 (27%) | 197 (50%) | 202 (52%) | 234 (59%) | 217 (57%) |
| Other | 0 ( 0%) | 0 ( 0%) | 0 ( 0%) | 1 (<1%) | 1 (<1%) | 0 ( 0%) | 0 ( 0%) | 0 ( 0%) |
| Missing | 0 | 0 | 0 | 0 | 0 | 1 | 0 | 1 |
| **Primary light source** |  |  |  |  |  |  |  |  |
| Torch (battery) | 5 ( 1%) | 5 ( 1%) | 4 ( 1%) | 7 ( 2%) | 4 ( 1%) | 8 ( 2%) | 90 (22%) | 83 (21%) |
| Kerosene lamp | 1 (<1%) | 5 ( 1%) | 9 ( 2%) | 7 ( 2%) | 1 (<1%) | 0 ( 0%) | 37 ( 9%) | 22 ( 6%) |
| Solar light | 0 ( 0%) | 2 ( 1%) | 2 ( 1%) | 1 (<1%) | 12 ( 3%) | 11 ( 3%) | 138 (34%) | 119 (30%) |
| Electricity | 361 (90%) | 348 (87%) | 384 (96%) | 385 (96%) | 371 (93%) | 359 (91%) | 88 (22%) | 138 (35%) |
| Other | 33 ( 8%) | 40 (10%) | 0 ( 0%) | 0 ( 0%) | 13 ( 3%) | 18 ( 5%) | 50 (12%) | 31 ( 8%) |
| Missing | 0 | 0 | 0 | 0 | 1 | 0 | 1 | 1 |
| **Presence of smoker in home** |  |  |  |  |  |  |  |  |
| No | 378 (95%) | 378 (95%) | 265 (66%) | 281 (70%) | 397 (99%) | 392 (99%) | 381 (95%) | 385 (98%) |
| Yes | 22 ( 6%) | 22 ( 6%) | 134 (34%) | 119 (30%) | 3 ( 1%) | 4 ( 1%) | 22 ( 5%) | 8 ( 2%) |
| Missing | 0 | 0 | 0 | 0 | 2 | 0 | 1 | 1 |
| **Maternal characteristics** |  |  |  |  |  |  |  |  |
| **Age (years)** |  |  |  |  |  |  |  |  |
| Mean ± SD | 25.0 ± 4.5 | 24.5 ± 4.4 | 23.9 ± 3.9 | 24.0 ± 3.7 | 25.4 ± 4.6 | 25.6 ± 4.3 | 27.3 ± 4.5 | 27.3 ± 4.3 |
| Range | 18-35 | 18-35 | 18-35 | 18-35 | 18-35 | 18-35 | 18-35 | 18-35 |
| Missing | 0 | 0 | 0 | 0 | 0 | 0 | 0 | 0 |
| **Occupation** |  |  |  |  |  |  |  |  |
| Household | 377 (94%) | 369 (92%) | 213 (53%) | 219 (55%) | 67 (17%) | 65 (16%) | 22 (5%) | 37 (9%) |
| Agriculture | 0 (0%) | 6 (2%) | 170 (43%) | 168 (42%) | 303 (75%) | 298 (75%) | 322 (80%) | 269 (68%) |
| Commercial | 6 (2%) | 11 (3%) | 3 (1%) | 1 (<1%) | 60 (15%) | 51 (13%) | 62 (15%) | 74 (19%) |
| Other or N/A | 16 (4%) | 13 (3%) | 13 (3%) | 12 (3%) | 22 ( 5%) | 20 (5%) | 25 (6%) | 37 (9%) |
| Missing | 1 | 1 | 0 | 0 | 0 | 0 | 0 | 0 |

<sup>a</sup>Kitchen with a dimension greater than 25 meters or less than 0.5 meter was considered unreasonable and a data entry error

### b. Exposure measurements, data completeness, and quality assurance and control

Across the baseline visit and first two post-intervention visits, HAPIN field staff made over 9000 exposure monitoring visits for 3195 pregnant women. 82% of the pregnant women had a valid baseline PM_2.5_ sample and at least one valid PM_2.5_ post-intervention sample. Approximately 14% of invalid gravimetric samples were replaced with ECM-specific, gravimetric-adjusted nephelometer values. For CO, 84% had a valid baseline sample and at least one valid post-intervention sample. For BC, 73% had a valid baseline measurement and at least one valid post-intervention measurement. The percentage of samples successfully collected - by treatment arm, measurement visit, and IRC - is displayed in Table 2. The final dataset as reported here includes 7673 PM_2.5_, 7165 BC, and 7943 CO samples for pregnant women. Details on exclusions are in the SI (Tables S2 and S6).

**Table 2.** Summarized Maternal exposures to PM2.5, Black Carbon, and Carbon Monoxide by IRC and Overall

| PM<sub>2.5</sub> Exposure | Guatemala |  | India |  | Peru |  | Rwanda |  | Overall |  |
| --- | --- | --- | --- | --- | --- | --- | --- | --- | --- | --- |
|  | Control | Intervention | Control | Intervention | Control | Intervention | Control | Intervention | Control | Intervention |
| **Baseline** |  |  |  |  |  |  |  |  |  |  |
| Enrolled Participants | 400 | 400 | 399 | 400 | 402 | 396 | 404 | 394 | 1605 | 1590 |
| N (% of enrolled) | 373 (93) | 360 (90) | 358 (90) | 357 (89) | 327(81) | 331 (84) | 364 (90) | 353 (90) | 1422 (89) | 1401 (88) |
| Average ± SD | 140 ± 133.) | 153 ± 118.7 | 103.9 ± 100.1 | 127.1 ± 181.3 | 80.2 ± 94.8 | 89.5 ± 116.8 | 115.6 ± 97.2 | 108 ± 98.4 | 110.9 ± 110 | 120.1 ± 134.8 |
| Range | 10.5 - 1799 | 9.9 - 780.3 | 10.8 - 1033.9 | 9.4 - 2099.9 | 10.7 - 697.1 | 11 - 1400.1 | 14.3 - 1089.8 | 14.2 - 865.6 | 10.5 - 1799 | 9.4 - 2099.9 |
| Median (IQR) | 110.2 (63.3-176) | 122.3 (65.8-200.2) | 73.8 (46.7-123.3) | 78.4 (47.9-142.9) | 46.1 (15.2-108.5) | 53.3 (22.8-111.2) | 94 (58.6-144.8) | 82.7 (47.8-134.1) | 83.1 (45.9-141.4) | 81.7 (45.9-150.8) |
| **Post-intervention Visit 1** |  |  |  |  |  |  |  |  |  |  |
| Enrolled Participants | 396 | 398 | 394 | 393 | 387 | 390 | 403 | 388 | 1580 | 1569 |
| N (% of enrolled) | 339 (86) | 361 (91) | 311 (79) | 314 (80) | 269 (70) | 289 (74) | 332 (82) | 321 (83) | 1251 (79) | 1285 (82) |
| Average ± SD | 133 ± 116.1 | 31.3 ± 33.4 | 102.9 ± 114.5 | 39.2 ± 39.1 | 64.5 ± 104.2 | 20.8 ± 19.3 | 108.9 ± 109.8 | 43.1 ± 32.3 | 104.4 ± 113.9 | 33.8 ± 33.1 |
| Range | 9.9 - 681.7 | 9.6 - 459 | 10.5 - 890.3 | 10.4 - 300.8 | 9.9 - 1116.8 | 9.6 - 258.8 | 14 - 1093 | 14.2 - 283.9 | 9.9 - 1116.8 | 9.6 - 459 |
| Median (IQR) | 98.4 (58.7-164.7) | 23.3 (14.9-36.2) | 67.3 (38.9-117.8) | 28.7 (16.9-45.5) | 31.4 (14.5-74.4) | 14.6 (14.1-23.1) | 79.6 (48.5-129.6) | 33.6 (23.8-50) | 71.5 (38.5-125.9) | 24.1 (15-39.5) |
| **Post-intervention Visit 2** |  |  |  |  |  |  |  |  |  |  |
| Enrolled Participants | 392 | 391 | 390 | 391 | 368 | 385 | 401 | 385 | 1551 | 1552 |
| N (% of enrolled) | 317 (81) | 330 (84) | 284 (73) | 293 (75) | 219 (60) | 264 (69) | 318 (79) | 289 (75) | 1138 (73) | 1176 (76) |
| Average ± SD | 123.9 ± 99.5 | 33.4 ± 37.9 | 109.3 ± 123.5 | 36.5 ± 39.6 | 67 ± 124.7 | 28.1 ± 77.1 | 99.4 ± 77.9 | 45 ± 58.1 | 102.5 ± 107.7 | 35.8 ± 54.6 |
| Range | 10.3 - 689.3 | 9.7 - 441.8 | 10.4 - 793.8 | 5.7 - 462.5 | 10.2 - 1208.4 | 10.5 - 850.8 | 14.6 - 664.7 | 12.5 - 751.7 | 10.2 - 1208.4 | 5.7 - 850.8 |
| Median (IQR) | 93.6 (53.6-168.2) | 23.8 (16.3-38.5) | 68.2 (36.3-129.3) | 25.3 (16.9-41.8) | 24.7 (14.5-57.9) | 14.6 (13.9-18.6) | 79.9 (45.9-128.1) | 28.2 (23.6-49.1) | 69.5 (36.5-130.8) | 23.7 (14.9-39.7) |

| BC Exposure | Guatemala |  | India |  | Peru |  | Rwanda |  | Overall |  |
| --- | --- | --- | --- | --- | --- | --- | --- | --- | --- | --- |
|  | Control | Intervention | Control | Intervention | Control | Intervention | Control | Intervention | Control | Intervention |
| **Baseline** |  |  |  |  |  |  |  |  |  |  |
| Enrolled Participants | 400 | 400 | 399 | 400 | 402 | 396 | 404 | 394 | 1605 | 1590 |
| N (% of enrolled) | 343 (86) | 332 (83) | 351 (88) | 348 (87) | 289 (72) | 307 (78) | 289 (72) | 280 (71) | 1272 (79) | 1267 (80) |
| Average ± SD | 13 ± 7.5 | 13.4 ± 10.7 | 12.3 ± 9.9 | 13.5 ± 12.6 | 11 ± 11.6 | 11.6 ± 11.2 | 13 ± 8.4 | 11.5 ± 8.8 | 12.4 ± 9.4 | 12.6 ± 11 |
| Range | 2.5 - 95.6 | 2.6 - 132.6 | 0.7 - 73.2 | 0.6 - 102.7 | 1.5 - 74.9 | 1.5 - 75.3 | 2.7 - 70.4 | 2.7 - 76.9 | 0.7 - 95.6 | 0.6 - 132.6 |
| Median (IQR) | 12.1 (9.1-15.3) | 11.7 (9.4-14.8) | 9.4 (5.5-15.7) | 9.8 (5.5-16.4) | 7.9 (2.3-15.7) | 8.7 (3.6-15.5) | 11.8 (8.3-15.6) | 9.7 (6.6-14.2) | 10.8 (6.8-15.5) | 10.5 (6.2-15.3) |
| **Post-intervention Visit 1** |  |  |  |  |  |  |  |  |  |  |
| Enrolled Participants | 396 | 398 | 394 | 393 | 387 | 390 | 403 | 388 | 1580 | 1569 |
| N (% of enrolled) | 330 (83) | 359 (90) | 304 (77) | 305 (78) | 246 (64) | 268 (69) | 307 (76) | 294 (76) | 1187 (75) | 1226 (78) |
| Average ± SD | 12.3 ± 6.6 | 4.9 ± 7.5 | 11.1 ± 10.3 | 3.5 ± 4.7 | 8.5 ± 10.1 | 1.9 ± 1.1 | 11.8 ± 10.6 | 5.3 ± 5 | 11.1 ± 9.6 | 4 ± 5.5 |
| Range | 2.6 - 52.9 | 2.2 - 131.5 | 0.7 - 98.5 | 0.7 - 44.6 | 1.4 - 73.6 | 1.4 - 13.8 | 2.8 - 122 | 2.6 - 54.7 | 0.7 - 122 | 0.7 - 131.5 |
| Median (IQR) | 11.5 (8.1-15.1) | 2.7 (2.6-5.3) | 8.9 (4.5-14.5) | 2.1 (1.1-3.6) | 4.4 (1.6-12.1) | 1.6 (1.5-1.6) | 10 (7-13.9) | 4.2 (2.9-5.9) | 9.7 (5.3-14.4) | 2.7 (1.6-4.7) |
| **Post-intervention Visit 2** |  |  |  |  |  |  |  |  |  |  |
| Enrolled Participants | 392 | 391 | 390 | 391 | 368 | 385 | 401 | 385 | 1551 | 1552 |
| N (% of enrolled) | 310 (79) | 326 (83) | 277 (71) | 289 (74) | 201 (55) | 242 (63) | 291 (73) | 277 (72) | 1079 (70) | 1134 (73) |
| Average ± SD | 11.9 ± 7.1 | 5 ± 4.9 | 11.8 ± 12.3 | 4.2 ± 7.6 | 8.7 ± 13.8 | 2 ± 1.8 | 11.2 ± 7.1 | 5.5 ± 4.9 | 11.1 ± 10.2 | 4.3 ± 5.4 |
| Range | 2.5 - 88.1 | 2.5 - 64.8 | 0.7 - 97.8 | 0.6 - 105.2 | 1.3 - 123.5 | 1.4 - 14.5 | 2.8 - 61.5 | 2.5 - 44 | 0.7 - 123.5 | 0.6 - 105.2 |
| Median (IQR) | 11.1 (8.4-14.5) | 2.9 (2.6-5.7) | 8.2 (4.4-14) | 2.5 (1.5-4.1) | 3.7 (1.6-10.8) | 1.6 (1.5-1.6) | 10.3 (6.4-13.7) | 3.8 (2.9-6.2) | 9.6 (5.2-13.7) | 2.8 (1.7-4.8) |

| CO Exposure | Guatemala |  | India |  | Peru |  | Rwanda |  | Overall |  |
| --- | --- | --- | --- | --- | --- | --- | --- | --- | --- | --- |
|  | Control | Intervention | Control | Intervention | Control | Intervention | Control | Intervention | Control | Intervention |
| **Baseline** |  |  |  |  |  |  |  |  |  |  |
| Enrolled Participants | 400 | 400 | 399 | 400 | 402 | 396 | 404 | 394 | 1605 | 1590 |
| N (% of enrolled) | 382 (96) | 375 (94) | 372 (93) | 373 (93) | 333 (83) | 326 (82) | 360 (89) | 356 (90) | 1447 (90) | 1430 (90) |
| Average ± SD | 2 ± 3.7 | 2.1 ± 2.3 | 1.8 ± 3.6 | 1.7 ± 2.6 | 3.5 ± 5 | 4.4 ± 7.5 | 2 ± 3.3 | 3 ± 4.8 | 2.3 ± 4 | 2.7 ± 4.8 |
| Range | 0 - 60.2 | 0 - 21.7 | 0 - 46.9 | 0 - 31.6 | 0 - 54.3 | 0 - 69.5 | 0 - 29.6 | 0 - 44.4 | 0 - 60.2 | 0 - 69.5 |
| Median (IQR) | 1.3 (0.5-2.5) | 1.4 (0.6-2.8) | 0.8 (0.3-1.9) | 0.9 (0.3-2.1) | 1.9 (0.7-4.3) | 1.9 (0.8-4.8) | 1.1 (0.5-2) | 1.2 (0.5-3) | 1.2 (0.5-2.5) | 1.3 (0.5-3) |
| **Post-intervention Visit 1** |  |  |  |  |  |  |  |  |  |  |
| Enrolled Participants | 396 | 398 | 394 | 393 | 387 | 390 | 403 | 388 | 1580 | 1569 |
| N (% of enrolled) | 355 (90) | 363 (91) | 352 (89) | 347 (88) | 269 (70) | 264 (68) | 335 (83) | 341 (88) | 1311 (83) | 1315 (84) |
| Average ± SD | 1.9 ± 2.2 | 0.5 ± 1 | 1.9 ± 3.2 | 0.4 ± 1.4 | 3.3 ± 6.7 | 1.4 ± 2.4 | 2.1 ± 3.3 | 0.6 ± 1.2 | 2.2 ± 4.1 | 0.7 ± 1.5 |
| Range | 0 - 21.7 | 0 - 11.9 | 0 - 24.7 | 0 - 17.3 | 0 - 64.2 | 0 - 23.9 | 0 - 25.9 | 0 - 9.3 | 0 - 64.2 | 0 - 23.9 |
| Median (IQR) | 1.2 (0.5-2.5) | 0.1 (0-0.5) | 0.8 (0.2-2.1) | 0 (0-0.3) | 1.2 (0.3-3.4) | 0.7 (0.2-1.8) | 1 (0.4-2.1) | 0.2 (0.1-0.7) | 1.1 (0.4-2.5) | 0.2 (0-0.7) |
| **Post-intervention Visit 2** |  |  |  |  |  |  |  |  |  |  |
| Enrolled Participants | 392 | 391 | 390 | 391 | 368 | 385 | 401 | 385 | 1551 | 1552 |
| N (% of enrolled) | 336 (86) | 344 (88) | 320 (82) | 313 (80) | 212 (58) | 250 (65) | 345 (86) | 320 (83) | 1213 (78) | 1227 (79) |
| Average ± SD | 1.7 ± 2.1 | 0.6 ± 1.1 | 2 ± 3.8 | 0.4 ± 0.9 | 3.5 ± 6.3 | 1.2 ± 2.1 | 2.2 ± 3.6 | 0.6 ± 1 | 2.2 ± 4 | 0.7 ± 1.3 |
| Range | 0 - 14.4 | 0 - 8.9 | 0 - 35.9 | 0 - 6.9 | 0 - 43.7 | 0 - 21.2 | 0 - 26.4 | 0 - 6.7 | 0 - 43.7 | 0 - 21.2 |
| Median (IQR) | 1.2 (0.4-2.2) | 0.2 (0-0.6) | 0.7 (0.1-2.2) | 0 (0-0.2) | 1.5 (0.5-3.3) | 0.6 (0.1-1.3) | 1.1 (0.4-2.2) | 0.2 (0.1-0.7) | 1.1 (0.3-2.3) | 0.2 (0-0.7) |

‘Enrolled participants’ are the number of pregnant women participating in each IRC and treatment arm at each measurement round. Deviations from the overall number of participants at baseline are due to study exits. ‘N’ is the number of valid measurements by IRC and treatment arm in each measurement round. ‘% of enrolled’ is the ‘N’ / ‘Enrolled Participants’.

Duplicate gravimetric PM_2.5_ samples had correlation coefficients (R^2^) of 0.96, 0.81, and 0.93, and RMSE values of 18.2, 36.5, and 17.4 μg/m^3^ for Guatemala, Peru, and Rwanda, respectively. India had limited data available through this point of study to allow duplicate sample analysis. The duplicate CO samples had correlation coefficients (Pearson’s R) of 0.66, 0.79, and 0.60, and RMSE values of 1.4, 2.4, and 2.2 ppm for Guatemala, Peru, and Rwanda, respectively (India had limited data available for duplicate analysis; SI Figure S4 and Tables S6 and S7).

### c. Exposure summary

We summarized personal exposures to PM_2.5_, BC, and CO exposures for pregnant women by IRC and visit in Table 2. Exposure distributions are displayed graphically in Figure 1 (IRC-specific plots are in Figure S5). HAPIN-wide, there was no significant difference between baseline PM_2.5_ exposures (Wilcoxon rank sum, p = 0.463) in the control (median 83.1 μg/m^3^, IQR 45.9 - 141.4) and intervention arms (median 81.7 μg/m^3^, IQR 45.9 - 150.8), nor for BC (Wilcoxon rank sum, p = 0.436), with median control exposures estimated at 10.8 μg/m^3^ (IQR 6.8 - 15.5) and 10.5 μg/m^3^ (IQR 6.2 - 15.3) in the intervention arm. CO exposures were significantly higher (Wilcoxon rank sum test, p = 0.03), although still similar, in the intervention arm (1.3 ppm, IQR 0.5 - 3) at baseline versus the control arm (1.2 ppm, IQR 0.5 - 2.5).

**Figure 1.**
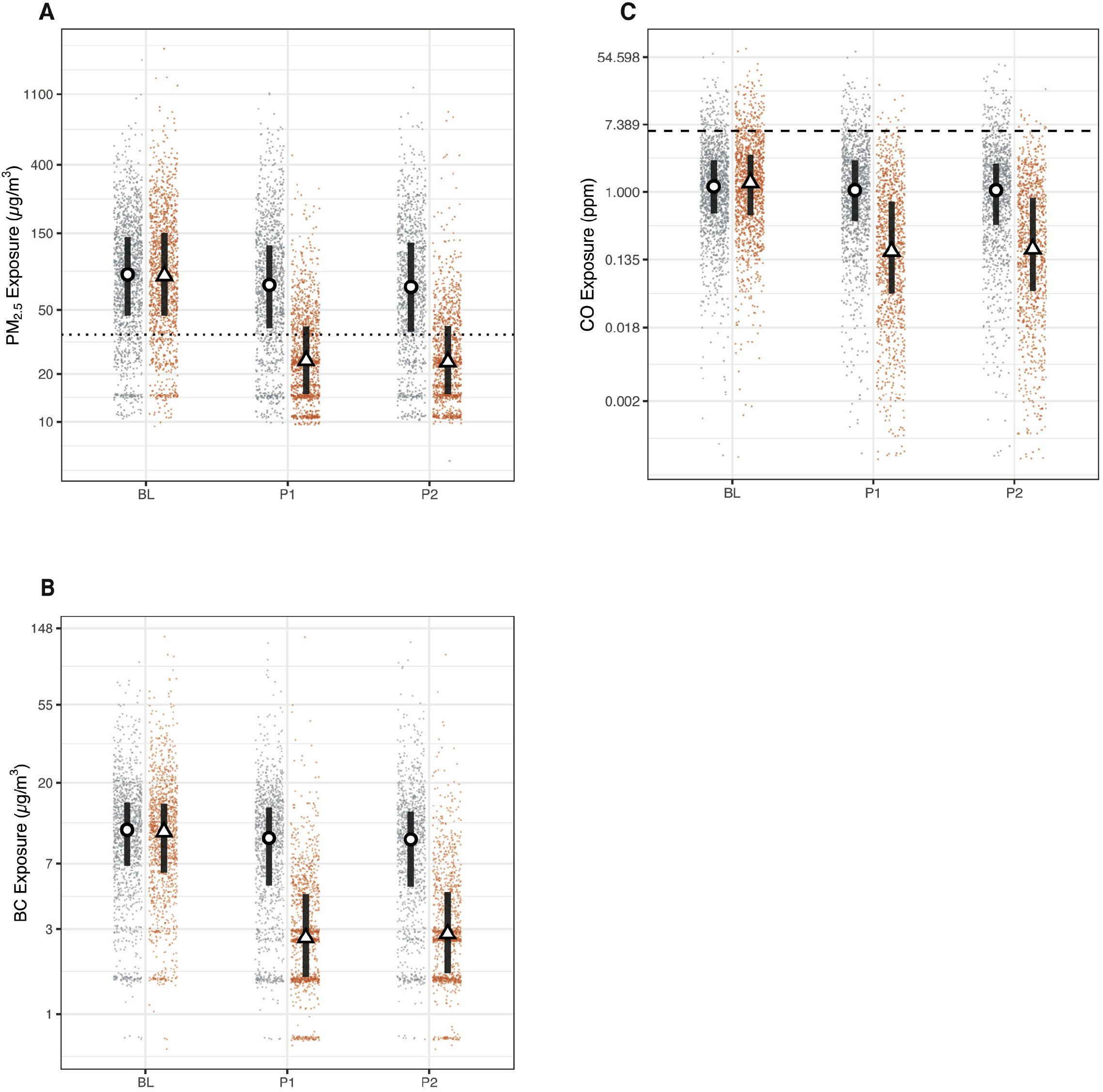
HAPIN (A) PM_2.5_, (B) Black Carbon (BC), and (C) Carbon Monoxide (CO) exposures. Red triangles and blue dots are study round samples in intervention and control households, respectively. Circles and triangles outlined in black are median values in control and intervention households, respectively. Lines are interquartile ranges. BL = baseline (9-20 weeks gestation), P1 = post-intervention visit 1 (24-28 weeks gestation), and P2 = post-intervention visit 2 (32-36 weeks gestation). The dotted line in the PM panels is the annual WHO Interim Target 1 guideline value (35 μg/m^3^); the dashed line in the CO plots is the WHO guideline value of 6.11 ppm (7 mg/m^3^).

During the baseline period, approximately 17% of measurements in both the control and intervention arms had PM_2.5_ exposures less than or equal to WHO-IT1. During the post-intervention period, 23% of control exposures were at or below the annual WHO-IT1; 69% of intervention exposures fell below this value.

At baseline, 92% and 90% of 24-hr exposures to CO in the control and intervention arms, respectively, were below the WHO guideline value for CO. Post-intervention, 93% of control exposures were below the guideline value, while 99% of intervention exposures were less than the guideline.

#### Exposures over time

Trial-wide and within IRCs, we observed changes in PM_2.5_ exposures between baseline and post-intervention rounds. The magnitude, consistency, and significance of these changes varied by study site and arm.

##### Study-wide

We plotted personal exposures to PM_2.5_ over time after randomization trial-wide (Figure 2) and by IRC (SI Figure S4), highlighting the relative overlap in exposures during the baseline period and the distinct separation of exposures between control and intervention groups after intervention. Baseline exposures were not significantly different (p ∼ 0.5) between control and intervention households. While the magnitude of the exposures and the exposure contrast vary between sites post-randomization, we note the relative stability of exposures across control and intervention arms. The mean absolute difference between post-intervention measurements taken in the same household was 35 μg/m^3^ (SD 31) and 36 μg/m^3^ (SD 54) for intervention and control households, respectively. Additional details by IRC and pollutant are in the SI.

**Figure 2.**
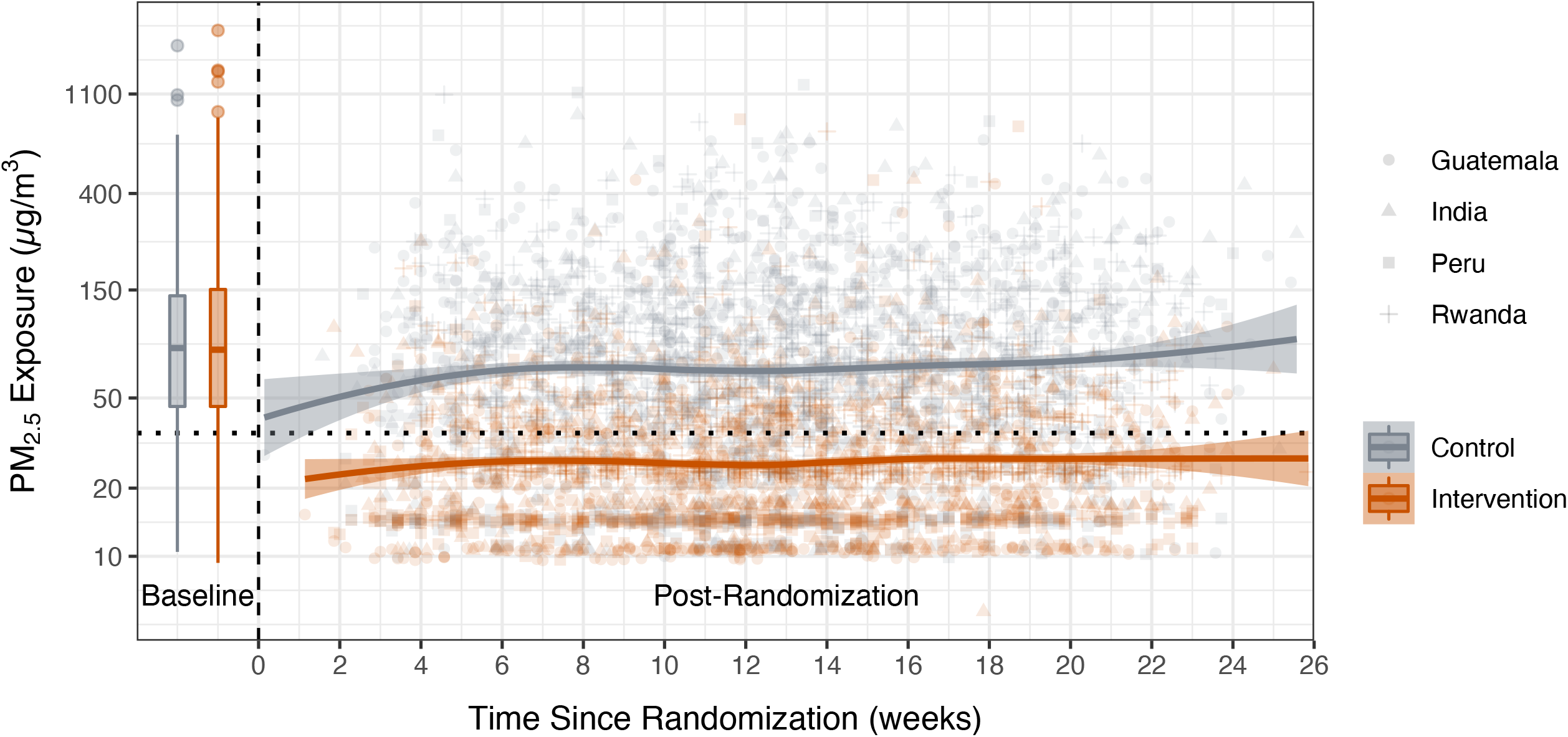
Trends in PM_2.5_ exposure. The x-axis is the time since randomization; time before 0 indicates the baseline period. Data from the baseline period are presented as box plots. The lower and upper hinges correspond to the first and third quartiles (the 25th and 75th percentiles). The upper and lower whiskers extend 1.5 * IQR above and below the upper and lower hinges. Data beyond the whiskers are outliers. Solid lines are a locally weighted smoothing (LOESS) function. Shaded areas are standard errors. Grey points are individual data points from control households; red points are from intervention households. The shape of the points indicates the IRC where the sample was collected.

##### Control households

There was a significant reduction (p < 0.001) in PM_2.5_ and BC between measurement rounds. The magnitude of this reduction was relatively small for PM_2.5_ (approximately 5% decrease in means and 14% decrease in medians between baseline and post-intervention round 1; and 2% and 3% decrease in means and medians, respectively, between post-intervention rounds 1 and 2) and for BC (approximately 10% between baseline and post-intervention round 1). For CO, there was a significant reduction (p ∼ 0.001) between baseline and post-intervention visit 2.

##### Intervention households

There was a significant (p < 0.001) and large decrease in PM_2.5_ exposures between baseline and post-intervention measurements. Mean exposures decreased by 86 μg/m^3^ (72%) between baseline and post-intervention visit 1 and by 84 μg/m^3^ (70%) when comparing baseline with post-intervention visit 2. Similar trends held for other measured pollutants.

Across all sites, BC values differed significantly between baseline and post-intervention visit 2 (p < 0.05). Additionally, in Rwanda and Peru, baseline and post-intervention visit 1 BC exposures differed significantly (p < 0.01 and p < 0.001, respectively).

#### Correlations between measurement rounds

Correlations (Spearman’s ρ) between measurement rounds were moderate. For PM_2.5_ in the control arm, correlations between baseline and post-intervention round 1, baseline and post-intervention round 2, and post-intervention rounds 1 and 2 were 0.42, 0.40, and 0.51, respectively. Correlations for BC in the control arm were weaker (0.29, 0.33, and 0.47), as were correlations for CO (0.29, 0.26, and 0.29).

Among intervention households, correlations between baseline and post-intervention round 1, baseline and post-intervention round 2, and post-intervention visits 1 and 2 were 0.21, 0.18, and 0.39, respectively. BC values followed a similar trend (0.18, 0.11, and 0.56 for the same comparisons) as did CO (0.14, 0.12, and 0.30). Weak correlations between baseline and post-intervention rounds among intervention households were expected, as the intervention was placed and in use after baseline but prior to post-intervention measurements.

#### Correlations between pollutants

The relationship between PM_2.5_ and CO among biomass using households (intervention arm at baseline; control arm at baseline and post-intervention rounds) was moderate (Spearman ρ ∼ 0.5), though much stronger than in LPG using households (Spearman ρ ∼ 0.06, SI Figure S6). For the relationship between PM and BC, across all measurements with biomass, the Spearman ρ was 0.8, while it was 0.7 among LPG users. More details on these relationships between pollutants is available in the SI (Figures S6, S7).

### d. Modeling Results

#### The effect of the LPG stove and fuel intervention on personal exposures

All models of the impact of the HAPIN LPG fuel and stove intervention indicated significant reductions in all measured pollutants. A discussion of PM_2.5_-specific results follow; results for CO and BC are similar (SI Figures S8 and S9; Tables S9 and S10).

Table 3 and Figure 3 report results from the between groups, before-and-after, and comparison-of-changes modeling approaches. Estimates of the percent reduction in PM_2.5_ exposure due to the intervention were similar across models: 61% (95% CI 59–63%) for the ‘between groups’ approach; 68% (95% CI 66–69%) for the ‘before-and-after’ approach; and 62% (95% CI 59– 64%) for the ‘comparison-of-changes’ approach.

**Table 3.** Percent decreases in PM2.5 exposure associated with the HAPIN LPG intervention.

| Model Type | Details | Percent decrease<br>in<br>PM<sub>2.5</sub> Exposure |
| --- | --- | --- |
|  |  | Estimate (CI) |
| Between Groups |  | 61 (59, 63) |
| Before and After | Control | 15 (12, 19) |
|  | Intervention | 68 (66, 69) |
| Comparison-of-changes | Overall | 62 (59, 64) |
|  | Visit 1 | 62 (59, 65) |
|  | Visit 2 | 61 (58, 64) |

**Figure 3.**
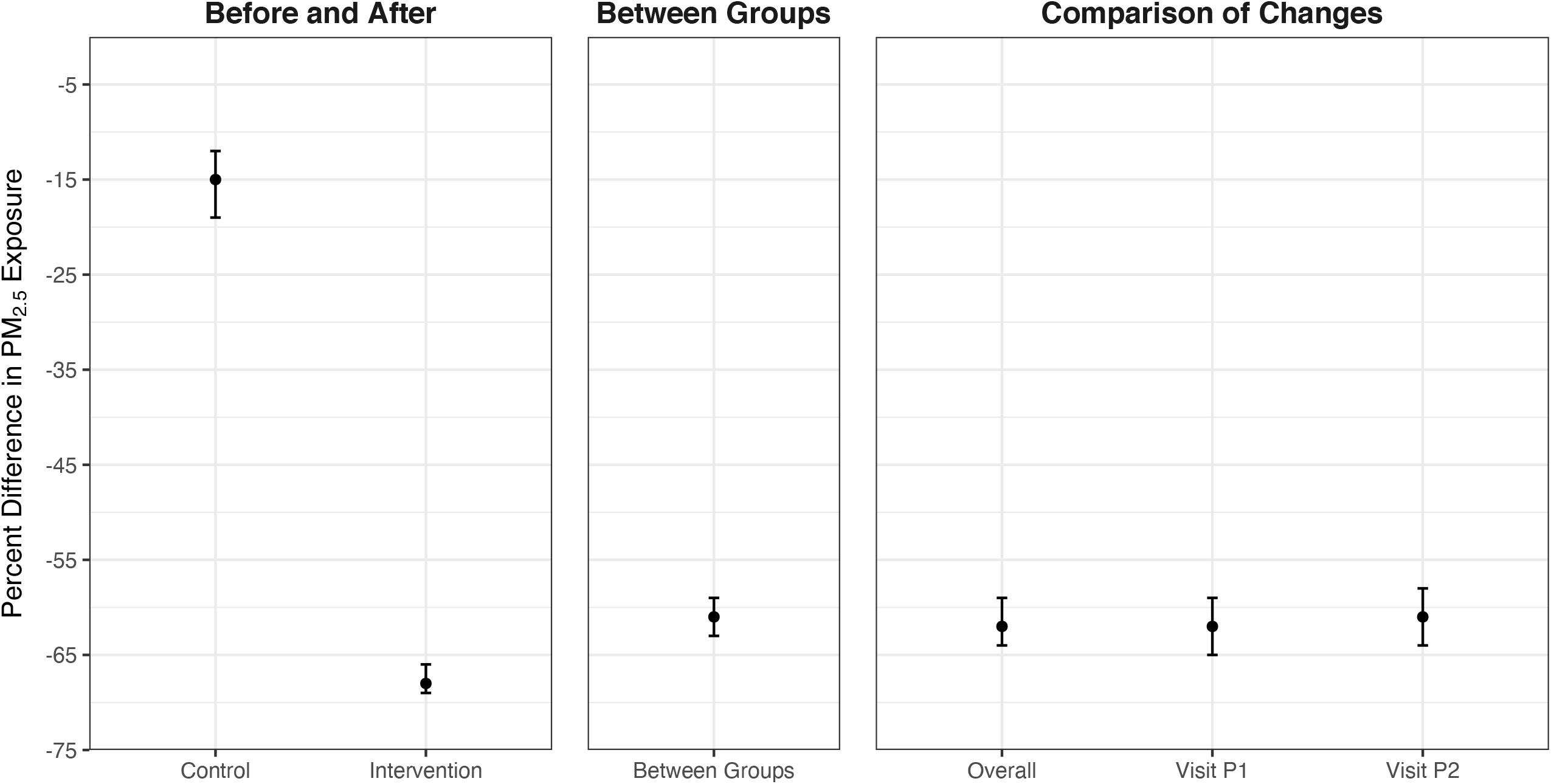
Estimated impacts of the HAPIN LPG intervention on PM_2.5_ exposure. All linear mixed effects models had log transformed PM_2.5_ as the dependent variable. Whiskers are 95% confidence intervals. The first panel (“Before and After”) uses data from both the control and intervention arms and compares the intervention period to the baseline period. The second panel (“Between Groups”) uses only data from the intervention period and contrasts the intervention arm with the control arm. The third panel (“Comparison-of-Changes”) uses all data from both study arms and both study periods; the model term of interest is the interaction between study arm and period, after controlling for each variable separately in the model. The “Overall” points consider an average post-intervention exposure; the Visit-specific points consider each post-randomization visit separately.

In models with untransformed outcomes, these percent reductions translate to absolute PM_2.5_ reductions of 68 μg/m^3^ (95% CI 63 – 74), 86 μg/m^3^ (95% CI 80 – 91), and 76 μg/m^3^ (95% CI 68 – 85), respectively. Of note, the before-and-after approach indicated a 15% (95% CI 12–19%) reduction in exposures between baseline and post-intervention periods for the control group (10 μg/m^3^ (95% CI 3 - 16). The visit-specific comparison-of-changes models (labeled Visit P1 and Visit P2 in Figure 3 and Table 3) were relatively consistent, indicating little to no change in intervention effectiveness over time. Models are presented separately for each IRC in the SI (SI Tables S11-S13; Figures S10-S12).

## 4. Discussion

### a. Exposure comparisons with previous studies

The HAPIN intervention of a free LPG stove and fuel supply, along with behavior change efforts, resulted in substantial and significant personal exposure reductions for pregnant women receiving the intervention when compared to the control arm for all pollutants and in all countries. Median PM_2.5_ post-intervention exposure measurements, approximately three months apart, were relatively consistent, suggesting the intervention had a stable effect through pregnancy. In total, these findings indicate consistent exposure reductions to near or below the annual WHO-IT1 guideline value of 35 μg/m^3^.

The exposure concentrations in the intervention arm were at the lower end of what has been reported for LPG or other clean fuel interventions, with 69% of all 24-hr PM_2.5_ samples falling below the annual WHO-IT1 target of 35 μg/m^3^. A systematic review by Pope et al. (2021) reported a pooled mean of 59 μg/m^3^ for the six LPG studies included in the analysis. CO exposures were similarly low for HAPIN, with a post-intervention median of 0.2 ppm, lower than the single study of 0.68ppm for the LPG studies (Pope et al. 2021).

A few recent HAP studies are of special interest given their scope and sample size, even without the same focus we placed on near-exclusive use of LPG. The Prospective Urban and Rural Epidemiological (PURE) study conducted observational PM_2.5_ and BC exposure measurements across 120 different communities (∼2500 homes) in Bangladesh, Chile, China, Colombia, India, Pakistan, Tanzania, and Zimbabwe (Shupler et al. 2020). The reported PM_2.5_ geometric means for women’s exposure among wood users were 89, 39, and 153 μg/m^3^ in India, South America, and Africa; among LPG users, exposures were 70, 32, and 146 μg/m^3^, respectively (Shupler et al. 2020). This study also offers the best comparison for BC exposures (as elemental carbon), with women’s geometric mean exposures in primarily wood using homes reported at 2.5-8.8 μg/m^3^ and 2.0-4.5 μg/m^3^ for the same user/region groupings. We estimated median BC exposures at 4-12 μg/m^3^ for all wood-using households (control and intervention groups at baseline and controls post-intervention) and 2-10 μg/m^3^ post-intervention. The trend of higher PM_2.5_ exposures in Africa and lower exposures in Latin America is similar to what was observed in HAPIN, although the post-invention exposures in HAPIN were substantially lower than those reported in PURE, assuming that the geometric means and medians are estimating similar central tendencies. Similarly, our post-intervention PM_2.5_ exposures in the intervention arm were lower than those reported for the Ghana Randomized Air Pollution and Health Study (GRAPHS), which included an LPG arm of 361 pregnant women (Chillrud et al. 2021). GRAPHS’ median 24-hour PM_2.5_ exposures for women in control homes using traditional biomass were 67 μg/m^3^ (HAPIN median baseline and control exposures: 67-140 μg/m^3^) and 45 μg/m^3^ post-LPG intervention (HAPIN mean post-intervention exposures: 20-45 μg/m^3^). GRAPHS reported women’s median CO exposures at 0.82 ppm in control homes and 0.52 ppm post-intervention in the LPG arm, compared to 1.95ppm in the controls and 0.2 post-intervention in the LPG arm for HAPIN.

Comparisons with exposure estimates from studies in similar regions also suggest the HAPIN intervention performed well in terms of exposure reductions. In Peru, the Cardiopulmonary outcomes and Health and Air Pollution (CHAP) trial reported median PM_2.5_ exposures of 57, 61, and 71 μg/m^3^ for the primary biomass-using control arm at 3, 6, and 12 months post-intervention, respectively, while the LPG-arm exposures were reported as 25, 16, and 15 μg/m^3^ (compared to medians of 31-25 in the control arm and 15 μg/m^3^ in the intervention arm for the HAPIN Peru site) (Checkley et al. 2020). In Rwanda, a trial of rocket-style cookstoves and water filters reported median exposures of 158 and 146 μg/m^3^ in the control and intervention arms, respectively, for the primary cook (compared to 80 μg/m^3^ in the control arm and 28-34 μg/m^3^ post-intervention for the HAPIN site in Rwanda) (Kirby et al. 2019). A study of pregnant women in Guatemala reported median exposures of 148 μg/m^3^ for open fire users and 55 μg/m^3^ for those using LPG (compared to 94-98 μg/m^3^ in the control arm and 23-24 μg/m^3^ in the intervention arm for the HAPIN site in Guatemala) (Grajeda et al. 2020), while in India the Tamil Nadu Air Pollution and Health Effects (TAPHE) cohort study of pregnant women estimated median PM_2.5_ exposures of 75 μg/m^3^ for biomass stove users and 46 μg/m^3^ for those using primarily LPG (compared to 67-68 and 25-29 μg/m^3^ in the control and intervention arms, respectively, for the HAPIN site in Tamil Nadu, India) (Balakrishnan et al. 2018).

There are several potential reasons for the differences in the reported exposures, especially for PM_2.5_, between these studies and HAPIN. Perhaps most importantly, as an efficacy trial, HAPIN has a strong emphasis on supporting exclusive LPG use, with free provision and delivery of stoves and fuel supply. Consistent usage was supported by behavior change strategies and stove repair or maintenance. Continuous biomass stove use monitoring was conducted in all intervention households, with reinforcement of exclusive LPG use provided when any biomass stove use was detected in a participant’s home. The approach resulted in near-exclusive gas stove use through pregnancy in intervention households: monitoring of biomass stoves in the LPG intervention homes indicated traditional stove use on less than 3% of days during gestation (Quinn et al. In Submission).

Other contextual factors are also important. In GRAPHS, for example, there was a high proportion of households cooking outdoors, which could imply lower baseline exposures, and homes were close together, which may have mitigated potential exposure contrasts due to ‘neighborhood’ effects. PURE was an observational study; while the groups provided a basis for comparison, there was no intervention effect to measure. And perhaps most importantly, stove use in the groupings was likely mixed, which could explain the higher exposures for the LPG users.

Finally, we note that our exposures for pregnant women in biomass-using homes (at baseline and post-randomization in the control group), were also somewhat lower than typically reported (mean of 116 μg/m^3^ and median of 83 μg/m^3^). The Pope et al. (2021) review reported a pooled mean of 220 μg/m^3^ for the baseline personal (biomass-using) exposures in the six LPG intervention studies; other reviews of HAP exposure have reported similar estimates (Balakrishnan et al. 2014; Pope et al. 2017; Quansah et al. 2017). It is possible that our field sites are contextually different from previous studies given HAPIN’s formative work to identify locations with low background concentrations and relatively low-density housing. There may also be secular changes and/or differences in measurement approaches contributing to these differences, although it is unclear what and how these specific factors would result in these differences.

### b. Multi-pollutant relationships

Correlations between co-emitted pollutants have been used to justify measurement of HAP exposure proxies, most commonly CO as a surrogate for PM_2.5_ (Clark et al. 2013; Dionisio et al. 2012; McCracken et al. 2013). CO is of interest given its relative ease of measurement compared to PM_2.5_, although a systematic review of this approach by Carter et al. (2017), found that the PM_2.5_-CO exposure correlations varied widely (Pearson’s R range 0.22-0.97). This broad range in the strength of the relationship is likely due to variability in combustion (including predominant fuel types and mixes contributing to HAP) and subsequent exposures, as well as the reliability of measurements.

We present a dataset of four unique settings where transitions from biomass (primarily wood) to LPG allow for a clear comparison between locations and fuel use types (SI Figures S5 and S6). Our findings among biomass-using households fall into the middle of the range described by Carter et al. (2017) and were stronger among biomass-using households than among LPG users. Few analyses characterize HAP exposure relationships between PM_2.5_ and BC, with the largest coming from the PURE study. They reported spearman ρ correlations of 0.65-0.9 (Shupler et al. 2020), similar to our findings.

### c. Study limitations

While this study represents one of the largest efforts to characterize the impact of a household energy intervention on personal exposures, there are still several considerations for interpreting our findings. First, HAPIN is an efficacy trial, in which the stove, fuel, and support services were provided for free, resulting in high intervention fidelity and minimal stove stacking with biomass through pregnancy (Quinn et al. In Submission). It is unclear if exposure reductions with an LPG intervention, as reported here, could be achieved in most contexts without similar support. The field sites were also specifically vetted for their likelihood to have low background air pollution levels (Clasen et al. 2020; Johnson et al. 2020). While it is hoped that comprehensive, community-scale interventions may reduce HAP’s contribution to reduced ambient air quality, and thus further reduce exposures, this is a largely untested hypothesis, and for many areas background concentrations are high due to emissions from a variety of sources (Piedrahita et al. 2017; Rooney et al. 2019), limiting potential exposure reductions for even the cleanest household energy interventions.

The relatively large number of exposure samples collected, analyzed, and reported here represent only three snapshots of exposure over several months for households in diverse settings. Behavioral and environmental factors change over time, resulting in some risk of exposure misclassification. Still, the high fidelity to the intervention and relatively stable exposures evident in Figures 2 and 3 suggest that our limited measurements provided reasonable exposure estimates over the pregnancy period. A subset of more intensive measurements (twice the number of measurements in 10% of the study population) is being conducted and will characterize how well our standard sampling protocol performs in predicting the longer and more intense exposure monitoring of the subset.

With the large number of samples being collected, some sample loss was inevitable. Approximately 19%, 16%, and 24% of the PM_2.5_, CO, and BC samples were invalid due to being missing, equipment failure, damaged or misplaced filters, or failure to meet quality assurance criteria. This level of missing data is not unexpected given the large-scale nature of the assessment, and having been conducted across our four diverse international research sites. The PURE and GRAPHS studies, for example, both reported over 80% (exact figures were not provided) of their PM_2.5_ samples as valid, with GRAPHS also reporting between 47-70% of the CO deployments as valid across the various sampling sessions (Chillrud et al. 2021; Shupler et al. 2020).

Finally, our study population for this analysis was pregnant women, a subgroup which has different behavioral considerations to others in the home. These exposures are clearly relevant for birthweight and other maternal and child health outcomes, but generalizability to other populations – or even for the same women post-pregnancy – may be limited due to differences in behavior during pregnancy which may impact HAP exposure (e.g. cooking, occupational, domestic, childcare other tasks) (Clark et al. 2013).

### d. Conclusions

The results presented here suggest that an LPG intervention can substantially reduce pregnant women’s exposures to health damaging pollutants. These exposure reductions represent, to our knowledge, some of the largest for a household energy intervention. While HAPIN is an efficacy trial with specific contextual considerations that limit the generalizability of the results, our findings demonstrate that, in four geographic regions with different behavioral, sociocultural, and environmental contexts, it is possible for a clean fuel intervention to reduce personal PM_2.5_ exposures below the WHO-IT1 guideline value.

These exposure reductions also suggest the potential for similar exposure contrasts throughout HAPIN for other participants, including the child born during the trial and non-pregnant adult women (ages 40 - 79) participants living in the same household as the pregnant women. Air pollution exposure for non-pregnant adult women is being measured six times over the course of the study, with corresponding measures of blood pressure and collection of samples for biomarker analyses. Children resulting from the pregnancies are being measured for exposure three times over their first year of life, with additional measurements related to health (acute lower respiratory infection, anthropometry, and cognitive development) and collection of samples for biomarker analyses.

Relationships between PM exposure and health endpoints (including those measured in the HAPIN trial for adults and children) are thought to have a supralinear shape; that is, larger health gains are expected as one moves toward lower PM exposure (Burnett et al. 2014; Steenland et al. 2018), where the exposure-response relationship appears visually steeper. Should the exposure contrasts observed for pregnant women be similar for other adult women and children, this suggests promise for measuring corresponding improvements in health.

## Supporting information

SI-HAPIN-Exposure-Contrasts

## Data Availability

All data produced in the present study are available upon reasonable request to the authors after completion of the trial.

## 5. Acknowledgements

We would like to express our deep and sincere gratitude to the households that graciously invited us into their homes for this study. We also would like to thank the field teams, which worked diligently to collect the data presented here. We thank Simeon Nyandwi, Patrick Karakwende, Adolphe Ndikumana, and other colleagues at Eagle Research Centre, Ltd, Kigali, Rwanda and School of Public Health, College of Medicine and Health Sciences, University of Rwanda. We thank the field staff from the Guatemala IRC: Jose Polanco-Lemus, Miguel Arana-Marroquin, Oscar Ureta, Ramiro Mencos, Evelia Gonzalez, Angela Baldizón, Norma Aquino, Yamileth Lucero, and Miriam Tellez. We thank the field staff from the Peru IRC: Jose Polanco-Lemus, Miguel Arana-Marroquin, Oscar Ureta, Ramiro Mencos, Evelia Gonzalez, Angela Baldizón, Norma Aquino, Yamileth Lucero, Miriam Tellez.

We also wish to extend our deep gratitude to the late Kirk R. Smith, who helped shape the HAPIN study and provided his always-keen insights for the exposure assessment.

^+^HAPIN Investigators: Vigneswari Aravindalochanan, Gloriose Bankundiye, Dana Boyd Barr, Alejandra Bussalleu, Eduardo Canuz, Adly Castañaza, Yunyun Chen, Marilú Chiang, Rachel Craik, Victor G. Davila-Roman, Lisa de las Fuentes, Oscar De León, Lisa Elon, Juan Gabriel Espinoza, Sarada Garg, Sarah Hamid, Stella Hartinger, Steven A. Harvey, Mayari Hengstermann, Ian Hennessee, Phabiola M. Herrera, Shakir Hossen, Penelope P. Howards, Lindsay Jaacks, Shirin Jabbarzadeh, Pattie Lenzen, Amy E. Lovvorn, Jane Mbabazi, Eric McCollum, Rachel Meyers, Erick Mollinedo, Lawrence Moulton, Alexie Mukeshimana, Bernard Mutariyani, Durairaj Natesan, Azhar Nizam, Jean de Dieu Ntivuguruzwa, Aris Papageorghiou, Naveen Puttaswamy, Elisa Puzzolo, Ashlinn Quinn, Karthikeyan Dharmapuri Rajamani, Usha Ramakrishnan, Rengaraj Ramasami, Alexander Ramirez, P. Barry Ryan, Sudhakar Saidam, Jeremy A. Sarnat, Suzanne Simkovich, Sheela S.Sinharoy, Kirk R. Smith, Damien Swearing, Gurusamy Thangavel, Lisa M. Thompson, Ashley Toenjes, Viviane Valdes, Kendra N. Williams, Wenlu Ye, Bonnie N. Young

This study was funded by the U.S. National Institutes of Health [cooperative agreement 1UM1HL134590] in collaboration with the Bill & Melinda Gates Foundation [OPP1131279]. Under the grant conditions of the Foundation, a Creative Commons Attribution 4.0 Generic License has already been assigned to the Author Accepted Manuscript version that might arise from this submission. A multidisciplinary, independent Data and Safety Monitoring Board (DSMB) appointed by the National Heart, Lung, and Blood Institute (NHLBI) monitors the quality of the data and protects the safety of patients enrolled in the HAPIN trial. NHLBI DSMB: Nancy R Cook, Stephen Hecht, Catherine Karr (Chair), Joseph Millum, Nalini Sathiakumar, Paul K Whelton, Gail G Weinmann (Executive Secretary). Program Coordination: Gail Rodgers, Bill & Melinda Gates Foundation; Claudia L Thompson, National Institute of Environmental Health Science; Mark J. Parascandola, National Cancer Institute; Marion Koso-Thomas, Eunice Kennedy Shriver National Institute of Child Health and Human Development; Joshua P Rosenthal, Fogarty International Center; Conception R Nierras, NIH Office of Strategic Coordination Common Fund; Katherine Kavounis, Dong-Yun Kim, Antonello Punturieri, and Barry S Schmetter, NHLBI. The findings and conclusions in this report are those of the authors and do not necessarily represent the official position of the US National Institutes of Health or Department of Health and Human Services.

The findings and conclusions in this report are those of the authors and do not necessarily represent the official position of the U.S. National Institutes of Health or Department of Health and Human Services or the Bill and Melinda Gates Foundation.

