## Supplementary material for "Exposure contrasts of pregnant women during the Household Air Pollution Intervention Network randomized controlled trial": SI-HAPIN-Exposure-Contrasts

*13 Tables and 12 Figures*

**Table of Contents**

| | |
|---|---|
| Study site information | 2 |
| Baseline household and maternal characteristics in HAPIN by IRC and intervention arm (Table S1) | 3-5 |
| Additional household and exposure-related characteristics (Table S2) | 6-14 |
| Additional Methods and QA/QC: PM2.5 Sampling | 15-17 |
| Gravimetric sample validity (Table S2) | 15 |
| ECM gravimetric data analysis | 15 |
| ECM duplicates (Figure S1, Table S3) | 16 |
| Processing and use of nephelometric data when gravimetric data is absent | 17 |
| Compliance (Figure S2, Table S4) | 18 |
| Additional Methods and QA/QC: Black Carbon | 19-20 |
| BC duplicate measures (Figure S3) | 19 |
| BC duplicate performance and fit metrics (Table S5) | 20 |
| Additional Methods and QA/QC: CO | 21-22 |
| CO sample validity (Table S6) | 21 |
| CO duplicate performance fit metrics (Table S7) | 21 |
| CO duplicate measures (Figure S4) | 22 |
| IRC and pollutant-specific findings (Figure S5) | 23-24 |
| Relationships between pollutants (Figures S6-S7) | 25-26 |
| Modeling approaches (Table S8) | 27 |
| BC and CO modeling output (Figure S8, S9; Table S9, S10) | 28-29 |
| IRC-specific model output (Figures S10, S11, S12; Tables S11, S12, S13) | 30-39 |

### Study site information

Within each IRC, specific study sites were chosen during a 12-month period of formative research. Numerous factors drove site selection, including population density, fuel use, household characteristics, socioeconomic status, and other potential sources of ambient and/or household air pollution which may impact the potential to understand and describe exposure reductions potentially attributable to the HAPIN intervention package.

In India, sites were chosen in Villupuram and Nagapattinam districts in the state of Tamil Nadu, where traditional mud and clay stoves fueled with wood were used predominantly indoors. In Guatemala, participants were recruited in Jalapa municipality, where indoor wood fuel use in chimney stoves and open fires was common. In Peru, activities occurred in the Department of Puno, where households burned wood and dung in built-in open or chimney stoves for cooking. In Rwanda, households that use three-stone fires or simple open stoves (ronderezas) – predominantly indoors with wood or portable charcoal-burning stoves (imbabura) – were recruited from Kayonza District in the Eastern Province.

**Table S1** Baseline household and maternal characteristics in HAPIN by IRC and intervention arm

| | Guatemala | | | | India | | | | Peru | | | | Rwanda | | | |
|---|---|---|---|---|---|---|---|---|---|---|---|---|---|---|---|---|
| Variable | Control (N=400) | | Intervention (N=400) | | Control (N=399) | | Intervention (N=400) | | Control (N=402) | | Intervention (N=396) | | Control (N=404) | | Intervention (N=394) | |
| **Household characteristics** | | | | | | | | | | | | | | | | |
| **Household size** | | | | | | | | | | | | | | | | |
| Mean (SD) | 5.1 | (2.6) | 5.3 | (2.7) | 3.8 | (1.5) | 3.7 | (1.6) | 4.7 | (1.8) | 4.5 | (1.7) | 3.5 | (1.5) | 3.5 | (1.5) |
| [Range] | | [2-18] | | [2-17] | | [2-9] | | [1-10] | | [2-12] | | [2-11] | | [1-10] | | [1-10] |
| Missing | | 0 | | 0 | | 0 | | 0 | | 0 | | 1 | | 0 | | 0 |
| **Floor type in main home[a]** | | | | | | | | | | | | | | | | |
| Concrete | 106 | ( 27%) | 102 | ( 26%) | 235 | ( 59%) | 216 | ( 54%) | 190 | ( 47%) | 191 | ( 48%) | 112 | ( 28%) | 164 | ( 42%) |
| Mud | 361 | ( 90%) | 362 | ( 91%) | 138 | ( 35%) | 147 | ( 37%) | 266 | ( 66%) | 279 | ( 70%) | 297 | ( 74%) | 229 | ( 58%) |
| Other | 18 | ( 5%) | 24 | ( 6%) | 31 | ( 8%) | 44 | ( 11%) | 45 | ( 11%) | 36 | ( 9%) | 6 | ( 1%) | 10 | ( 3%) |
| Missing | 1 | | 0 | | 0 | | 0 | | 0 | | 0 | | 0 | | 0 | |
| **Household wealth at national quintiles** | | | | | | | | | | | | | | | | |
| Lowest | 235 | ( 59%) | 236 | ( 59%) | 82 | ( 21%) | 97 | ( 24%) | 210 | ( 52%) | 202 | ( 51%) | 21 | ( 5%) | 11 | ( 3%) |
| Second lowest | 103 | ( 26%) | 102 | ( 26%) | 206 | ( 52%) | 196 | ( 49%) | 117 | ( 29%) | 103 | ( 26%) | 67 | ( 17%) | 57 | ( 14%) |
| Medium | 55 | ( 14%) | 47 | ( 12%) | 90 | ( 23%) | 85 | ( 21%) | 62 | ( 15%) | 85 | ( 21%) | 117 | ( 29%) | 86 | ( 22%) |
| Second highest | 7 | ( 2%) | 15 | ( 4%) | 21 | ( 5%) | 22 | ( 6%) | 13 | ( 3%) | 6 | ( 2%) | 149 | ( 37%) | 142 | ( 36%) |
| Highest | 0 | ( 0%) | 0 | ( 0%) | 0 | ( 0%) | 0 | ( 0%) | 0 | ( 0%) | 0 | ( 0%) | 50 | ( 12%) | 98 | ( 25%) |
| **Access to electricity** | | | | | | | | | | | | | | | | |
| No | 40 | ( 10%) | 46 | ( 12%) | 16 | ( 4%) | 12 | ( 3%) | 22 | ( 5%) | 24 | ( 6%) | 272 | ( 72%) | 213 | ( 58%) |
| Yes | 360 | ( 90%) | 354 | ( 89%) | 383 | ( 96%) | 388 | ( 97%) | 380 | ( 95%) | 372 | ( 94%) | 104 | ( 28%) | 156 | ( 42%) |
| Missing | 0 | | 0 | | 0 | | 0 | | 0 | | 0 | | 28 | | 25 | |
| **Maternal characteristics** | | | | | | | | | | | | | | | | |
| **Age (year)** | | | | | | | | | | | | | | | | |
| Mean (SD) | 25.0 | (4.5) | 24.5 | (4.4) | 23.9 | (3.9) | 24.0 | (3.7) | 25.4 | (4.6) | 25.6 | (4.3) | 27.3 | (4.5) | 27.3 | (4.3) |
| [Range] | | [18-35] | | [18-35] | | [18-35] | | [18-35] | | [18-35] | | [18-35] | | [18-35] | | [18-35] |
| Missing | | 0 | | 0 | | 0 | | 0 | | 0 | | 0 | | 0 | | 0 |
| **Mother: highest level of education completed** | | | | | | | | | | | | | | | | |
| No formal education or Primary school incomplete | 192 | ( 48%) | 189 | ( 47%) | 155 | ( 39%) | 130 | ( 33%) | 20 | ( 5%) | 15 | ( 4%) | 191 | ( 47%) | 147 | ( 37%) |
| Primary school complete or Secondary school incomplete | 152 | ( 38%) | 160 | ( 40%) | 111 | ( 28%) | 116 | ( 29%) | 103 | ( 26%) | 131 | ( 33%) | 167 | ( 41%) | 151 | ( 38%) |
| Secondary school complete or Vocational or Some college or university | 56 | ( 14%) | 51 | ( 13%) | 133 | ( 33%) | 154 | ( 39%) | 279 | ( 69%) | 249 | ( 63%) | 46 | ( 11%) | 96 | ( 24%) |
| Missing | 0 | | 0 | | 0 | | 0 | | 0 | | 1 | | 0 | | 0 | |
| **Mother's occupation[b]** | | | | | | | | | | | | | | | | |
| Household | 377 | ( 94%) | 369 | ( 92%) | 213 | ( 53%) | 219 | ( 55%) | 67 | ( 17%) | 65 | ( 16%) | 22 | ( 5%) | 37 | ( 9%) |
| Agriculture | 0 | ( 0%) | 6 | ( 2%) | 170 | ( 43%) | 168 | ( 42%) | 303 | ( 75%) | 298 | ( 75%) | 322 | ( 80%) | 269 | ( 68%) |
| Commercial | 6 | ( 2%) | 11 | ( 3%) | 3 | ( 1%) | 1 | ( <1%) | 60 | ( 15%) | 51 | ( 13%) | 62 | ( 15%) | 74 | ( 19%) |
| Other or N/A | 16 | ( 4%) | 13 | ( 3%) | 13 | ( 3%) | 12 | ( 3%) | 22 | ( 5%) | 20 | ( 5%) | 25 | ( 6%) | 37 | ( 9%) |
| Missing | 1 | | 1 | | 0 | | 0 | | 0 | | 0 | | 0 | | 0 | |

| Variable | Guatemala Control (N=400) | | Guatemala Intervention (N=400) | | India Control (N=399) | | India Intervention (N=400) | | Peru Control (N=402) | | Peru Intervention (N=396) | | Rwanda Control (N=404) | | Rwanda Intervention (N=394) | |
|---|---|---|---|---|---|---|---|---|---|---|---|---|---|---|---|---|
| **Exposure visit observations** | | | | | | | | | | | | | | | | |
| **Primary cooking area roof** | | | | | | | | | | | | | | | | |
| No | 1 | ( <1%) | 1 | ( <1%) | 1 | ( <1%) | 3 | ( 1%) | 88 | ( 22%) | 91 | ( 23%) | 124 | ( 31%) | 119 | ( 30%) |
| Yes | 395 | (100%) | 398 | (100%) | 398 | (100%) | 397 | ( 99%) | 312 | ( 78%) | 305 | ( 77%) | 279 | ( 69%) | 274 | ( 70%) |
| Missing | 4 | | 1 | | 0 | | 0 | | 2 | | 0 | | 1 | | 1 | |
| **Primary cooking area walls (#)** | | | | | | | | | | | | | | | | |
| None | 3 | ( 1%) | 4 | ( 1%) | 11 | ( 3%) | 5 | ( 1%) | 2 | ( 1%) | 1 | ( <1%) | 4 | ( 1%) | 2 | ( 1%) |
| One to three | 33 | ( 8%) | 39 | ( 10%) | 77 | ( 19%) | 66 | ( 17%) | 33 | ( 11%) | 21 | ( 7%) | 5 | ( 2%) | 6 | ( 2%) |
| Four or more | 359 | ( 91%) | 355 | ( 89%) | 309 | ( 78%) | 326 | ( 82%) | 277 | ( 89%) | 283 | ( 93%) | 269 | ( 97%) | 266 | ( 97%) |
| **Kitchen volume ($m^3$)[c]** | | | | | | | | | | | | | | | | |
| Mean (SD) | 33.4 | (16.4) | 34.0 | (17.2) | 20.1 | (13.3) | 21.9 | (15.9) | 19.9 | (12.5) | 18.8 | (11.7) | 12.3 | (7.6) | 13.3 | (7.0) |
| [Range] | | [7-100] | | [7-126] | | [0-86] | | [2-133] | | [2-116] | | [2-82] | | [0-83] | | [0-53] |
| N | | 378 | | 379 | | 386 | | 391 | | 296 | | 290 | | 272 | | 270 |
| Missing | | 14 | | 15 | | 0 | | 1 | | 14 | | 14 | | 2 | | 2 |
| **Baseline Exposure Questionnaire** | | | | | | | | | | | | | | | | |
| **Primary cook** | | | | | | | | | | | | | | | | |
| Myself | 337 | ( 84%) | 339 | ( 85%) | 374 | ( 94%) | 383 | ( 96%) | 221 | ( 55%) | 234 | ( 59%) | 386 | ( 96%) | 351 | ( 89%) |
| Mother/Mother-in-law | 56 | ( 14%) | 57 | ( 14%) | 21 | ( 5%) | 14 | ( 4%) | 173 | ( 43%) | 158 | ( 40%) | 4 | ( 1%) | 1 | ( <1%) |
| Sister/Sister-in-law | 6 | ( 2%) | 1 | ( <1%) | 3 | ( 1%) | 1 | ( <1%) | 1 | ( <1%) | 3 | ( 1%) | 5 | ( 1%) | 9 | ( 2%) |
| Daughter | 0 | ( 0%) | 2 | ( 1%) | 0 | ( 0%) | 1 | ( <1%) | 1 | ( <1%) | 0 | ( 0%) | 2 | ( <1%) | 6 | ( 2%) |
| Hired cook in the home | 0 | ( 0%) | 0 | ( 0%) | 0 | ( 0%) | 0 | ( 0%) | 0 | ( 0%) | 0 | ( 0%) | 2 | ( <1%) | 23 | ( 6%) |
| Other | 1 | ( <1%) | 0 | ( 0%) | 1 | ( <1%) | 1 | ( <1%) | 5 | ( 1%) | 1 | ( <1%) | 4 | ( 1%) | 3 | ( 1%) |
| Missing | 0 | | 1 | | 0 | | 0 | | 1 | | 0 | | 1 | | 1 | |
| **Cook Times/day** | | | | | | | | | | | | | | | | |
| One or two | 11 | ( 3%) | 7 | ( 2%) | 339 | ( 85%) | 343 | ( 86%) | 164 | ( 41%) | 181 | ( 46%) | 357 | ( 89%) | 322 | ( 82%) |
| Three or more | 389 | ( 97%) | 393 | ( 98%) | 60 | ( 15%) | 57 | ( 14%) | 237 | ( 59%) | 215 | ( 54%) | 46 | ( 11%) | 71 | ( 18%) |
| Missing | 0 | | 0 | | 0 | | 0 | | 1 | | 0 | | 1 | | 1 | |
| **Cook times/week[d]** | | | | | | | | | | | | | | | | |
| One to Seven | 20 | ( 5%) | 12 | ( 3%) | 53 | ( 13%) | 66 | ( 17%) | 102 | ( 26%) | 89 | ( 22%) | 56 | ( 14%) | 90 | ( 23%) |
| Eight to Fourteen | 33 | ( 8%) | 20 | ( 5%) | 301 | ( 75%) | 287 | ( 72%) | 150 | ( 38%) | 154 | ( 39%) | 308 | ( 76%) | 259 | ( 66%) |
| Fifteen or more | 347 | ( 87%) | 368 | ( 92%) | 45 | ( 11%) | 47 | ( 12%) | 148 | ( 37%) | 153 | ( 39%) | 39 | ( 10%) | 44 | ( 11%) |
| Missing | 0 | | 0 | | 0 | | 0 | | 2 | | 0 | | 1 | | 1 | |
| **# of Stoves** | | | | | | | | | | | | | | | | |
| None | 0 | ( 0%) | 0 | ( 0%) | 0 | ( 0%) | 0 | ( 0%) | 0 | ( 0%) | 1 | ( <1%) | 1 | ( <1%) | 0 | ( 0%) |
| One | 138 | ( 35%) | 135 | ( 34%) | 271 | ( 68%) | 282 | ( 71%) | 105 | ( 26%) | 90 | ( 23%) | 248 | ( 62%) | 228 | ( 58%) |
| Two or more | 262 | ( 66%) | 265 | ( 66%) | 128 | ( 32%) | 118 | ( 30%) | 295 | ( 74%) | 304 | ( 77%) | 154 | ( 38%) | 165 | ( 42%) |
| Missing | 0 | | 0 | | 0 | | 0 | | 2 | | 1 | | 1 | | 1 | |
| **Primary Stove: chimney** | | | | | | | | | | | | | | | | |
| No | 310 | ( 78%) | 313 | ( 78%) | 397 | ( 99%) | 398 | (100%) | 249 | ( 62%) | 252 | ( 64%) | 382 | ( 95%) | 379 | ( 96%) |
| Yes | 90 | ( 23%) | 87 | ( 22%) | 2 | ( 1%) | 2 | ( 1%) | 152 | ( 38%) | 144 | ( 36%) | 21 | ( 5%) | 14 | ( 4%) |
| Missing | 0 | | 0 | | 0 | | 0 | | 1 | | 0 | | 1 | | 1 | |

| Variable | Guatemala Control (N=400) | | Guatemala Intervention (N=400) | | India Control (N=399) | | India Intervention (N=400) | | Peru Control (N=402) | | Peru Intervention (N=396) | | Rwanda Control (N=404) | | Rwanda Intervention (N=394) | |
|---|---|---|---|---|---|---|---|---|---|---|---|---|---|---|---|---|
| **Baseline Exposure** | | | | | | | | | | | | | | | | |
| **Primary Fuel type** | | | | | | | | | | | | | | | | |
| Cow dung | 0 | ( 0%) | 0 | ( 0%) | 0 | ( 0%) | 0 | ( 0%) | 351 | ( 88%) | 346 | ( 87%) | 0 | ( 0%) | 0 | ( 0%) |
| Wood | 394 | ( 99%) | 399 | (100%) | 399 | (100%) | 400 | (100%) | 48 | ( 12%) | 42 | ( 11%) | 323 | ( 80%) | 257 | ( 65%) |
| Charcoal | 0 | ( 0%) | 0 | ( 0%) | 0 | ( 0%) | 0 | ( 0%) | 0 | ( 0%) | 0 | ( 0%) | 72 | ( 18%) | 125 | ( 32%) |
| Other | 2 | ( 1%) | 1 | ( <1%) | 0 | ( 0%) | 0 | ( 0%) | 2 | ( <1%) | 8 | ( 2%) | 8 | ( 2%) | 11 | ( 3%) |
| Missing | 4 | | 0 | | 0 | | 0 | | 1 | | 0 | | 1 | | 1 | |
| **Primary lighting source** | | | | | | | | | | | | | | | | |
| Torch (battery) | 5 | ( 1%) | 5 | ( 1%) | 4 | ( 1%) | 7 | ( 2%) | 4 | ( 1%) | 8 | ( 2%) | 90 | ( 22%) | 83 | ( 21%) |
| Kerosene lamp | 1 | ( <1%) | 5 | ( 1%) | 9 | ( 2%) | 7 | ( 2%) | 1 | ( <1%) | 0 | ( 0%) | 37 | ( 9%) | 22 | ( 6%) |
| Solar light | 0 | ( 0%) | 2 | ( 1%) | 2 | ( 1%) | 1 | ( <1%) | 12 | ( 3%) | 11 | ( 3%) | 138 | ( 34%) | 119 | ( 30%) |
| Electricity | 361 | ( 90%) | 348 | ( 87%) | 384 | ( 96%) | 385 | ( 96%) | 371 | ( 93%) | 359 | ( 91%) | 88 | ( 22%) | 138 | ( 35%) |
| Other | 33 | ( 8%) | 40 | ( 10%) | 0 | ( 0%) | 0 | ( 0%) | 13 | ( 3%) | 18 | ( 5%) | 50 | ( 12%) | 31 | ( 8%) |
| Missing | 0 | | 0 | | 0 | | 0 | | 1 | | 0 | | 1 | | 1 | |

[a]Multiple floor materials may be reported for the same household, so households may appear more than once
[b]Among multiparous women
[c]Kitchen with a dimension greater than 25 meters or less than 0.5 meter was considered unreasonable and a data entry error
[d]Calculated as multiplying [cook times/day] with [cook days/week]

**Table S2** Additional household and exposure-related characteristics

| Variable | Guatemala Control (N=400) | Guatemala Intervention (N=400) | India Control (N=399) | India Intervention (N=400) | Peru Control (N=402) | Peru Intervention (N=396) | Rwanda Control (N=404) | Rwanda Intervention (N=394) |
|---|---|---|---|---|---|---|---|---|
| **Exposure Equipment** | | | | | | | | |
| **Primary cooking area: roof** | | | | | | | | |
| No | 1 ( <1%) | 1 ( <1%) | 1 ( <1%) | 3 ( 1%) | 88 ( 22%) | 91 ( 23%) | 124 ( 31%) | 119 ( 30%) |
| Yes | 395 (100%) | 398 (100%) | 398 (100%) | 397 ( 99%) | 312 ( 78%) | 305 ( 77%) | 279 ( 69%) | 274 ( 70%) |
| Missing | 4 | 1 | 0 | 0 | 2 | 0 | 1 | 1 |
| **Primary cooking area: walls#** | | | | | | | | |
| None | 3 ( 1%) | 4 ( 1%) | 11 ( 3%) | 5 ( 1%) | 2 ( 1%) | 1 ( <1%) | 4 ( 1%) | 2 ( 1%) |
| One to three | 33 ( 8%) | 39 ( 10%) | 77 ( 19%) | 66 ( 17%) | 33 ( 11%) | 21 ( 7%) | 5 ( 2%) | 6 ( 2%) |
| Four or more | 359 ( 91%) | 355 ( 89%) | 309 ( 78%) | 326 ( 82%) | 277 ( 89%) | 283 ( 93%) | 269 ( 97%) | 266 ( 97%) |
| Missing | 0 | 0 | 1 | 0 | 0 | 0 | 1 | 0 |
| **Primary material: kitchen wall** | | | | | | | | |
| Impermeable, like brick/cement/stone/wood/corrugated metal | 344 ( 88%) | 335 ( 85%) | 230 ( 60%) | 232 ( 59%) | 266 ( 86%) | 271 ( 89%) | 262 ( 96%) | 264 ( 98%) |
| Permeable, like reed/thatch/mesh/wattle | 48 ( 12%) | 59 ( 15%) | 156 ( 40%) | 160 ( 41%) | 44 ( 14%) | 33 ( 11%) | 10 ( 4%) | 6 ( 2%) |
| Missing | 0 | 0 | 0 | 0 | 0 | 0 | 2 | 2 |
| **Primary material: kitchen roof** | | | | | | | | |
| Impermeable, like brick/cement/stone/wood/corrugated metal | 375 ( 95%) | 379 ( 95%) | 201 ( 51%) | 211 ( 53%) | 209 ( 67%) | 217 ( 71%) | 273 ( 98%) | 269 ( 98%) |
| Permeable, like reed/thatch/mesh/wattle | 20 ( 5%) | 19 ( 5%) | 197 ( 49%) | 186 ( 47%) | 103 ( 33%) | 88 ( 29%) | 5 ( 2%) | 5 ( 2%) |
| Missing | 0 | 0 | 0 | 0 | 0 | 0 | 1 | 0 |
| **Door/entrance: # open during stove use** | | | | | | | | |
| None | 6 ( 2%) | 8 ( 2%) | 0 ( 0%) | 0 ( 0%) | 13 ( 4%) | 21 ( 7%) | 4 ( 1%) | 2 ( 1%) |
| One | 290 ( 74%) | 287 ( 72%) | 191 ( 48%) | 190 ( 48%) | 280 ( 90%) | 264 ( 87%) | 263 ( 95%) | 264 ( 96%) |
| Two or more | 98 ( 25%) | 103 ( 26%) | 207 ( 52%) | 207 ( 52%) | 19 ( 6%) | 19 ( 6%) | 11 ( 4%) | 8 ( 3%) |
| Missing | 1 | 0 | 0 | 0 | 0 | 1 | 1 | 0 |
| **Window/eve/unfinished wall: # open during stove** | | | | | | | | |
| None | 25 ( 6%) | 28 ( 7%) | 14 ( 4%) | 10 ( 3%) | 120 ( 38%) | 118 ( 39%) | 109 ( 39%) | 95 ( 35%) |
| One or two | 21 ( 5%) | 18 ( 5%) | 66 ( 17%) | 63 ( 16%) | 131 ( 42%) | 136 ( 45%) | 109 ( 39%) | 134 ( 49%) |
| Three or more | 348 ( 88%) | 352 ( 88%) | 318 ( 80%) | 324 ( 82%) | 61 ( 20%) | 50 ( 16%) | 60 ( 22%) | 45 ( 16%) |
| Missing | 1 | 0 | 0 | 0 | 0 | 1 | 1 | 0 |
| **Window/opening above stove** | | | | | | | | |
| No | 25 ( 6%) | 23 ( 6%) | 291 ( 73%) | 299 ( 75%) | 112 ( 36%) | 144 ( 48%) | 148 ( 53%) | 145 ( 53%) |
| Yes | 370 ( 94%) | 375 ( 94%) | 106 ( 27%) | 98 ( 25%) | 200 ( 64%) | 159 ( 52%) | 130 ( 47%) | 129 ( 47%) |
| Missing | 0 | 0 | 1 | 0 | 0 | 2 | 1 | 0 |
| **Kitchen volume ($m^3$)[a]** | | | | | | | | |
| Mean (SD) | 33.4 (16.4) | 34.0 (17.2) | 20.1 (13.3) | 21.9 (15.9) | 19.9 (12.5) | 18.8 (11.7) | 12.3 (7.6) | 13.3 (7.0) |
| [Range] | [7-100] | [7-126] | [0-86] | [2-133] | [2-116] | [2-82] | [0-83] | [0-53] |
| N | 378 | 379 | 386 | 391 | 296 | 290 | 272 | 270 |
| Missing | 14 | 15 | 0 | 1 | 14 | 14 | 2 | 2 |

| | Guatemala | | | | India | | | | Peru | | | | Rwanda | | | |
|---|---|---|---|---|---|---|---|---|---|---|---|---|---|---|---|---|
| **Variable** | **Control (N=400)** | | **Intervention (N=400)** | | **Control (N=399)** | | **Intervention (N=400)** | | **Control (N=402)** | | **Intervention (N=396)** | | **Control (N=404)** | | **Intervention (N=394)** | |
| **Exposure Compliance** | | | | | | | | | | | | | | | | |
| **Use any cook stoves** | | | | | | | | | | | | | | | | |
| No | 4 | ( 1%) | 3 | ( 1%) | 5 | ( 1%) | 3 | ( 1%) | 6 | ( 2%) | 3 | ( 1%) | 6 | ( 1%) | 10 | ( 3%) |
| Yes | 396 | ( 99%) | 397 | ( 99%) | 394 | ( 99%) | 397 | ( 99%) | 392 | ( 98%) | 392 | ( 99%) | 398 | ( 99%) | 382 | ( 97%) |
| Missing | 0 | | 0 | | 0 | | 0 | | 4 | | 1 | | 0 | | 2 | |
| **Type of main stove** | | | | | | | | | | | | | | | | |
| Stove with chimney (Biomass, Rondereza, Comal) | 84 | ( 21%) | 77 | ( 19%) | 0 | ( 0%) | 0 | ( 0%) | 29 | ( 7%) | 32 | ( 8%) | 203 | ( 50%) | 212 | ( 54%) |
| Stove1: Open/3 stone fire, Mud/Metal Chula, pollo/plancha | 313 | ( 78%) | 319 | ( 80%) | 378 | ( 95%) | 380 | ( 95%) | 332 | ( 83%) | 329 | ( 83%) | 173 | ( 43%) | 151 | ( 39%) |
| Stove1: Portable | 1 | ( <1%) | 2 | ( 1%) | 16 | ( 4%) | 16 | ( 4%) | 0 | ( 0%) | 0 | ( 0%) | 24 | ( 6%) | 23 | ( 6%) |
| Stove1: Kerosene | 0 | ( 0%) | 1 | ( <1%) | 0 | ( 0%) | 0 | ( 0%) | 0 | ( 0%) | 0 | ( 0%) | 2 | ( <1%) | 1 | ( <1%) |
| Stove1: LPG | 1 | ( <1%) | 0 | ( 0%) | 0 | ( 0%) | 1 | ( <1%) | 28 | ( 7%) | 31 | ( 8%) | 1 | ( <1%) | 1 | ( <1%) |
| Stove1: Type Other | 0 | ( 0%) | 0 | ( 0%) | 0 | ( 0%) | 0 | ( 0%) | 3 | ( 1%) | 0 | ( 0%) | 2 | ( <1%) | 1 | ( <1%) |
| Missing | 0 | | 0 | | 0 | | 0 | | 2 | | 0 | | 0 | | 2 | |
| **Stove1: lit hours**[b] | | | | | | | | | | | | | | | | |
| None | 1 | ( <1%) | 0 | ( 0%) | 0 | ( 0%) | 0 | ( 0%) | 1 | ( <1%) | 0 | ( 0%) | 1 | ( <1%) | 1 | ( <1%) |
| One or two | 51 | ( 13%) | 38 | ( 10%) | 132 | ( 34%) | 144 | ( 36%) | 153 | ( 39%) | 173 | ( 44%) | 158 | ( 41%) | 137 | ( 36%) |
| Three or more | 341 | ( 87%) | 357 | ( 90%) | 262 | ( 66%) | 253 | ( 64%) | 237 | ( 61%) | 216 | ( 56%) | 228 | ( 59%) | 238 | ( 63%) |
| Missing | 3 | | 2 | | 0 | | 0 | | 1 | | 3 | | 11 | | 6 | |
| **Usage of main stove**[c] | | | | | | | | | | | | | | | | |
| Stove1: Cook lunch | 373 | ( 93%) | 373 | ( 93%) | 54 | ( 14%) | 57 | ( 14%) | 241 | ( 60%) | 254 | ( 64%) | 319 | ( 79%) | 296 | ( 76%) |
| Stove1: Cook dinner | 379 | ( 95%) | 388 | ( 97%) | 352 | ( 88%) | 347 | ( 87%) | 258 | ( 65%) | 253 | ( 64%) | 357 | ( 88%) | 341 | ( 87%) |
| Stove1: Cook breakfast | 373 | ( 93%) | 378 | ( 95%) | 75 | ( 19%) | 100 | ( 25%) | 322 | ( 81%) | 312 | ( 79%) | 120 | ( 30%) | 133 | ( 34%) |
| Stove1: Reheat food | 259 | ( 65%) | 253 | ( 63%) | 35 | ( 9%) | 32 | ( 8%) | 50 | ( 13%) | 46 | ( 12%) | 20 | ( 5%) | 23 | ( 6%) |
| Stove1: Make animal food | 5 | ( 1%) | 9 | ( 2%) | 10 | ( 3%) | 6 | ( 2%) | 239 | ( 60%) | 217 | ( 55%) | 1 | ( <1%) | 0 | ( 0%) |
| Stove1: Cook beans (Rwanda and Guatemala only) | 212 | ( 53%) | 186 | ( 47%) | N/A | N/A | N/A | N/A | N/A | N/A | N/A | N/A | 58 | ( 14%) | 59 | ( 15%) |
| Stove1: Reheat beans (Rwanda and Guatemala only) | 338 | ( 85%) | 329 | ( 82%) | N/A | N/A | N/A | N/A | N/A | N/A | N/A | N/A | 15 | ( 4%) | 12 | ( 3%) |
| Stove1: Use: Other | 3 | ( 1%) | 4 | ( 1%) | 1 | ( <1%) | 1 | ( <1%) | 2 | ( 1%) | 3 | ( 1%) | 5 | ( 1%) | 6 | ( 2%) |
| Missing | 0 | | 0 | | 0 | | 0 | | 2 | | 0 | | 0 | | 2 | |
| **Stove1: location** | | | | | | | | | | | | | | | | |
| In participant's bedroom | 18 | ( 5%) | 18 | ( 5%) | 78 | ( 20%) | 88 | ( 22%) | 5 | ( 1%) | 4 | ( 1%) | 6 | ( 2%) | 2 | ( 1%) |
| Room immediately adjacent to the participant's bedroom | 194 | ( 49%) | 186 | ( 47%) | 143 | ( 36%) | 122 | ( 31%) | 31 | ( 8%) | 32 | ( 8%) | 9 | ( 2%) | 13 | ( 3%) |
| Separated from the participant's bedroom but inside the house | 103 | ( 26%) | 122 | ( 31%) | 75 | ( 19%) | 73 | ( 18%) | 72 | ( 18%) | 74 | ( 19%) | 24 | ( 6%) | 19 | ( 5%) |
| Outside the house (outdoors) | 4 | ( 1%) | 7 | ( 2%) | 1 | ( <1%) | 4 | ( 1%) | 86 | ( 22%) | 79 | ( 20%) | 125 | ( 31%) | 130 | ( 34%) |
| In a separate building detached from the bedroom (main house) | 77 | ( 19%) | 64 | ( 16%) | 97 | ( 25%) | 109 | ( 27%) | 197 | ( 50%) | 202 | ( 52%) | 234 | ( 59%) | 217 | ( 57%) |
| Other | 0 | ( 0%) | 0 | ( 0%) | 0 | ( 0%) | 1 | ( <1%) | 1 | ( <1%) | 0 | ( 0%) | 0 | ( 0%) | 0 | ( 0%) |
| Missing | 0 | | 0 | | 0 | | 0 | | 0 | | 1 | | 0 | | 1 | |

| | Guatemala | | | | India | | | | Peru | | | | Rwanda | | |
|---|---|---|---|---|---|---|---|---|---|---|---|---|---|---|---|
| **Variable** | **Control (N=400)** | | **Intervention (N=400)** | | **Control (N=399)** | | **Intervention (N=400)** | | **Control (N=402)** | | **Intervention (N=396)** | | **Control (N=404)** | | **Intervention (N=394)** |
| **Exposure Compliance** | | | | | | | | | | | | | | | |

| | | | | | | | | | | | | | | | | |
|---|---|---|---|---|---|---|---|---|---|---|---|---|---|---|---|---|
| **Persons who mainly used the cookstove?[d]** | | | | | | | | | | | | | | | | |
| Stove1: Myself | 392 | ( 98%) | 387 | ( 97%) | 379 | ( 95%) | 388 | ( 97%) | 332 | ( 83%) | 344 | ( 87%) | 384 | ( 95%) | 345 | ( 88%) |
| Stove1: Mother/Mother-in-law | 109 | ( 27%) | 118 | ( 30%) | 51 | ( 13%) | 50 | ( 13%) | 168 | ( 42%) | 153 | ( 39%) | 12 | ( 3%) | 8 | ( 2%) |
| Stove1: Sister/Sister-in-law | 44 | ( 11%) | 58 | ( 15%) | 4 | ( 1%) | 6 | ( 2%) | 19 | ( 5%) | 18 | ( 5%) | 10 | ( 2%) | 20 | ( 5%) |
| Stove1: Daughter | 9 | ( 2%) | 9 | ( 2%) | 0 | ( 0%) | 0 | ( 0%) | 3 | ( 1%) | 6 | ( 2%) | 12 | ( 3%) | 18 | ( 5%) |
| Stove1: Hired cook | 0 | ( 0%) | 0 | ( 0%) | 1 | ( <1%) | 0 | ( 0%) | 0 | ( 0%) | 0 | ( 0%) | 6 | ( 1%) | 25 | ( 6%) |
| Stove1: Other used | 3 | ( 1%) | 9 | ( 2%) | 2 | ( 1%) | 1 | ( <1%) | 9 | ( 2%) | 5 | ( 1%) | 14 | ( 3%) | 18 | ( 5%) |
| Missing | 0 | | 0 | | 0 | | 0 | | 2 | | 0 | | 0 | | 2 | |
| **Any stoves currently lit?[e]** | | | | | | | | | | | | | | | | |
| Currently lit: Open fire, Chula, pollo/plancha | 231 | ( 58%) | 240 | ( 60%) | 83 | ( 21%) | 99 | ( 25%) | 93 | ( 23%) | 81 | ( 20%) | 32 | ( 8%) | 29 | ( 7%) |
| Currently lit: Biomass stove/Plancha with Chimney, Imbabura | 69 | ( 17%) | 57 | ( 14%) | 0 | ( 0%) | 0 | ( 0%) | 9 | ( 2%) | 14 | ( 4%) | 22 | ( 5%) | 32 | ( 8%) |
| Currently lit: Rondereza | 0 | ( 0%) | 0 | ( 0%) | 0 | ( 0%) | 0 | ( 0%) | 0 | ( 0%) | 0 | ( 0%) | 36 | ( 9%) | 20 | ( 5%) |
| Currently lit: Portable | 1 | ( <1%) | 2 | ( 1%) | 1 | ( <1%) | 6 | ( 2%) | 0 | ( 0%) | 0 | ( 0%) | 9 | ( 2%) | 8 | ( 2%) |
| Currently lit: LPG | 0 | ( 0%) | 0 | ( 0%) | 0 | ( 0%) | 1 | ( <1%) | 12 | ( 3%) | 16 | ( 4%) | 0 | ( 0%) | 0 | ( 0%) |
| Currently lit: Electric | 0 | ( 0%) | 1 | ( <1%) | 0 | ( 0%) | 0 | ( 0%) | 0 | ( 0%) | 0 | ( 0%) | 0 | ( 0%) | 0 | ( 0%) |
| Currently lit: Comal | 3 | ( 1%) | 2 | ( 1%) | 0 | ( 0%) | 0 | ( 0%) | 1 | ( <1%) | 0 | ( 0%) | 0 | ( 0%) | 0 | ( 0%) |
| Currently lit: Other stove | 0 | ( 0%) | 0 | ( 0%) | 0 | ( 0%) | 0 | ( 0%) | 2 | ( 1%) | 2 | ( 1%) | 0 | ( 0%) | 0 | ( 0%) |
| Missing | 0 | | 0 | | 0 | | 0 | | 2 | | 0 | | 0 | | 2 | |
| **Sources of smoke inside the house[f]** | | | | | | | | | | | | | | | | |
| Inside: Combustion powered corn/flour mill | 0 | ( 0%) | 2 | ( 1%) | 0 | ( 0%) | 0 | ( 0%) | 0 | ( 0%) | 1 | ( <1%) | 0 | ( 0%) | 0 | ( 0%) |
| Inside: Other kitchen | 5 | ( 1%) | 7 | ( 2%) | 0 | ( 0%) | 3 | ( 1%) | 4 | ( 1%) | 3 | ( 1%) | 10 | ( 2%) | 5 | ( 1%) |
| Inside: Outside of home | 0 | ( 0%) | 0 | ( 0%) | 0 | ( 0%) | 0 | ( 0%) | 0 | ( 0%) | 0 | ( 0%) | 0 | ( 0%) | 1 | ( <1%) |
| Inside: Trash burning | 0 | ( 0%) | 0 | ( 0%) | 0 | ( 0%) | 0 | ( 0%) | 3 | ( 1%) | 3 | ( 1%) | 2 | ( <1%) | 1 | ( <1%) |
| Inside: Tobacco smoking | 0 | ( 0%) | 0 | ( 0%) | 0 | ( 0%) | 0 | ( 0%) | 0 | ( 0%) | 0 | ( 0%) | 1 | ( <1%) | 0 | ( 0%) |
| Inside: Burning of agricultural waste | 0 | ( 0%) | 0 | ( 0%) | 0 | ( 0%) | 0 | ( 0%) | 0 | ( 0%) | 0 | ( 0%) | 1 | ( <1%) | 0 | ( 0%) |
| Inside: Burning from preparing fields | 0 | ( 0%) | 0 | ( 0%) | 0 | ( 0%) | 0 | ( 0%) | 0 | ( 0%) | 1 | ( <1%) | 0 | ( 0%) | 0 | ( 0%) |
| Inside: Incense | 0 | ( 0%) | 0 | ( 0%) | 0 | ( 0%) | 0 | ( 0%) | 0 | ( 0%) | 0 | ( 0%) | 1 | ( <1%) | 0 | ( 0%) |
| Inside: mosquito coils | 0 | ( 0%) | 1 | ( <1%) | 0 | ( 0%) | 0 | ( 0%) | 0 | ( 0%) | 1 | ( <1%) | 0 | ( 0%) | 0 | ( 0%) |
| Inside: Intense vehicular emissions | 0 | ( 0%) | 0 | ( 0%) | 0 | ( 0%) | 0 | ( 0%) | 2 | ( 1%) | 1 | ( <1%) | 0 | ( 0%) | 0 | ( 0%) |
| Inside: Smoke source: Other | 0 | ( 0%) | 0 | ( 0%) | 0 | ( 0%) | 0 | ( 0%) | 0 | ( 0%) | 0 | ( 0%) | 1 | ( <1%) | 0 | ( 0%) |
| Missing | 0 | | 0 | | 0 | | 0 | | 2 | | 0 | | 0 | | 2 | |
| **Outdoor sources of smoke[f]** | | | | | | | | | | | | | | | | |
| Outdoor: Combustion powered corn/flour mill | 1 | ( <1%) | 1 | ( <1%) | 0 | ( 0%) | 0 | ( 0%) | 0 | ( 0%) | 0 | ( 0%) | 2 | ( <1%) | 0 | ( 0%) |
| Outdoor: Other kitchen | 39 | ( 10%) | 45 | ( 11%) | 1 | ( <1%) | 2 | ( 1%) | 2 | ( 1%) | 0 | ( 0%) | 45 | ( 11%) | 47 | ( 12%) |
| Outdoor: Outside of home | 0 | ( 0%) | 0 | ( 0%) | 0 | ( 0%) | 0 | ( 0%) | 0 | ( 0%) | 0 | ( 0%) | 2 | ( <1%) | 3 | ( 1%) |
| Outdoor: Trash burning | 2 | ( 1%) | 2 | ( 1%) | 1 | ( <1%) | 0 | ( 0%) | 1 | ( <1%) | 3 | ( 1%) | 2 | ( <1%) | 1 | ( <1%) |
| Outdoor: Tobacco smoking | 0 | ( 0%) | 0 | ( 0%) | 0 | ( 0%) | 2 | ( 1%) | 0 | ( 0%) | 0 | ( 0%) | 1 | ( <1%) | 0 | ( 0%) |
| Outdoor: Burning of agricultural waste | 0 | ( 0%) | 0 | ( 0%) | 0 | ( 0%) | 0 | ( 0%) | 0 | ( 0%) | 1 | ( <1%) | 0 | ( 0%) | 1 | ( <1%) |
| Outdoor: Burning from preparing fields | 1 | ( <1%) | 0 | ( 0%) | 0 | ( 0%) | 0 | ( 0%) | 0 | ( 0%) | 1 | ( <1%) | 0 | ( 0%) | 0 | ( 0%) |
| Outdoor: Generator | 0 | ( 0%) | 0 | ( 0%) | 0 | ( 0%) | 0 | ( 0%) | 0 | ( 0%) | 1 | ( <1%) | 0 | ( 0%) | 0 | ( 0%) |
| Outdoor: Intense vehicular emissions | 0 | ( 0%) | 2 | ( 1%) | 0 | ( 0%) | 0 | ( 0%) | 0 | ( 0%) | 0 | ( 0%) | 2 | ( <1%) | 5 | ( 1%) |
| Outdoor: Smoke source: Other | 0 | ( 0%) | 0 | ( 0%) | 0 | ( 0%) | 0 | ( 0%) | 0 | ( 0%) | 0 | ( 0%) | 1 | ( <1%) | 0 | ( 0%) |
| Missing | 0 | | 0 | | 0 | | 0 | | 2 | | 0 | | 0 | | 2 | |

| | Guatemala | | | | India | | | | Peru | | | | Rwanda | | | |
|---|---|---|---|---|---|---|---|---|---|---|---|---|---|---|---|---|
| Variable | Control (N=400) | | Intervention (N=400) | | Control (N=399) | | Intervention (N=400) | | Control (N=402) | | Intervention (N=396) | | Control (N=404) | | Intervention (N=394) | |
| **Exposure Compliance** | | | | | | | | | | | | | | | | |
| **Other smoke source to participants** | | | | | | | | | | | | | | | | |
| No | 328 | ( 82%) | 332 | ( 83%) | 390 | ( 98%) | 391 | ( 98%) | 371 | ( 93%) | 360 | ( 91%) | 378 | ( 94%) | 358 | ( 91%) |
| Yes | 72 | ( 18%) | 68 | ( 17%) | 9 | ( 2%) | 9 | ( 2%) | 27 | ( 7%) | 35 | ( 9%) | 26 | ( 6%) | 34 | ( 9%) |

| | | | | | | | | | | | | | | | | |
|---|---|---|---|---|---|---|---|---|---|---|---|---|---|---|---|---|
| Missing | 0 | | 0 | | 0 | | 0 | | 4 | | 1 | | 0 | | 2 | |
| **Detailed other smoke sources to participants**[g] | | | | | | | | | | | | | | | | |
| Combustion powered corn/flour mill | 0 | ( 0%) | 0 | ( 0%) | 0 | ( 0%) | 0 | ( 0%) | 0 | ( 0%) | 0 | ( 0%) | 1 | ( 4%) | 0 | ( 0%) |
| Other kitchen | 1 | ( 1%) | 4 | ( 6%) | 7 | ( 78%) | 6 | ( 67%) | 10 | ( 37%) | 7 | ( 20%) | 13 | ( 50%) | 15 | ( 44%) |
| Outside of home | 0 | ( 0%) | 1 | ( 1%) | 0 | ( 0%) | 1 | ( 11%) | 0 | ( 0%) | 2 | ( 6%) | 2 | ( 8%) | 7 | ( 21%) |
| Trash burning | 1 | ( 1%) | 1 | ( 1%) | 2 | ( 22%) | 1 | ( 11%) | 11 | ( 41%) | 12 | ( 34%) | 1 | ( 4%) | 1 | ( 3%) |
| Tobacco smoking | 0 | ( 0%) | 0 | ( 0%) | 0 | ( 0%) | 0 | ( 0%) | 0 | ( 0%) | 0 | ( 0%) | 5 | ( 19%) | 5 | ( 15%) |
| Burning of agricultural waste | 0 | ( 0%) | 0 | ( 0%) | 0 | ( 0%) | 0 | ( 0%) | 0 | ( 0%) | 1 | ( 3%) | 1 | ( 4%) | 1 | ( 3%) |
| Burning from preparing fields | 0 | ( 0%) | 0 | ( 0%) | 0 | ( 0%) | 0 | ( 0%) | 1 | ( 4%) | 7 | ( 20%) | 0 | ( 0%) | 1 | ( 3%) |
| Incense | 0 | ( 0%) | 1 | ( 1%) | 0 | ( 0%) | 0 | ( 0%) | 1 | ( 4%) | 3 | ( 9%) | 0 | ( 0%) | 0 | ( 0%) |
| Generator | 28 | ( 39%) | 21 | ( 31%) | 0 | ( 0%) | 1 | ( 11%) | 0 | ( 0%) | 0 | ( 0%) | 0 | ( 0%) | 0 | ( 0%) |
| Intense vehicular emissions | 42 | ( 58%) | 40 | ( 59%) | 0 | ( 0%) | 0 | ( 0%) | 3 | ( 11%) | 1 | ( 3%) | 3 | ( 12%) | 4 | ( 12%) |
| Missing | 0 | | 0 | | 0 | | 0 | | 0 | | 0 | | 0 | | 0 | |
| **Baseline Exposure** | | | | | | | | | | | | | | | | |
| **HH: Primary cook** | | | | | | | | | | | | | | | | |
| Myself | 337 | ( 84%) | 339 | ( 85%) | 374 | ( 94%) | 383 | ( 96%) | 221 | ( 55%) | 234 | ( 59%) | 386 | ( 96%) | 351 | ( 89%) |
| Mother/Mother-in-law | 56 | ( 14%) | 57 | ( 14%) | 21 | ( 5%) | 14 | ( 4%) | 173 | ( 43%) | 158 | ( 40%) | 4 | ( 1%) | 1 | (<1%) |
| Sister/Sister-in-law | 6 | ( 2%) | 1 | (<1%) | 3 | ( 1%) | 1 | (<1%) | 1 | (<1%) | 3 | ( 1%) | 5 | ( 1%) | 9 | ( 2%) |
| Daughter | 0 | ( 0%) | 2 | ( 1%) | 0 | ( 0%) | 1 | (<1%) | 1 | (<1%) | 0 | ( 0%) | 2 | (<1%) | 6 | ( 2%) |
| Hired cook in the home | 0 | ( 0%) | 0 | ( 0%) | 0 | ( 0%) | 0 | ( 0%) | 0 | ( 0%) | 0 | ( 0%) | 2 | (<1%) | 23 | ( 6%) |
| Other | 1 | (<1%) | 0 | ( 0%) | 1 | (<1%) | 1 | (<1%) | 5 | ( 1%) | 1 | (<1%) | 4 | ( 1%) | 3 | ( 1%) |
| Missing | 0 | | 1 | | 0 | | 0 | | 1 | | 0 | | 1 | | 1 | |
| **HH: Cook Times/day** | | | | | | | | | | | | | | | | |
| None | 0 | ( 0%) | 0 | ( 0%) | 0 | ( 0%) | 0 | ( 0%) | 0 | ( 0%) | 1 | (<1%) | 0 | ( 0%) | 0 | ( 0%) |
| One or two | 11 | ( 3%) | 7 | ( 2%) | 339 | ( 85%) | 343 | ( 86%) | 164 | ( 41%) | 180 | ( 45%) | 357 | ( 89%) | 322 | ( 82%) |
| Three or more | 389 | ( 97%) | 393 | ( 98%) | 60 | ( 15%) | 57 | ( 14%) | 237 | ( 59%) | 215 | ( 54%) | 46 | ( 11%) | 71 | ( 18%) |
| Missing | 0 | | 0 | | 0 | | 0 | | 1 | | 0 | | 1 | | 1 | |
| **Mother: cook times/week**[h] | | | | | | | | | | | | | | | | |
| One to Seven | 20 | ( 5%) | 12 | ( 3%) | 53 | ( 13%) | 66 | ( 17%) | 102 | ( 26%) | 89 | ( 22%) | 56 | ( 14%) | 90 | ( 23%) |
| Eight to Fourteen | 33 | ( 8%) | 20 | ( 5%) | 301 | ( 75%) | 287 | ( 72%) | 150 | ( 38%) | 154 | ( 39%) | 308 | ( 76%) | 259 | ( 66%) |
| Fifteen or more | 347 | ( 87%) | 368 | ( 92%) | 45 | ( 11%) | 47 | ( 12%) | 148 | ( 37%) | 153 | ( 39%) | 39 | ( 10%) | 44 | ( 11%) |
| Missing | 0 | | 0 | | 0 | | 0 | | 2 | | 0 | | 1 | | 1 | |
| **Persons who prepare the meals other than the primary cook**[i] | | | | | | | | | | | | | | | | |
| Other Cook: Myself | 81 | ( 20%) | 81 | ( 20%) | 42 | ( 11%) | 34 | ( 9%) | 197 | ( 49%) | 199 | ( 50%) | 30 | ( 7%) | 56 | ( 14%) |
| Other Cook: Mother/in-law | 73 | ( 18%) | 91 | ( 23%) | 145 | ( 36%) | 176 | ( 44%) | 87 | ( 22%) | 74 | ( 19%) | 21 | ( 5%) | 17 | ( 4%) |
| Other Cook: Sister/in-law | 46 | ( 12%) | 44 | ( 11%) | 26 | ( 7%) | 22 | ( 6%) | 36 | ( 9%) | 31 | ( 8%) | 36 | ( 9%) | 41 | ( 10%) |
| Other Cook: Daughter | 11 | ( 3%) | 17 | ( 4%) | 0 | ( 0%) | 0 | ( 0%) | 15 | ( 4%) | 17 | ( 4%) | 36 | ( 9%) | 22 | ( 6%) |
| Other Cook: Hired cook | 1 | (<1%) | 0 | ( 0%) | 0 | ( 0%) | 0 | ( 0%) | 0 | ( 0%) | 0 | ( 0%) | 9 | ( 2%) | 13 | ( 3%) |
| Other Cook: Other | 24 | ( 6%) | 17 | ( 4%) | 12 | ( 3%) | 9 | ( 2%) | 108 | ( 27%) | 104 | ( 26%) | 138 | ( 34%) | 132 | ( 34%) |
| Missing | 0 | | 0 | | 0 | | 0 | | 0 | | 0 | | 1 | | 1 | |

| | Guatemala | | | | India | | | | Peru | | | | Rwanda | | | |
|---|---|---|---|---|---|---|---|---|---|---|---|---|---|---|---|---|
| **Variable** | **Control (N=400)** | | **Intervention (N=400)** | | **Control (N=399)** | | **Intervention (N=400)** | | **Control (N=402)** | | **Intervention (N=396)** | | **Control (N=404)** | | **Intervention (N=394)** | |
| **# of Stoves** | | | | | | | | | | | | | | | | |
| One | 138 | ( 35%) | 135 | ( 34%) | 271 | ( 68%) | 282 | ( 71%) | 105 | ( 26%) | 90 | ( 23%) | 248 | ( 62%) | 228 | ( 58%) |
| Two or more | 262 | ( 66%) | 265 | ( 66%) | 128 | ( 32%) | 118 | ( 30%) | 295 | ( 74%) | 304 | ( 77%) | 154 | ( 38%) | 165 | ( 42%) |
| Missing | 0 | | 0 | | 0 | | 0 | | 2 | | 1 | | 1 | | 1 | |
| **Stove1: chimney** | | | | | | | | | | | | | | | | |
| No | 310 | ( 78%) | 313 | ( 78%) | 397 | ( 99%) | 398 | (100%) | 249 | ( 62%) | 252 | ( 64%) | 382 | ( 95%) | 379 | ( 96%) |
| Yes | 90 | ( 23%) | 87 | ( 22%) | 2 | ( 1%) | 2 | ( 1%) | 152 | ( 38%) | 144 | ( 36%) | 21 | ( 5%) | 14 | ( 4%) |

| | | | | | | | | | | | | | | | | |
|---|---|---|---|---|---|---|---|---|---|---|---|---|---|---|---|---|
| Missing | 0 | | 0 | | 0 | | 0 | | 1 | | 0 | | 1 | | 1 | |
| **Stove1: used marks/signs** | | | | | | | | | | | | | | | | |
| Stove used during the visit | 279 | ( 70%) | 265 | ( 66%) | 117 | ( 29%) | 133 | ( 33%) | 139 | ( 35%) | 122 | ( 31%) | 133 | ( 33%) | 150 | ( 38%) |
| Stove not used in visit but used recently | 120 | ( 30%) | 133 | ( 33%) | 281 | ( 70%) | 264 | ( 66%) | 240 | ( 60%) | 243 | ( 62%) | 270 | ( 67%) | 242 | ( 62%) |
| Stove not used recently | 1 | ( <1%) | 1 | ( <1%) | 1 | ( <1%) | 3 | ( 1%) | 21 | ( 5%) | 28 | ( 7%) | 0 | ( 0%) | 0 | ( 0%) |
| unable to observe stove | 0 | ( 0%) | 0 | ( 0%) | 0 | ( 0%) | 0 | ( 0%) | 1 | ( <1%) | 2 | ( 1%) | 0 | ( 0%) | 1 | ( <1%) |
| Missing | 0 | | 1 | | 0 | | 0 | | 1 | | 1 | | 1 | | 1 | |
| **Stove1: location** | | | | | | | | | | | | | | | | |
| In participant's bedroom | 18 | ( 5%) | 19 | ( 5%) | 73 | ( 18%) | 97 | ( 24%) | 1 | ( <1%) | 4 | ( 1%) | 2 | ( <1%) | 4 | ( 1%) |
| Room adjacent to bedroom | 198 | ( 50%) | 184 | ( 46%) | 146 | ( 37%) | 121 | ( 30%) | 29 | ( 7%) | 22 | ( 6%) | 10 | ( 2%) | 16 | ( 4%) |
| Separated from bedroom, inside | 104 | ( 26%) | 123 | ( 31%) | 70 | ( 18%) | 74 | ( 19%) | 69 | ( 17%) | 71 | ( 18%) | 18 | ( 4%) | 19 | ( 5%) |
| Outside the house | 5 | ( 1%) | 7 | ( 2%) | 1 | ( <1%) | 2 | ( 1%) | 96 | ( 24%) | 94 | ( 24%) | 123 | ( 31%) | 125 | ( 32%) |
| In separate building | 74 | ( 19%) | 67 | ( 17%) | 109 | ( 27%) | 106 | ( 27%) | 205 | ( 51%) | 204 | ( 52%) | 250 | ( 62%) | 229 | ( 58%) |
| Other | 0 | ( 0%) | 0 | ( 0%) | 0 | ( 0%) | 0 | ( 0%) | 1 | ( <1%) | 1 | ( <1%) | 0 | ( 0%) | 0 | ( 0%) |
| Missing | 1 | | 0 | | 0 | | 0 | | 1 | | 0 | | 1 | | 1 | |
| **Stove1: roof** | | | | | | | | | | | | | | | | |
| No | 6 | ( 2%) | 4 | ( 1%) | 10 | ( 3%) | 8 | ( 2%) | 93 | ( 23%) | 88 | ( 22%) | 125 | ( 31%) | 131 | ( 33%) |
| Yes | 394 | ( 99%) | 396 | ( 99%) | 389 | ( 97%) | 392 | ( 98%) | 308 | ( 77%) | 308 | ( 78%) | 278 | ( 69%) | 262 | ( 67%) |
| **Stove1: # of walls** | | | | | | | | | | | | | | | | |
| None | 5 | ( 1%) | 5 | ( 1%) | 15 | ( 4%) | 7 | ( 2%) | 39 | ( 10%) | 45 | ( 11%) | 118 | ( 29%) | 117 | ( 30%) |
| One to three | 29 | ( 7%) | 36 | ( 9%) | 75 | ( 19%) | 67 | ( 17%) | 85 | ( 21%) | 62 | ( 16%) | 9 | ( 2%) | 13 | ( 3%) |
| Four | 366 | ( 92%) | 359 | ( 90%) | 309 | ( 77%) | 326 | ( 82%) | 274 | ( 69%) | 288 | ( 73%) | 276 | ( 68%) | 263 | ( 67%) |
| Missing | 0 | | 0 | | 0 | | 0 | | 4 | | 1 | | 1 | | 1 | |
| **Stove1: used days/week** | | | | | | | | | | | | | | | | |
| Less than seven days | 21 | ( 5%) | 22 | ( 6%) | 6 | ( 2%) | 4 | ( 1%) | 124 | ( 31%) | 110 | ( 28%) | 84 | ( 21%) | 76 | ( 19%) |
| Seven days | 379 | ( 95%) | 378 | ( 95%) | 393 | ( 98%) | 396 | ( 99%) | 277 | ( 69%) | 286 | ( 72%) | 319 | ( 79%) | 317 | ( 81%) |
| Missing | 0 | | 0 | | 0 | | 0 | | 1 | | 0 | | 1 | | 1 | |
| **Stove1: used hours/day** | | | | | | | | | | | | | | | | |
| One or two | 47 | ( 12%) | 47 | ( 12%) | 148 | ( 37%) | 129 | ( 32%) | 125 | ( 31%) | 128 | ( 32%) | 152 | ( 38%) | 142 | ( 36%) |
| Three or more | 352 | ( 88%) | 353 | ( 88%) | 251 | ( 63%) | 271 | ( 68%) | 275 | ( 69%) | 268 | ( 68%) | 251 | ( 62%) | 251 | ( 64%) |
| Missing | 1 | | 0 | | 0 | | 0 | | 2 | | 0 | | 1 | | 1 | |
| **Stove1: Last time** | | | | | | | | | | | | | | | | |
| Within past day | 398 | (100%) | 397 | ( 99%) | 396 | ( 99%) | 393 | ( 98%) | 368 | ( 92%) | 349 | ( 88%) | 399 | ( 99%) | 389 | ( 99%) |
| Within past week | 2 | ( 1%) | 2 | ( 1%) | 3 | ( 1%) | 5 | ( 1%) | 27 | ( 7%) | 46 | ( 12%) | 3 | ( 1%) | 4 | ( 1%) |
| Within past month | 0 | ( 0%) | 0 | ( 0%) | 0 | ( 0%) | 1 | ( <1%) | 3 | ( 1%) | 0 | ( 0%) | 0 | ( 0%) | 0 | ( 0%) |
| more than one month ago | 0 | ( 0%) | 1 | ( <1%) | 0 | ( 0%) | 1 | ( <1%) | 2 | ( 1%) | 1 | ( <1%) | 1 | ( <1%) | 0 | ( 0%) |
| Missing | 0 | | 0 | | 0 | | 0 | | 2 | | 0 | | 1 | | 1 | |

| Variable | Guatemala Control (N=400) | | Guatemala Intervention (N=400) | | India Control (N=399) | | India Intervention (N=400) | | Peru Control (N=402) | | Peru Intervention (N=396) | | Rwanda Control (N=404) | | Rwanda Intervention (N=394) | |
|---|---|---|---|---|---|---|---|---|---|---|---|---|---|---|---|---|
| **Baseline Exposure** | | | | | | | | | | | | | | | | |
| **The main stove is used for[j]** | | | | | | | | | | | | | | | | |
| Stove1: Cooking food | 400 | (100%) | 399 | (100%) | 396 | ( 99%) | 394 | ( 99%) | 396 | ( 99%) | 391 | ( 99%) | 402 | (100%) | 391 | ( 99%) |
| Stove1: Roasting food | 382 | ( 96%) | 377 | ( 94%) | 193 | ( 48%) | 195 | ( 49%) | 8 | ( 2%) | 10 | ( 3%) | 279 | ( 69%) | 272 | ( 69%) |
| Stove1: Making animal food | 22 | ( 6%) | 16 | ( 4%) | 72 | ( 18%) | 60 | ( 15%) | 319 | ( 79%) | 293 | ( 74%) | 30 | ( 7%) | 33 | ( 8%) |
| Stove1: Making alcohol | 3 | ( 1%) | 2 | ( 1%) | 0 | ( 0%) | 2 | ( 1%) | 0 | ( 0%) | 3 | ( 1%) | 9 | ( 2%) | 8 | ( 2%) |
| Stove1: Heating water for bathing | 381 | ( 95%) | 386 | ( 97%) | 268 | ( 67%) | 259 | ( 65%) | 364 | ( 91%) | 357 | ( 90%) | 323 | ( 80%) | 308 | ( 78%) |
| Stove1: Heating water--cleaning,washing | 216 | ( 54%) | 195 | ( 49%) | 41 | ( 10%) | 42 | ( 11%) | 320 | ( 80%) | 307 | ( 78%) | 55 | ( 14%) | 54 | ( 14%) |
| Stove1: Boiling drinking water/tea/coffee | 387 | ( 97%) | 383 | ( 96%) | 361 | ( 90%) | 363 | ( 91%) | 386 | ( 96%) | 384 | ( 97%) | 293 | ( 73%) | 305 | ( 78%) |
| Stove1: Heating home | 203 | ( 51%) | 218 | ( 55%) | 39 | ( 10%) | 47 | ( 12%) | 12 | ( 3%) | 27 | ( 7%) | 2 | ( <1%) | 3 | ( 1%) |

| | | | | | | | | | | | | | | | | |
|---|---|---|---|---|---|---|---|---|---|---|---|---|---|---|---|---|
| Stove1: Providing light | 38 | ( 10%) | 43 | ( 11%) | 11 | ( 3%) | 13 | ( 3%) | 0 | ( 0%) | 0 | ( 0%) | 3 | ( 1%) | 0 | ( 0%) |
| Stove1: Business | 11 | ( 3%) | 18 | ( 5%) | 0 | ( 0%) | 1 | (<1%) | 11 | ( 3%) | 10 | ( 3%) | 5 | ( 1%) | 6 | ( 2%) |
| Missing | 0 | | 0 | | 0 | | 0 | | 0 | | 0 | | 1 | | 1 | |
| **Primary Fuel type** | | | | | | | | | | | | | | | | |
| Cow dung | 0 | ( 0%) | 0 | ( 0%) | 0 | ( 0%) | 0 | ( 0%) | 351 | ( 88%) | 346 | ( 87%) | 0 | ( 0%) | 0 | ( 0%) |
| Wood | 394 | ( 99%) | 399 | (100%) | 399 | (100%) | 400 | (100%) | 48 | ( 12%) | 42 | ( 11%) | 323 | ( 80%) | 257 | ( 65%) |
| Charcoal | 0 | ( 0%) | 0 | ( 0%) | 0 | ( 0%) | 0 | ( 0%) | 0 | ( 0%) | 0 | ( 0%) | 72 | ( 18%) | 125 | ( 32%) |
| Other | 2 | ( 1%) | 1 | (<1%) | 0 | ( 0%) | 0 | ( 0%) | 2 | (<1%) | 8 | ( 2%) | 8 | ( 2%) | 11 | ( 3%) |
| Missing | 4 | | 0 | | 0 | | 0 | | 1 | | 0 | | 1 | | 1 | |
| **Stove1: collect/purchase fuel** | | | | | | | | | | | | | | | | |
| Collect only | 179 | ( 45%) | 157 | ( 39%) | 351 | ( 88%) | 355 | ( 89%) | 382 | ( 95%) | 368 | ( 93%) | 255 | ( 63%) | 170 | ( 43%) |
| Purchase only | 115 | ( 29%) | 127 | ( 32%) | 43 | ( 11%) | 42 | ( 11%) | 13 | ( 3%) | 18 | ( 5%) | 119 | ( 30%) | 196 | ( 50%) |
| Given to use | 1 | (<1%) | 1 | (<1%) | 0 | ( 0%) | 0 | ( 0%) | 1 | (<1%) | 2 | ( 1%) | 0 | ( 0%) | 1 | (<1%) |
| Both collect and buy | 104 | ( 26%) | 115 | ( 29%) | 5 | ( 1%) | 3 | ( 1%) | 4 | ( 1%) | 4 | ( 1%) | 29 | ( 7%) | 26 | ( 7%) |
| Other | 1 | (<1%) | 0 | ( 0%) | 0 | ( 0%) | 0 | ( 0%) | 1 | (<1%) | 4 | ( 1%) | 0 | ( 0%) | 0 | ( 0%) |
| Missing | 0 | | 0 | | 0 | | 0 | | 1 | | 0 | | 1 | | 1 | |
| **Stove1: purchase fuel money/month in USD($)[k]** | | | | | | | | | | | | | | | | |
| Mean (SD) | 26.2 | (20.1) | 26.5 | (22.8) | 10.5 | (7.9) | 10.5 | (7.6) | 6.0 | (6.9) | 9.6 | (10.3) | 7.5 | (10.1) | 7.7 | (9.0) |
| [Range] | | [1-104] | | [0-195] | | [1-39] | | [1-42] | | [1-31] | | [1-39] | | [0-120] | | [0-126] |
| N | | 219 | | 242 | | 48 | | 45 | | 17 | | 22 | | 148 | | 222 |
| Missing | | 0 | | 0 | | 0 | | 0 | | 0 | | 0 | | 0 | | 0 |
| **Stove1: collect fuel time/week** | | | | | | | | | | | | | | | | |
| Mean (SD) | 6.2 | (13.6) | 5.4 | (11.2) | 5.7 | (4.4) | 6.1 | (5.2) | 3.7 | (3.2) | 3.6 | (2.9) | 60.3 | (557.6) | 68.1 | (676.0) |
| [Range] | | [0-200] | | [0-180] | | [1-30] | | [1-35] | | [1-28] | | [1-20] | | [0-7,000] | | [0-9,000] |
| N | | 283 | | 271 | | 356 | | 358 | | 386 | | 372 | | 284 | | 196 |
| Missing | | 0 | | 1 | | 0 | | 0 | | 0 | | 0 | | 0 | | 0 |
| **Primary lighting source** | | | | | | | | | | | | | | | | |
| Torch (battery) | 5 | ( 1%) | 5 | ( 1%) | 4 | ( 1%) | 7 | ( 2%) | 4 | ( 1%) | 8 | ( 2%) | 90 | ( 22%) | 83 | ( 21%) |
| Kerosene lamp | 1 | (<1%) | 5 | ( 1%) | 9 | ( 2%) | 7 | ( 2%) | 1 | (<1%) | 0 | ( 0%) | 37 | ( 9%) | 22 | ( 6%) |
| Solar light | 0 | ( 0%) | 2 | ( 1%) | 2 | ( 1%) | 1 | (<1%) | 12 | ( 3%) | 11 | ( 3%) | 138 | ( 34%) | 119 | ( 30%) |
| Electricity | 361 | ( 90%) | 348 | ( 87%) | 384 | ( 96%) | 385 | ( 96%) | 371 | ( 93%) | 359 | ( 91%) | 88 | ( 22%) | 138 | ( 35%) |
| Other | 33 | ( 8%) | 40 | ( 10%) | 0 | ( 0%) | 0 | ( 0%) | 13 | ( 3%) | 18 | ( 5%) | 50 | ( 12%) | 31 | ( 8%) |
| Missing | 0 | | 0 | | 0 | | 0 | | 1 | | 0 | | 1 | | 1 | |

| | Guatemala | | | | India | | | | Peru | | | | Rwanda | | | |
|---|---|---|---|---|---|---|---|---|---|---|---|---|---|---|---|---|
| Variable | Control (N=400) | | Intervention (N=400) | | Control (N=399) | | Intervention (N=400) | | Control (N=402) | | Intervention (N=396) | | Control (N=404) | | Intervention (N=394) | |
| **Baseline Exposure** | | | | | | | | | | | | | | | | |
| **Primary heating source** | | | | | | | | | | | | | | | | |
| Do not use heating | 268 | ( 67%) | 262 | ( 66%) | 368 | ( 92%) | 362 | ( 91%) | 401 | (100%) | 393 | ( 99%) | 399 | ( 99%) | 390 | ( 99%) |
| Traditional cookstove/Three-stone fire | 111 | ( 28%) | 116 | ( 29%) | 27 | ( 7%) | 35 | ( 9%) | 0 | ( 0%) | 2 | ( 1%) | 4 | ( 1%) | 2 | ( 1%) |
| Other | 21 | ( 5%) | 22 | ( 6%) | 4 | ( 1%) | 3 | ( 1%) | 0 | ( 0%) | 1 | (<1%) | 0 | ( 0%) | 1 | (<1%) |
| Missing | 0 | | 0 | | 0 | | 0 | | 1 | | 0 | | 1 | | 1 | |
| **Main way garbage disposal** | | | | | | | | | | | | | | | | |
| Throw it away on own land or nearby | 170 | ( 43%) | 156 | ( 39%) | 256 | ( 64%) | 285 | ( 71%) | 46 | ( 11%) | 39 | ( 10%) | 138 | ( 34%) | 133 | ( 34%) |
| Bury it | 51 | ( 13%) | 43 | ( 11%) | 9 | ( 2%) | 2 | ( 1%) | 107 | ( 27%) | 99 | ( 25%) | 1 | (<1%) | 0 | ( 0%) |
| Burn it | 68 | ( 17%) | 91 | ( 23%) | 63 | ( 16%) | 59 | ( 15%) | 194 | ( 48%) | 198 | ( 50%) | 1 | (<1%) | 0 | ( 0%) |
| Compost | 90 | ( 23%) | 87 | ( 22%) | 15 | ( 4%) | 10 | ( 3%) | 0 | ( 0%) | 2 | ( 1%) | 255 | ( 63%) | 242 | ( 62%) |
| Collected by government | 0 | ( 0%) | 1 | (<1%) | 56 | ( 14%) | 44 | ( 11%) | 48 | ( 12%) | 55 | ( 14%) | 6 | ( 1%) | 6 | ( 2%) |
| Collected by private service | 0 | ( 0%) | 1 | (<1%) | 0 | ( 0%) | 0 | ( 0%) | 0 | ( 0%) | 0 | ( 0%) | 2 | (<1%) | 11 | ( 3%) |

| | | | | | | | | | | | | | | | | |
|---|---|---|---|---|---|---|---|---|---|---|---|---|---|---|---|---|
| Other | 18 | ( 5%) | 21 | ( 5%) | 0 | ( 0%) | 0 | ( 0%) | 6 | ( 1%) | 3 | ( 1%) | 0 | ( 0%) | 1 | (<1%) |
| Missing | 3 | | 0 | | 0 | | 0 | | 1 | | 0 | | 1 | | 1 | |
| **Secondary way garbage disposal** | | | | | | | | | | | | | | | | |
| None | 37 | ( 9%) | 35 | ( 9%) | 237 | ( 59%) | 214 | ( 54%) | 142 | ( 35%) | 171 | ( 43%) | 260 | ( 65%) | 252 | ( 64%) |
| Throw it away on own land or nearby | 57 | ( 14%) | 64 | ( 16%) | 10 | ( 3%) | 7 | ( 2%) | 24 | ( 6%) | 14 | ( 4%) | 111 | ( 28%) | 100 | ( 25%) |
| Bury it | 24 | ( 6%) | 20 | ( 5%) | 4 | ( 1%) | 4 | ( 1%) | 81 | ( 20%) | 92 | ( 23%) | 4 | ( 1%) | 0 | ( 0%) |
| Burn it | 258 | ( 65%) | 248 | ( 62%) | 102 | ( 26%) | 131 | ( 33%) | 138 | ( 34%) | 112 | ( 28%) | 7 | ( 2%) | 7 | ( 2%) |
| Compost | 15 | ( 4%) | 17 | ( 4%) | 19 | ( 5%) | 20 | ( 5%) | 0 | ( 0%) | 0 | ( 0%) | 21 | ( 5%) | 30 | ( 8%) |
| Collected by government | 0 | ( 0%) | 0 | ( 0%) | 27 | ( 7%) | 24 | ( 6%) | 12 | ( 3%) | 2 | ( 1%) | 0 | ( 0%) | 2 | ( 1%) |
| Collected by private service | 1 | ( <1%) | 3 | ( 1%) | 0 | ( 0%) | 0 | ( 0%) | 0 | ( 0%) | 0 | ( 0%) | 0 | ( 0%) | 2 | ( 1%) |
| Other | 8 | ( 2%) | 12 | ( 3%) | 0 | ( 0%) | 0 | ( 0%) | 4 | ( 1%) | 5 | ( 1%) | 0 | ( 0%) | 0 | ( 0%) |
| Missing | 0 | | 1 | | 0 | | 0 | | 1 | | 0 | | 1 | | 1 | |
| **What other ways do you dispose of garbage from your home?[l]** | | | | | | | | | | | | | | | | |
| Other garbage disp: Throw it away | 8 | ( 2%) | 9 | ( 2%) | 1 | ( <1%) | 0 | ( 0%) | 5 | ( 1%) | 8 | ( 2%) | 8 | ( 2%) | 9 | ( 2%) |
| Other garbage disp: Bury | 10 | ( 3%) | 6 | ( 2%) | 1 | ( <1%) | 0 | ( 0%) | 17 | ( 4%) | 19 | ( 5%) | 3 | ( 1%) | 0 | ( 0%) |
| Other garbage disp: Burn | 19 | ( 5%) | 14 | ( 4%) | 9 | ( 2%) | 5 | ( 1%) | 14 | ( 3%) | 21 | ( 5%) | 4 | ( 1%) | 2 | ( 1%) |
| Other garbage disp: Compost | 6 | ( 2%) | 4 | ( 1%) | 0 | ( 0%) | 5 | ( 1%) | 1 | ( <1%) | 0 | ( 0%) | 9 | ( 2%) | 7 | ( 2%) |
| Other garbage disp: govt collection | 1 | ( <1%) | 0 | ( 0%) | 1 | ( <1%) | 4 | ( 1%) | 2 | ( <1%) | 2 | ( 1%) | 0 | ( 0%) | 0 | ( 0%) |
| Other garbage disp: private collection | 0 | ( 0%) | 0 | ( 0%) | 0 | ( 0%) | 0 | ( 0%) | 0 | ( 0%) | 0 | ( 0%) | 1 | ( <1%) | 0 | ( 0%) |
| Other garbage disp: Other | 4 | ( 1%) | 2 | ( 1%) | 0 | ( 0%) | 0 | ( 0%) | 2 | ( <1%) | 1 | ( <1%) | 0 | ( 0%) | 0 | ( 0%) |
| Missing | 0 | | 0 | | 0 | | 0 | | 0 | | 0 | | 1 | | 1 | |
| **Burn garbage in any way** | | | | | | | | | | | | | | | | |
| No | 61 | ( 15%) | 59 | ( 15%) | 227 | ( 57%) | 208 | ( 52%) | 65 | ( 16%) | 75 | ( 19%) | 391 | ( 97%) | 385 | ( 98%) |
| Yes | 339 | ( 85%) | 341 | ( 85%) | 172 | ( 43%) | 192 | ( 48%) | 336 | ( 84%) | 321 | ( 81%) | 12 | ( 3%) | 8 | ( 2%) |
| Missing | 0 | | 0 | | 0 | | 0 | | 1 | | 0 | | 1 | | 1 | |
| **Burn location** | | | | | | | | | | | | | | | | |
| Inside the house | 65 | ( 19%) | 55 | ( 16%) | 0 | ( 0%) | 2 | ( 1%) | 9 | ( 3%) | 12 | ( 4%) | 0 | ( 0%) | 0 | ( 0%) |
| Outside the house | 271 | ( 80%) | 284 | ( 84%) | 172 | (100%) | 190 | ( 99%) | 326 | ( 97%) | 308 | ( 96%) | 9 | ( 75%) | 8 | (100%) |
| Other | 2 | ( 1%) | 0 | ( 0%) | 0 | ( 0%) | 0 | ( 0%) | 1 | ( <1%) | 0 | ( 0%) | 3 | ( 25%) | 0 | ( 0%) |
| Missing | 1 | | 2 | | 0 | | 0 | | 0 | | 1 | | 0 | | 0 | |

| | Guatemala | | | | India | | | | Peru | | | | Rwanda | | | |
|---|---|---|---|---|---|---|---|---|---|---|---|---|---|---|---|---|
| Variable | Control (N=400) | | Intervention (N=400) | | Control (N=399) | | Intervention (N=400) | | Control (N=402) | | Intervention (N=396) | | Control (N=404) | | Intervention (N=394) | |
| **Baseline Exposure** | | | | | | | | | | | | | | | | |
| **Burn location: distance to entrance** | | | | | | | | | | | | | | | | |
| Mean (SD) | 13.7 | (61.6) | 16.1 | (75.6) | 21.3 | (15.7) | 23.0 | (22.4) | 62.2 | (125.0) | 66.0 | (160.2) | 20.6 | (30.3) | 13.5 | (16.0) |
| [Range] | | [1-1,000] | | [1-1,000] | | [5-100] | | [1-200] | | [1-1,000] | | [2-1,500] | | [3-100] | | [2-50] |
| N | | 271 | | 284 | | 172 | | 190 | | 326 | | 308 | | 9 | | 8 |
| Missing | | 0 | | 0 | | 0 | | 0 | | 0 | | 0 | | 0 | | 0 |
| **Burn times per month** | | | | | | | | | | | | | | | | |
| None | 1 | ( <1%) | 1 | ( <1%) | 0 | ( 0%) | 0 | ( 0%) | 0 | ( 0%) | 3 | ( 1%) | 4 | ( 33%) | 2 | ( 25%) |
| One or two | 135 | ( 40%) | 118 | ( 35%) | 81 | ( 52%) | 89 | ( 51%) | 205 | ( 61%) | 184 | ( 57%) | 7 | ( 58%) | 5 | ( 63%) |
| Three or more | 203 | ( 60%) | 221 | ( 65%) | 76 | ( 48%) | 84 | ( 49%) | 129 | ( 39%) | 134 | ( 42%) | 1 | ( 8%) | 1 | ( 13%) |
| Missing | 0 | | 1 | | 15 | | 19 | | 2 | | 0 | | 0 | | 0 | |
| **Burn leaves/debris--mosquitos (not for Peru)** | | | | | | | | | | | | | | | | |
| No | 266 | ( 67%) | 266 | ( 67%) | 351 | ( 88%) | 355 | ( 89%) | N/A | N/A | N/A | N/A | 381 | ( 95%) | 356 | ( 91%) |
| Yes | 131 | ( 33%) | 134 | ( 34%) | 48 | ( 12%) | 45 | ( 11%) | N/A | N/A | N/A | N/A | 22 | ( 5%) | 37 | ( 9%) |
| Missing | 3 | | 0 | | 0 | | 0 | | N/A | N/A | N/A | N/A | 1 | | 1 | |
| **Burn leaves/debris times/wk** | | | | | | | | | | | | | | | | |
| None | 1 | ( 1%) | 2 | ( 2%) | 0 | ( 0%) | 0 | ( 0%) | 0 | ( 0%) | 0 | ( 0%) | 1 | ( 5%) | 1 | ( 3%) |

| | | | | | | | | | | | | | | | | |
|---|---|---|---|---|---|---|---|---|---|---|---|---|---|---|---|---|
| One | 50 | ( 38%) | 37 | ( 28%) | 8 | ( 17%) | 10 | ( 22%) | 0 | ( 0%) | 0 | ( 0%) | 15 | ( 68%) | 28 | ( 76%) |
| Two or more | 80 | ( 61%) | 93 | ( 70%) | 40 | ( 83%) | 35 | ( 78%) | 0 | ( 0%) | 0 | ( 0%) | 6 | ( 27%) | 8 | ( 22%) |
| Missing | 0 | | 2 | | 0 | | 0 | | 0 | ( 0%) | 0 | ( 0%) | 0 | | 0 | |
| **Smoke from neighbor comes inside** | | | | | | | | | | | | | | | | |
| No | 236 | ( 59%) | 236 | ( 59%) | 276 | ( 69%) | 283 | ( 71%) | 217 | ( 54%) | 228 | ( 58%) | 256 | ( 64%) | 241 | ( 61%) |
| Yes | 163 | ( 41%) | 163 | ( 41%) | 123 | ( 31%) | 117 | ( 29%) | 184 | ( 46%) | 168 | ( 42%) | 147 | ( 36%) | 152 | ( 39%) |
| Missing | 1 | | 1 | | 0 | | 0 | | 1 | | 0 | | 1 | | 1 | |
| **Someone in household smokes** | | | | | | | | | | | | | | | | |
| No | 378 | ( 95%) | 378 | ( 95%) | 265 | ( 66%) | 281 | ( 70%) | 397 | ( 99%) | 392 | ( 99%) | 381 | ( 95%) | 385 | ( 98%) |
| Yes | 22 | ( 6%) | 22 | ( 6%) | 134 | ( 34%) | 119 | ( 30%) | 3 | ( 1%) | 4 | ( 1%) | 22 | ( 5%) | 8 | ( 2%) |
| Missing | 0 | | 0 | | 0 | | 0 | | 2 | | 0 | | 1 | | 1 | |
| **Smoke tobacco inside, #/wk** | | | | | | | | | | | | | | | | |
| Mean (SD) | 1.8 | (2.6) | 1.2 | (2.2) | 11.8 | (15.1) | 13.3 | (18.4) | 3.0 | (3.6) | 0.8 | (1.0) | 3.1 | (3.9) | 2.8 | (2.9) |
| [Range] | | [0-8] | | [0-7] | | [0-70] | | [0-140] | | [0-7] | | [0-2] | | [0-14] | | [0-7] |
| N | | 21 | | 20 | | 134 | | 119 | | 3 | | 4 | | 22 | | 8 |
| Missing | | 1 | | 2 | | 0 | | 0 | | 0 | | 0 | | 0 | | 0 |
| **Smoke tobacco outside, #/wk** | | | | | | | | | | | | | | | | |
| Mean (SD) | 4.0 | (2.5) | 3.2 | (2.6) | 26.8 | (26.2) | 25.8 | (31.3) | 0.7 | (0.6) | 0.8 | (0.5) | 10.4 | (10.5) | 6.8 | (2.9) |
| [Range] | | [0-8] | | [0-7] | | [0-154] | | [0-168] | | [0-1] | | [0-1] | | [1-35] | | [1-10] |
| N | | 22 | | 22 | | 134 | | 118 | | 3 | | 4 | | 22 | | 8 |
| Missing | | 0 | | 0 | | 0 | | 1 | | 0 | | 0 | | 0 | | 0 |
| **Burn incense in HH (India, Guatemala)** | | | | | | | | | | | | | | | | |
| No | 387 | ( 97%) | 385 | ( 96%) | 40 | ( 10%) | 42 | ( 11%) | N/A | N/A | N/A | N/A | N/A | N/A | N/A | N/A |
| Yes | 13 | ( 3%) | 14 | ( 4%) | 359 | ( 90%) | 358 | ( 90%) | N/A | N/A | N/A | N/A | N/A | N/A | N/A | N/A |
| Missing | 0 | | 1 | | 0 | | 0 | | N/A | N/A | N/A | N/A | N/A | N/A | N/A | N/A |

| | Guatemala | | | | India | | | | Peru | | | | Rwanda | | | |
|---|---|---|---|---|---|---|---|---|---|---|---|---|---|---|---|---|
| **Variable** | **Control (N=400)** | | **Intervention (N=400)** | | **Control (N=399)** | | **Intervention (N=400)** | | **Control (N=402)** | | **Intervention (N=396)** | | **Control (N=404)** | | **Intervention (N=394)** | |
| **Baseline Exposure** | | | | | | | | | | | | | | | | |
| **Burn incense in HH times/week** | | | | | | | | | | | | | | | | |
| None | 3 | ( 23%) | 2 | ( 15%) | 0 | ( 0%) | 0 | ( 0%) | N/A | N/A | N/A | N/A | N/A | N/A | N/A | N/A |
| One | 6 | ( 46%) | 9 | ( 69%) | 93 | ( 26%) | 100 | ( 28%) | N/A | N/A | N/A | N/A | N/A | N/A | N/A | N/A |
| Two or more | 4 | ( 31%) | 2 | ( 15%) | 266 | ( 74%) | 258 | ( 72%) | N/A | N/A | N/A | N/A | N/A | N/A | N/A | N/A |
| Missing | 0 | | 1 | | 0 | | 0 | | N/A | N/A | N/A | N/A | N/A | N/A | N/A | N/A |

[a]Kitchen with a dimension greater than 25 meters or less than 0.5 meter was considered unreasonable and a data entry error
[b]Lit hours were set to missing if multiple types of stove were reported
[c]Multiple usage of stove may be reported for the same household, so households may appear more than once
[d]Multiple usage of cookstove may be reported for the same household, so households may appear more than once
[e]Multiple stoves lit may be reported for the same household, so households may appear more than once
[f]Multiple sources of smoke may be reported for the same household, so households may appear more than once. Some sources of smoke were derived from 'Other Specified'
[g]All other smoke sources to participants were derived from 'Other Specified'
[h]Calculated as multiplying [cook times/day] with [cook days/week]
[i]Multiple persons may be reported for the same household, so households may appear more than once
[j]Multiple usage of this stove may be reported for the same household, so households may appear more than once
[k]Local currency was converted to USD based on currency exchange rates: 1 Guatemalan Quetzal=0.13 USD; 1 Rwanda Franc = 0.0010 USD; 1 Peru Sol = 0.26 USD; 1 Indian Rupee = 0.014 USD
[l]Multiple ways of disposing garbage may be reported for the same household, so households may appear more than once

### Additional Methods and QA/QC: PM2.5 Sampling

Gravimetric samples were evaluated as described in the main text. Filters were excluded if damaged and/or if pressure, flow, or duration issues were detected when examining sampler logs. Issues were not necessarily exclusive (a sample could have flow, pressure, and duration issues, for example). A summary of valid and invalid samples is in Table S1.

**Table S2** Gravimetric sample validity

| IRC | Valid Samples | % Valid Neph | % Invalid | % Outliers | % Flow Issues | % Pressure Issues | % Duration Issues | % Filter Damaged |
|---|---|---|---|---|---|---|---|---|
| **Guatemala** | **2428** | **1.7** | **7.6** | **0.2** | **1.9** | **1.7** | **6.3** | **1.3** |
| **India** | **2161** | **1.5** | **14.2** | **0.3** | **1.5** | **2.4** | **10.9** | **0.9** |
| **Peru** | **2007** | **2.6** | **16.2** | **0.8** | **2.8** | **3.0** | **13** | **1.6** |
| **Rwanda** | **2084** | **6.4** | **9.9** | **1.1** | **0.4** | **0.5** | **6** | **3.4** |

**ECM gravimetric data analysis**

- ECM data files are analyzed using an automated script to assess key performance parameters, including flow rates, inlet pressure, temperature and humidity ranges, sampling duration, and other related parameters.
- $PM_{2.5}$ concentration was estimated as *[(final_filter_mass – initial_filter_mass) – blank_correction] / sample_volume [m^3]* , where sample_volume = average of pre- and post-flow rates (liters per minute) * minutes operating * [1 m^3/1000 liters].
    - sample_volume: The average of the pre- and post-sample flow calibrations are used to calculate the total sample flow when possible. Where pre- and/or post-sample flow rates are not valid, we utilized the ECM's internal flow meter.
    - blank_correction is the median mass deposition on blank filters taken to the field to assess contamination. These are combined on an annual basis per-IRC to correspond to the filters that are used in that year. 392 blanks were collected (87 in Guatemala, 70 in Rwanda, 154 in Peru, and 81 in India).
- Limit of Detection (LOD) Correction Calculations: If there are filters that fall below the LOD (calculated as three times the standard deviation of the gravimetric mass weight of the collected field blanks (3*Std Dev of Field Blanks)), we assigned an LOD-corrected mass of LOD/(Sqrt(2)). This LOD-corrected mass weight was used for estimating time-weighted average concentrations / exposures.
- Nephelometric data is not used in this phase of the study, as the gravimetric 24-hr average data is the key exposure metric of interest.

**ECM Duplicates**

253 duplicate ECM samples were collected across IRCs. Due to equipment and staffing issues, only 2 duplicates were collected in India (subsequent data collection rounds, not analyzed as part of this manuscript, include sufficient duplicate data from India). The relationship between duplicate measures is depicted in Figure S1 and Table S2.

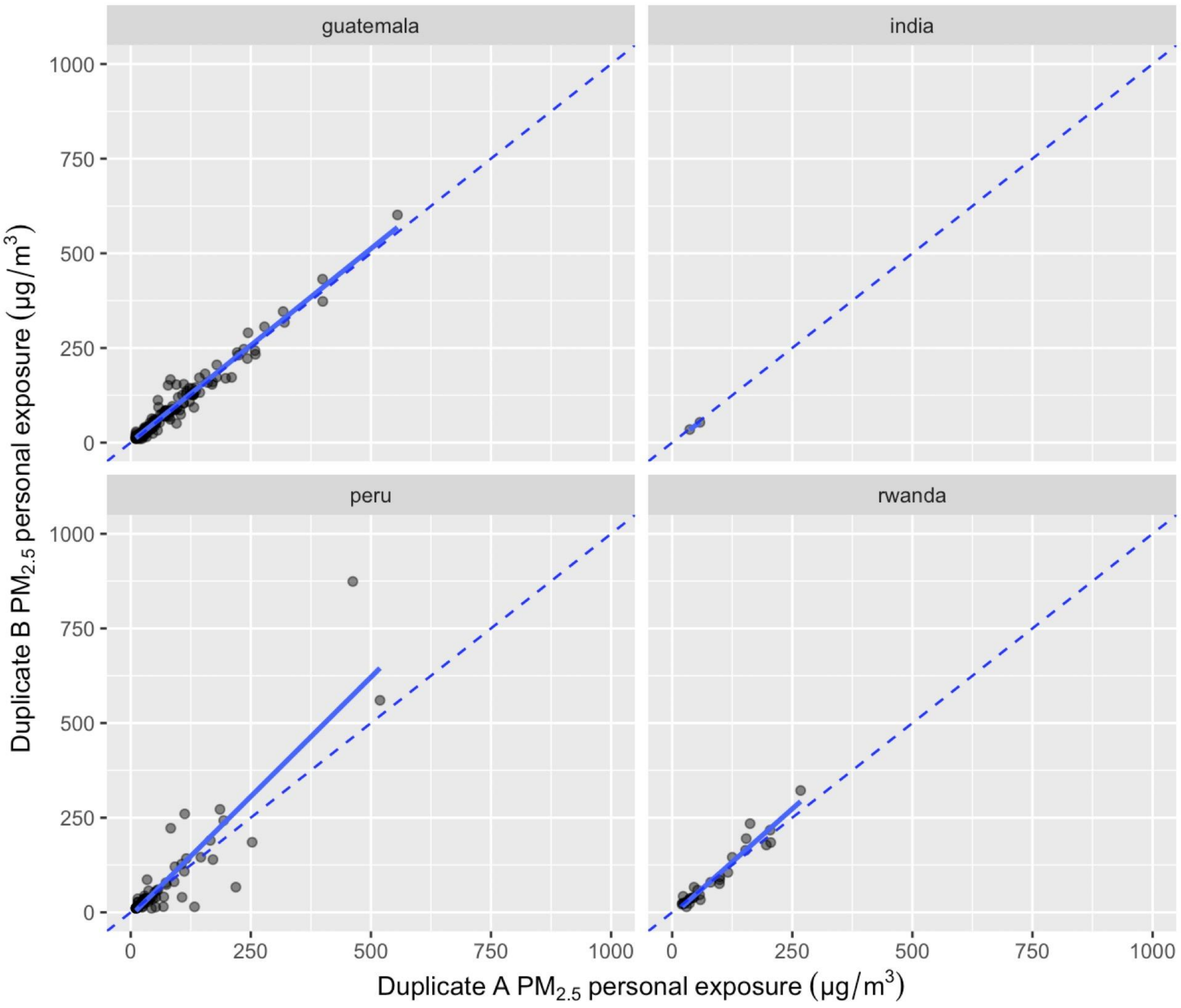

**Figure S1 ECM duplicate measures.** Panels are individual IRCs. Points are paired measurements using collocated ECMs. The dashed line is 1:1. The blue line is a linear model fit to the data points.

**Table S3** Gravimetric ECM duplicate performance and fit metrics

| IRC | $R^2$ | Slope | RMSE | N |
|---|---|---|---|---|
| Guatemala | 0.96 | 0.94 | 18 | 131 |
| India | 1 | 1.12 | 0 | 2 |
| Peru | 0.81 | 0.64 | 36 | 91 |

| | | | | |
|---|---|---|---|---|
| Rwanda | 0.93 | 0.83 | 17 | 29 |

**Processing and use of nephelometric data when gravimetric data is absent**

To generate nephelometer-based PM2.5 estimates, we developed models to calibrate light-scattering response with gravimetric data. First, we adjusted realtime baselines by setting the 1st percentile of data to 10μg/m$^3$, representative of a relatively clean ambient concentration (similar to Zhang et al[17]). Minute-averaged data were filtered for validity by excluding samples with greater than 10% of values above the saturation limit of 9000 μg/m$^3$ (9 exclusions), or if more than 10% of values were smaller than 0 μg/m$^3$ (50 exclusions), indicative of a malfunctioning nephelometer. We performed linear regressions for each instrument, with $\log_{10}$ baseline-adjusted nephelometer values as predictors of $\log_{10}$ gravimetric averages.

### Compliance

To calculate wearing compliance using accelerometer data from the ECM, a 20-minute rolling average of the 3-axis vector sum composite accelerometer data was estimated, and a threshold of 0.02 g was applied. If the value was higher than the threshold, the participant was considered as wearing the monitor for the given minute. Daytime compliance was calculated by restricting the sample to hours between 5 am and 9 pm for any given monitoring session / period. Figure S2 and Table S3 summarize compliance findings.

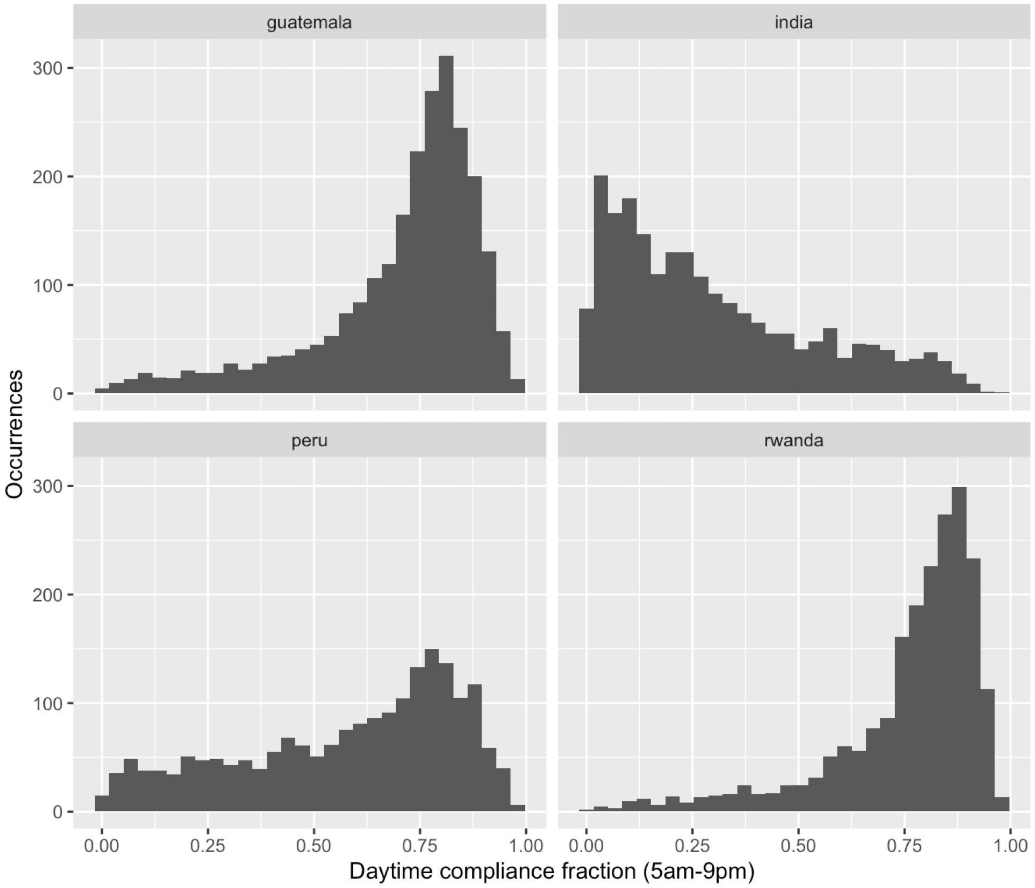

**Figure S2 Monitoring wearing compliance.** Panels are individual IRCs. Bars are the number of measurements shown as wearing compliant for a given fraction of the day. .

**Table S4** Wearing Compliance

| IRC | N | Mean | Median | SD | Min | Max |
|---|---|---|---|---|---|---|
| Guatemala | 2081 | 0.7 | 0.76 | 0.19 | 0 | 0.98 |
| India | 1919 | 0.31 | 0.25 | 0.24 | 0 | 0.98 |
| Peru | 1726 | 0.57 | 0.64 | 0.26 | 0 | 0.98 |

| | | | | | | |
|---|---|---|---|---|---|---|
| Rwanda | 1985 | 0.76 | 0.81 | 0.18 | 0 | 0.98 |

### Additional Methods and QA/QC: Black Carbon

Black carbon exposures were estimated following the methods in the main text. Data quality for BC was assessed using the same steps as for the gravimetric analysis, in addition to (1) an outlier identification step of attenuation values outside of threshold ranges (0 to 100 µg) and (2) for any sample with a gravimetric flag, the corresponding BC data were also flagged. Figure S3 and Table S4 describe relationships between duplicate BC measures (n = 234).

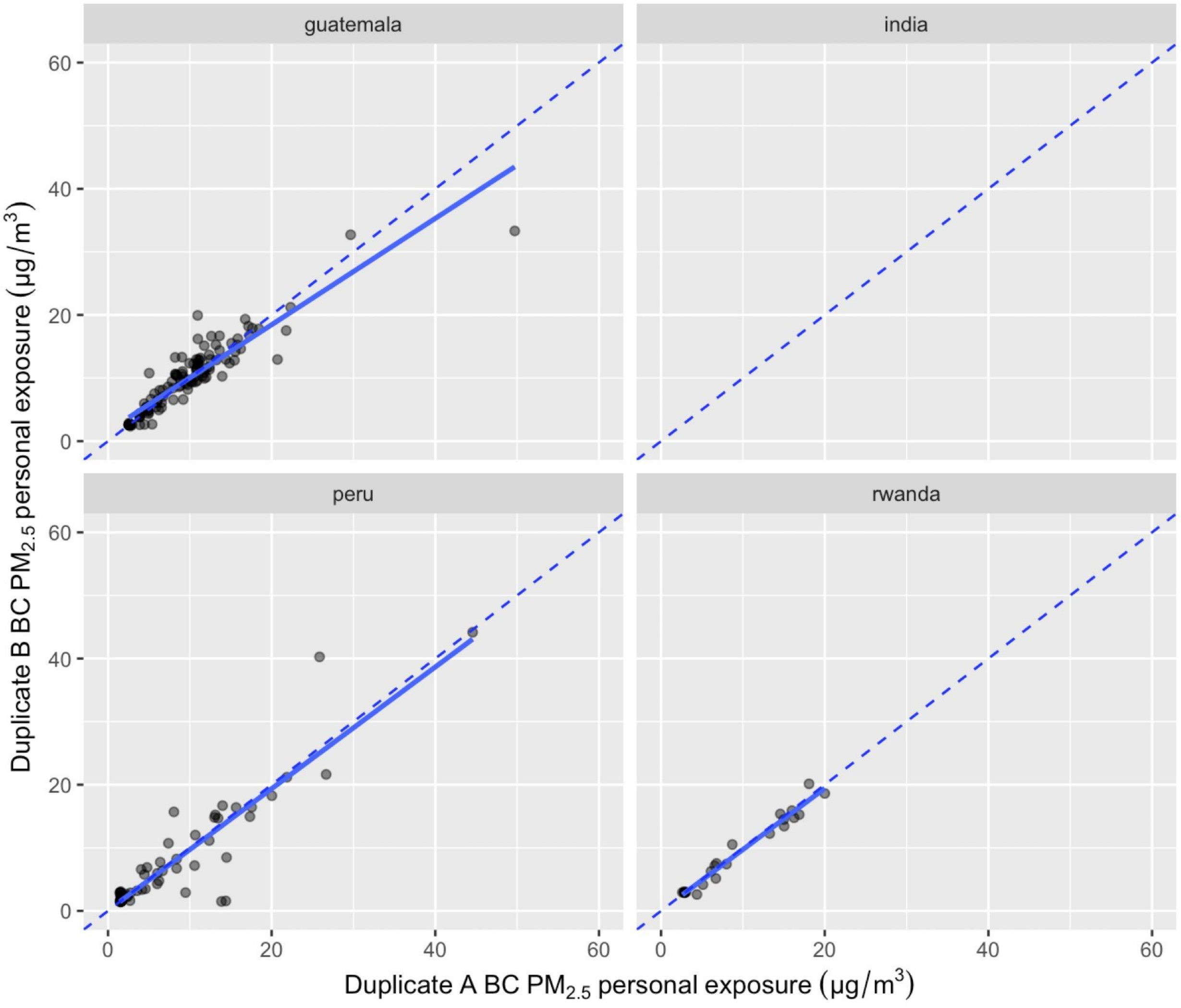

**Figure S3 BC duplicate measures.** Panels are individual IRCs. Points are paired measurements using collocated ECMs with filters analyzed via transmissometry for BC concentrations/exposures. The dashed line is 1:1. The blue line is a linear model fit to the data points.

**Table S5** BC duplicate performance and fit metrics

| IRC | $R^2$ | Slope | RMSE | N |
|---|---|---|---|---|
| Guatemala | 0.85 | 1.01 | 2.4 | 126 |
| Peru | 0.85 | 0.78 | 4.4 | 84 |
| Rwanda | 0.99 | 0.97 | 1.0 | 24 |

### Additional Methods and QA/QC: CO

Using the real-time traces from the Lascar, all CO files were visually inspected for potential artifacts or unrealistic CO exposures due to potentially contaminated or defective monitors. The files were rated as “Major Artifact”, “Minor Artifact”, “Unsure”, “Valid”, and “0ppm”. “Unsure” traces received a second blinded review and were categorized into the other three main groups. “0ppm” plots showed 0 ppm throughout the entire duration of the deployment and were removed representing around 6% of the total CO files. Ultimately, it was decided to remove only the “Major Artifacts” representing around 3% of the total CO files. Exclusions are summarized in Table S6.

**Table S6** CO sample validity

| IRC | % Invalid Samples | % Duration Issues | % Visual Flags | % Constant 0 ppm | % Values above 100 ppm |
|---|---|---|---|---|---|
| Guatemala | 3.9 | 3.2 | 1.8 | 2 | 0.1 |
| India | 5.1 | 1.7 | 3.9 | 3 | 0.1 |
| Peru | 8.8 | 4.4 | 5.7 | 9 | 0.05 |
| Rwanda | 4.9 | 3.1 | 2.4 | 5 | 0 |

422 valid, calibrated duplicate pairs of data among Lascar samples were collected. Table S7 and Figure S6 summarize the relationships between these measures.

**Table S7** CO duplicate performance and fit metrics

| IRC | $R^2$ | Slope | RMSE | N |
|---|---|---|---|---|
| Guatemala | 0.43 | 0.77 | 1.38 | 113 |
| India | 0.98 | 0.46 | 0.04 | 4 |
| Peru | 0.67 | 0.81 | 2.44 | 118 |
| Rwanda | 0.36 | 0.35 | 2.24 | 187 |

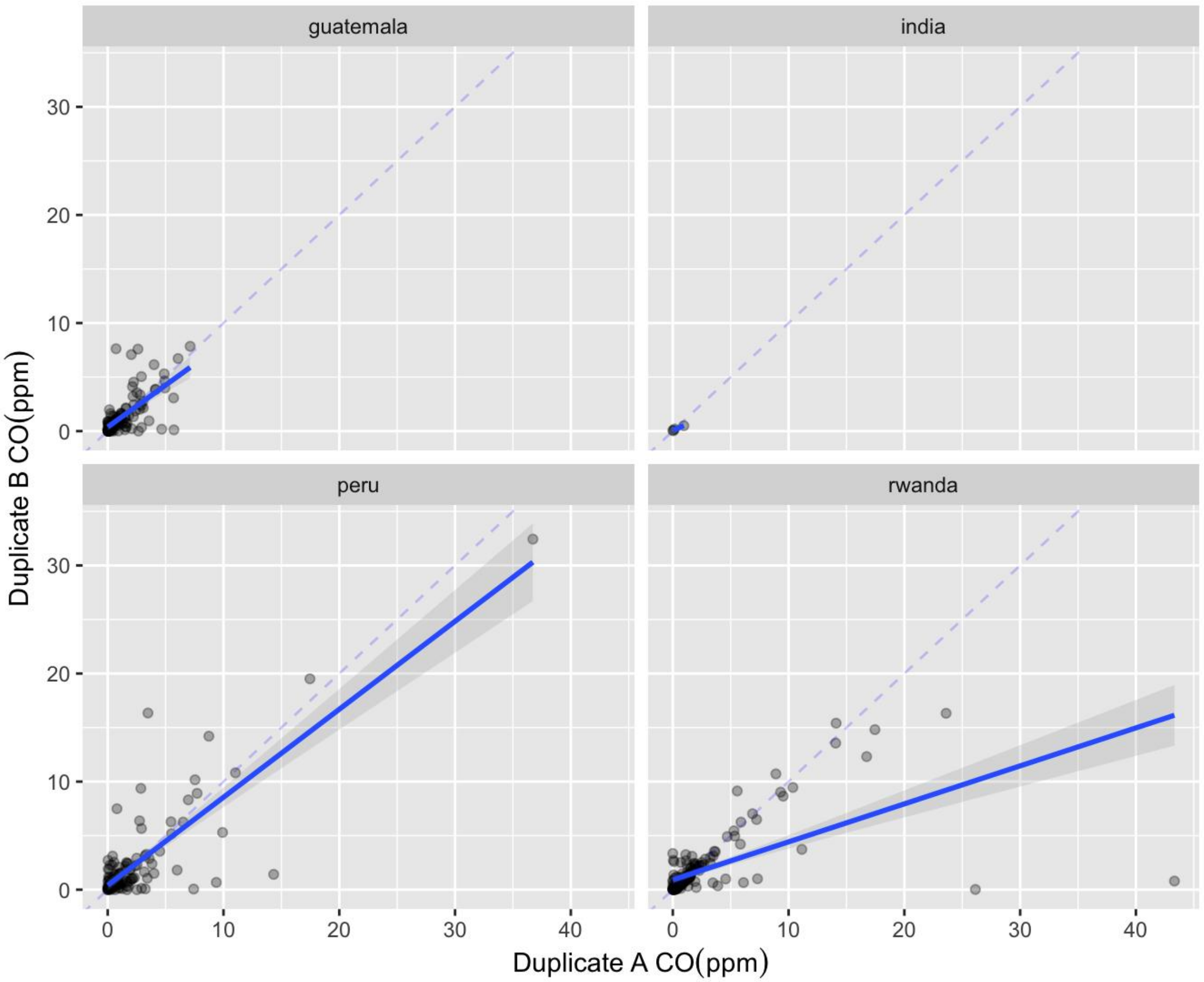

**Figure S4  CO duplicate measures.** Panels are individual IRCs. Points are paired measurements using collocated ECMs with filters analyzed via transmissometry for BC concentrations/exposures. The dashed line is 1:1. The blue line is a linear model fit to the data points. Axes are truncated.

### IRC and pollutant-specific findings

*IRC-specific control households*. In India and Guatemala, changes in PM2.5 exposures in the control group were not significant. For example, between baseline and post-intervention round 1 in Guatemala, PM levels decreased by 5% (on average 7 µg/m3); between baseline and post-intervention round 2, the mean PM exposure decreased by approximately 11 % (16 µg/m3). In India, between baseline and post-intervention round 1, the mean PM2.5 exposure decreased by 1 µg/m3 and increased by 5.6 µg/m3 on average when comparing baseline to post-intervention round 2. In Rwanda and Peru, contrastingly, control participant PM2.5 exposures decreased significantly between baseline and post-intervention measurement periods. In Rwanda, on average, control household PM levels decreased by 7 µg/m3 (6%) between baseline and post-intervention round 1 and by 16 $\mu g/m^3$ (14%) when comparing baseline and post-intervention round 2. In Peru, levels decreased by 16 ug/m3 (20%) between baseline and post-intervention round 1 and by 14 µg/m3 (17%) between baseline and post-intervention round 2.

*IRC-specific intervention households.* All sites had significant reductions between baseline and post-intervention visit 1 ($p < 0.0001$) and baseline and post-intervention visit 2 ($p < 0.0001$) for all pollutants. The difference between post-intervention rounds 1 and 2 was not significant for any pollutant at any individual study site.

*IRC-specific CO findings*. There was no significant difference between baseline and post-intervention visit 1 or post-intervention visit 2 exposures to CO among control households in Guatemala, India, and Rwanda. In Peru, there was a significant decrease in CO exposures between baseline and post-intervention round 1 ($p < 0.01$) and baseline and post-intervention round 2 ($p < 0.05$), though post-intervention rounds 1 and 2 were not significantly different.

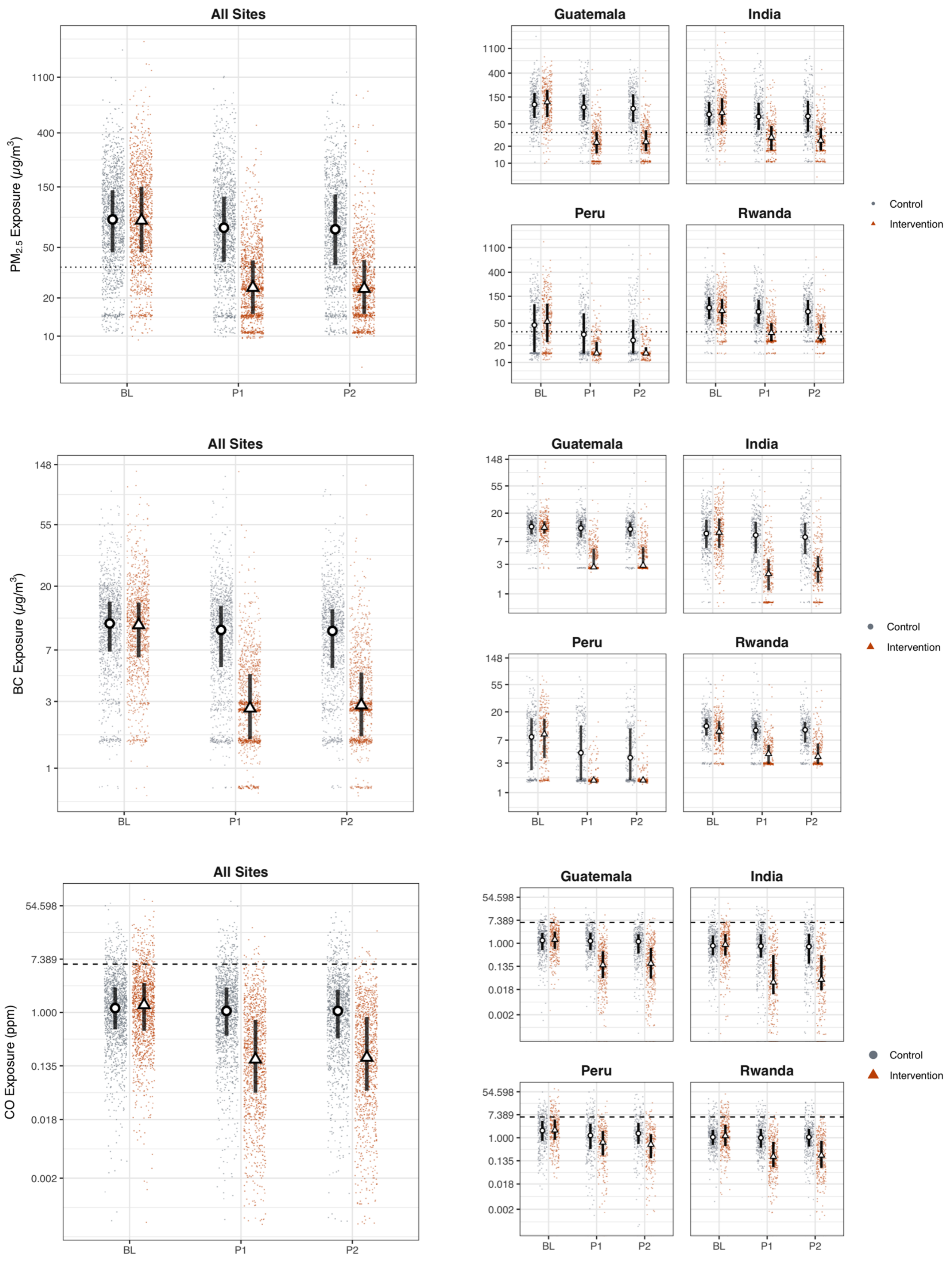

**Figure S5 HAPIN $PM_{2.5}$, Black Carbon (BC), and Carbon Monoxide (CO) exposures overall and by IRC.** Red triangles and blue dots are per-country and study round samples in intervention and control households, respectively. Circles and triangles outlined in black are median values in control and intervention households, respectively. Lines are interquartile ranges. BL = baseline (9-20 weeks gestation), P1 = post-intervention visit 1 (24-28 weeks gestation), and P2 = post-intervention visit 2 (32-36 weeks gestation). The dotted line in the PM panels is the annual WHO Interim Target 1 guideline value (35 μg/$m^3$); the dashed line in the CO plots is the WHO guideline value of 6.11 ppm (7 mg/$m^3$).

### Relationships between pollutants

Figure S5A shows a stronger relationship between PM and CO for households using a traditional biomass fuel than for households using LPG (Spearman ρ: 0.47 and 0.06, respectively). Panel B shows fairly clear and consistent correlations between PM and CO for sample groups using biomass, though the relationship varies by IRC. For traditional stoves, the Spearman ρ is 0.70, 0.62, 0.54, and 0.25 for Guatemala, India, Peru, and Rwanda, respectively. The overall, HAPIN-wide Spearman ρ was 0.51. Among post-intervention exposure samples of LPG users, relationships were much weaker (Spearman ρ 0.05-0.16), which was expected given the lack of a dominant biomass smoke source in the homes.

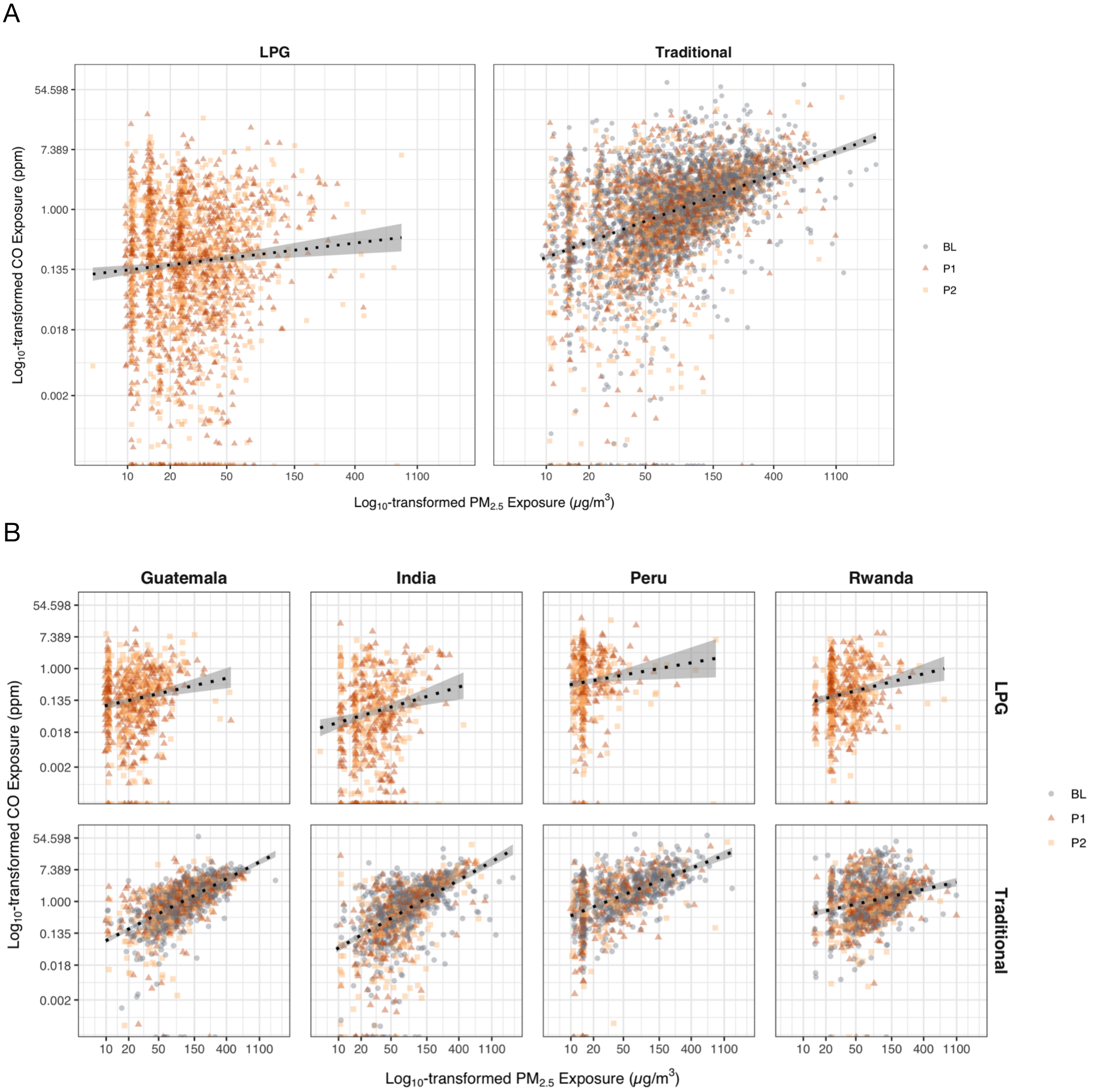

**Figure S6: HAPIN-wide and by country relationships between PM2.5 and CO by primary fuel for cooking.** Both axes are Log10 transformed. The solid lines are a linear model; the shaded areas are standard errors. "Traditional" panels include measurements made during baseline and during baseline and post-intervention 1 and 2 in control homes. "LPG" panels include measurements made post-intervention.

The relationship between PM and BC was stronger. There was some heterogeneity between countries (for biomass users, between 0.68 and 0.87; for LPG users, between 0.46 and 0.73). As BC is a constituent of PM2.5, the stronger relationships with PM2.5 compared to CO is not surprising.  Panel A of Figure S6 shows the relationship for samples from households using LPG or biomass across all of HAPIN, while Panel B shows relationships by IRC.

A

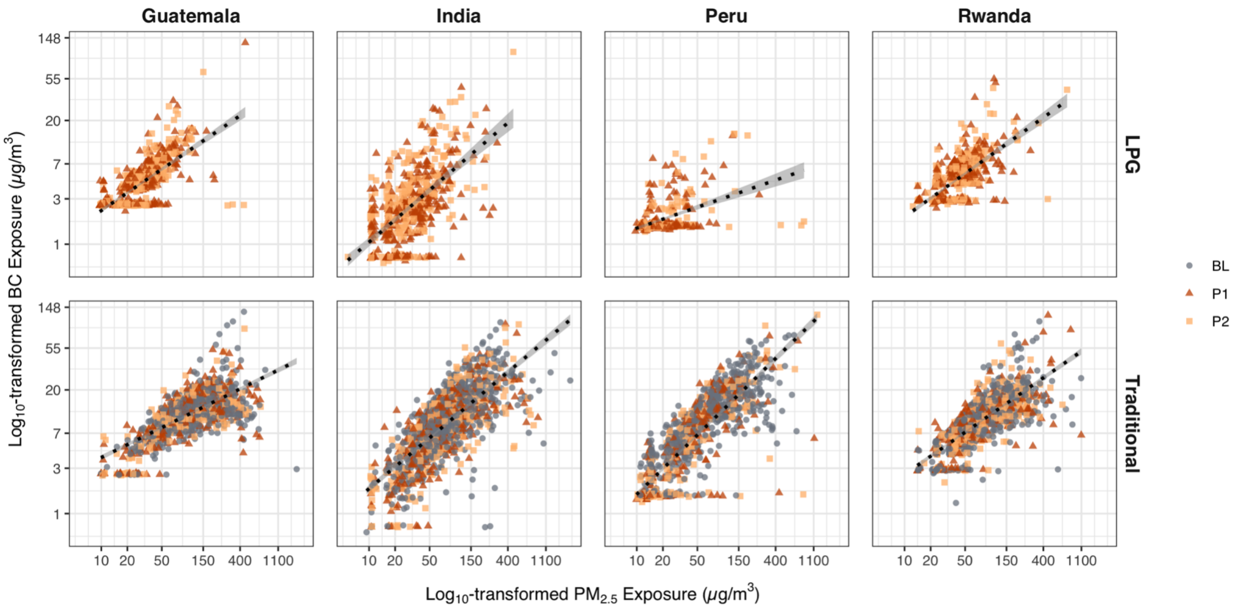

B

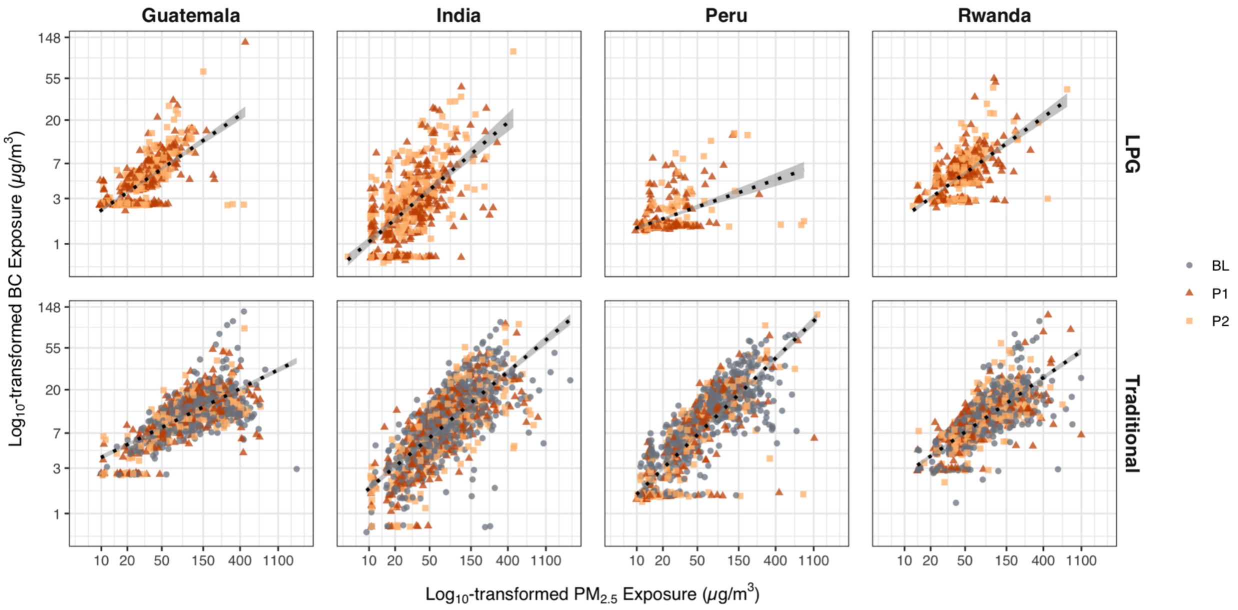

**Figure S7: HAPIN-wide and by country relationships between PM2.5 and black carbon by primary fuel for cooking.** Both axes are log10 transformed. The solid lines are a linear model; the shaded areas are standard errors. “Traditional” panels include measurements made during baseline and during baseline, post-intervention 1 and 2 in control homes. “LPG” panels include measurements made post-intervention.

**Table S8** Modeling approaches

| # | | Equation | Data | Model estimates... |
|---|---|---|---|---|
| 1 | Between groups | $log(y_{ij}) = \beta_{0ij} + \beta_{1}StudyArm_{j} + \beta_{2-10}StudySite + b_{i} + \varepsilon_{ij}$<br>$y_{ij}$is the pollutant exposure (PM2.5, CO, or BC) for participant $i$ in arm $j$. $StudyArm_{j}$ is the participant's assigned arm (reference: control), $b_{i}$is the random effect for participant $i$, $\varepsilon_{ij}$is the error term for participant $i$ in study arm $j$. $StudySite$ is the randomization strata. | Post-intervention | difference in mean exposure in intervention vs control households ($\beta_{1}$) |
| 2 | Before and after | $log(y_{ik}) = \beta_{0ik} + \beta_{1}Period_{k} + \beta_{2-10}StudySite + b_{i} + \varepsilon_{ik}$<br>$y_{ik}$is the pollutant exposure (PM2.5, CO, or BC) for participant $i$ in study period $k$. $Period_{k}$ is the measurement period (reference: baseline), $b_{i}$is the random effect for participant $i$, $\varepsilon_{ik}$is the error term for participant $i$ in study period $k$. $StudySite$ is the randomization strata. | Separately by arm | difference in mean post-intervention exposure as compared to baseline ($\beta_{1}$) |
| 3 | Comparison of Changes by period | $log(y_{ijk}) = \beta_{0ijk} + \beta_{1}StudyArm_{j} + \beta_{2}Period_{k} + \beta_{3}(StudyArm_{j} \times Period_{k}) + \beta_{4-12}StudySite + b_{i} + \varepsilon_{ijk}$<br>$y_{ijk}$is the pollutant exposure (PM2.5, CO, or BC) for participant $i$ in arm $j$and study period $k$. $StudyArm_{j}$ is the participant's assigned arm (reference: control). $Period_{k}$ is the measurement period (reference: baseline).$StudyArm_{j} \times Period_{k}$ are dummy variables for the interaction of $StudyArm_{j}$and $Period_{k}$(reference control, pre-intervention). $b_{i}$is the random effect for participant $i$. $\varepsilon_{ijk}$is the error term for participant $i$, in arm $j$, in period $k$. $StudySite$ is the randomization strata. | All data | difference in mean post-intervention exposure from the baseline period in intervention households versus the same difference in control households ($\beta_{3}$) |
| 4 | Differences-in-differences by study visit | $log(y_{ijl}) = \beta_{0ijl} + \beta_{1}StudyArm_{j} + \beta_{2}Visit_{l} + \beta_{3}(StudyArm_{j} \times Visit_{l}) + \beta_{4-12}StudySite + b_{i} + \varepsilon_{ijl}$<br>$y_{ijl}$is pollutant exposure for participant $i$ in arm $j$and study visit $l$. $StudyArm_{j}$ is the assigned arm (reference: control). $Visit_{l}$ is the measurement visit.$StudyArm_{j} \times Visit_{l}$ are dummy variables for the interaction of $StudyArm_{j}$and $Visit_{l}$(reference control, visit 1). $b_{i}$is the random effect for participant $i$. $\varepsilon_{ijl}$is the error term for participant $i$, in arm $j$, in period $l$. $StudySite$ is the randomization strata. | All data | difference in post-intervention exposure from the baseline period in intervention households by study visit versus the same difference in control households ($\beta_{3}$) |

### Modeling the impact of the intervention

As described in the main text, multiple model specifications -- with different parameters of interest -- were utilized to assess the impact of the intervention on personal exposures. Table S8 above provides a summary of modeling approaches assessed. Findings for the primary models for BC and CO are presented below.

*Black carbon.*

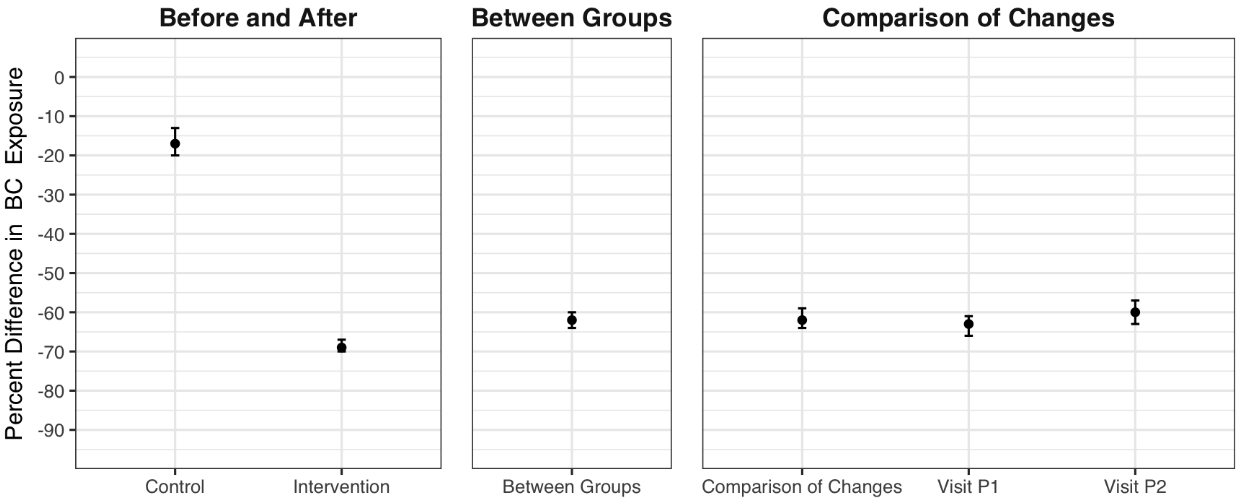

**Figure S8: Estimated impacts of the HAPIN LPG intervention on BC exposure.** All linear mixed effects models had log transformed BC exposure as the dependent variable. Whiskers are 95% confidence intervals. The first panel ("Before and After") uses data from both the control and intervention arms and compares the intervention period to the baseline period. The second panel ("Between Groups") uses only data from the intervention period and contrasts the intervention arm with the control arm. The third panel ("Comparison of Changes") uses all data from both study arms and both study periods; the model term of interest is the interaction between study arm and period, after controlling for each variable separately in the model. The "Overall" points consider an average post-intervention exposure; the Visit-specific points consider each post-randomization visit separately.

**Table S9** Percent decreases in BC exposure associated with the HAPIN LPG intervention.

| Model Type | Model Details | Estimate | Upper CI | Lower CI |
|---|---|---|---|---|
| Between Groups | Between Groups | -62 | -60 | -64 |
| Before and After | Control | -17 | -13 | -20 |
| Before and After | Intervention | -69 | -67 | -70 |
| Comparison of Changes | Overall | -62 | -59 | -64 |
| Comparison of Changes | Visit P1 | -63 | -61 | -66 |
| Comparison of Changes | Visit P2 | -60 | -57 | -63 |

*Carbon monoxide.*

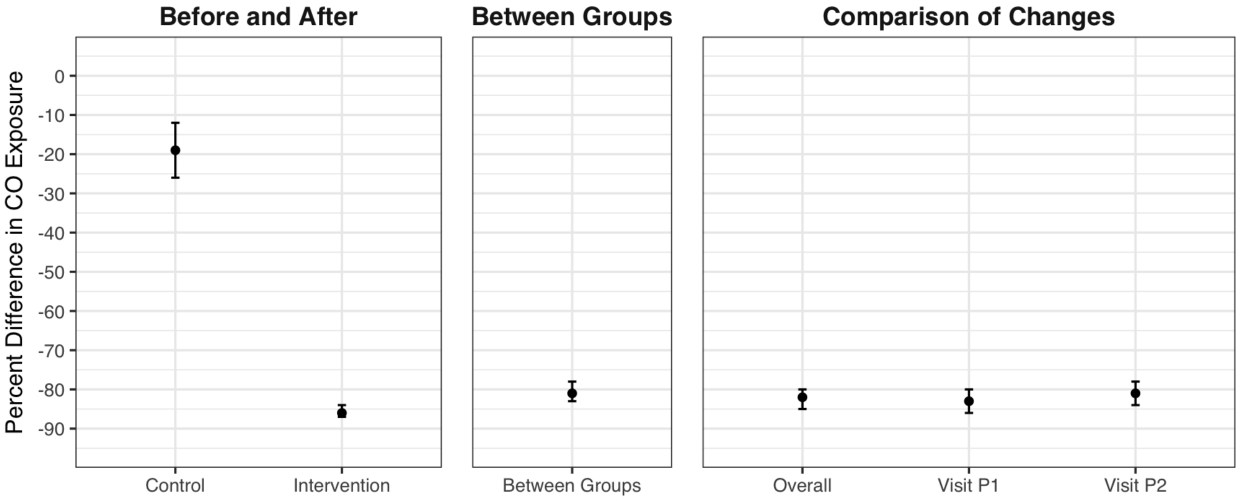

**Figure S9: Estimated impacts of the HAPIN LPG intervention on CO exposure.** All linear mixed effects models had log transformed CO exposure as the dependent variable. Whiskers are 95% confidence intervals. The first panel ("Before and After") uses data from both the control and intervention arms and compares the intervention period to the baseline period. The second panel ("Between Groups") uses only data from the intervention period and contrasts the intervention arm with the control arm. The third panel ("Comparison of Changes") uses all data from both study arms and both study periods; the model term of interest is the interaction between study arm and period, after controlling for each variable separately in the model. The "Overall" points consider an average post-intervention exposure; the Visit-specific points consider each post-randomization visit separately.

**Table S10** Percent decreases in CO exposure associated with the HAPIN LPG intervention.

| Model Type | Model Details | Estimate | Upper CI | Lower CI |
|---|---|---|---|---|
| Between Groups | Between Groups | -81 | -78 | -83 |
| Before and After | Control | -19 | -12 | -26 |
| Before and After | Intervention | -86 | -84 | -87 |
| Comparison of Changes | Overall | -82 | -80 | -85 |
| Comparison of Changes | Visit P1 | -83 | -80 | -86 |
| Comparison of Changes | Visit P2 | -81 | -78 | -84 |

### IRC-specific models of the impact of the intervention

Output of the same models as presented in the main text are presented below, run individually for each IRC (Guatemala, India, Peru, and Rwanda). Figures follow.

**Table S11: IRC-specific models of the impact of the intervention on $PM_{2.5}$**

| IRC | Model | Study Visit | Arm | Estimate | Upper CI | Lower CI |
|---|---|---|---|---|---|---|
| Guatemala | Between Groups | | | -74 | -71 | -76 |
| India | Between Groups | | | -59 | -55 | -63 |
| Peru | Between Groups | | | -50 | -44 | -55 |
| Rwanda | Between Groups | | | -54 | -51 | -58 |
| Guatemala | Before and After | | Intervention | -78 | -76 | -80 |
| Guatemala | Before and After | | Control | -12 | -6 | -18 |
| India | Before and After | | Intervention | -66 | -62 | -69 |
| India | Before and After | | Control | -9 | 0 | -17 |
| Peru | Before and After | | Intervention | -67 | -63 | -70 |
| Peru | Before and After | | Control | -28 | -19 | -37 |
| Rwanda | Before and After | | Intervention | -55 | -52 | -59 |
| Rwanda | Before and After | | Control | -13 | -6 | -20 |
| Guatemala | Comparison of Changes | | | -75 | -73 | -78 |
| India | Comparison of Changes | | | -62 | -57 | -67 |
| Peru | Comparison of Changes | | | -54 | -46 | -61 |
| Rwanda | Comparison of Changes | | | -49 | -43 | -54 |
| Guatemala | Comparison of Changes | 1 | | -77 | -74 | -80 |
| Guatemala | Comparison of Changes | 2 | | -74 | -70 | -77 |
| India | Comparison of Changes | 1 | | -60 | -54 | -65 |
| India | Comparison of Changes | 2 | | -64 | -58 | -69 |
| Peru | Comparison of Changes | 1 | | -55 | -47 | -63 |
| Peru | Comparison of Changes | 2 | | -52 | -42 | -61 |
| Rwanda | Comparison of Changes | 1 | | -49 | -42 | -55 |
| Rwanda | Comparison of Changes | 2 | | -48 | -41 | -55 |

**Figure S10 IRC-specific models of the impact of the intervention on $PM_{2.5}$**

**A. Between Groups**

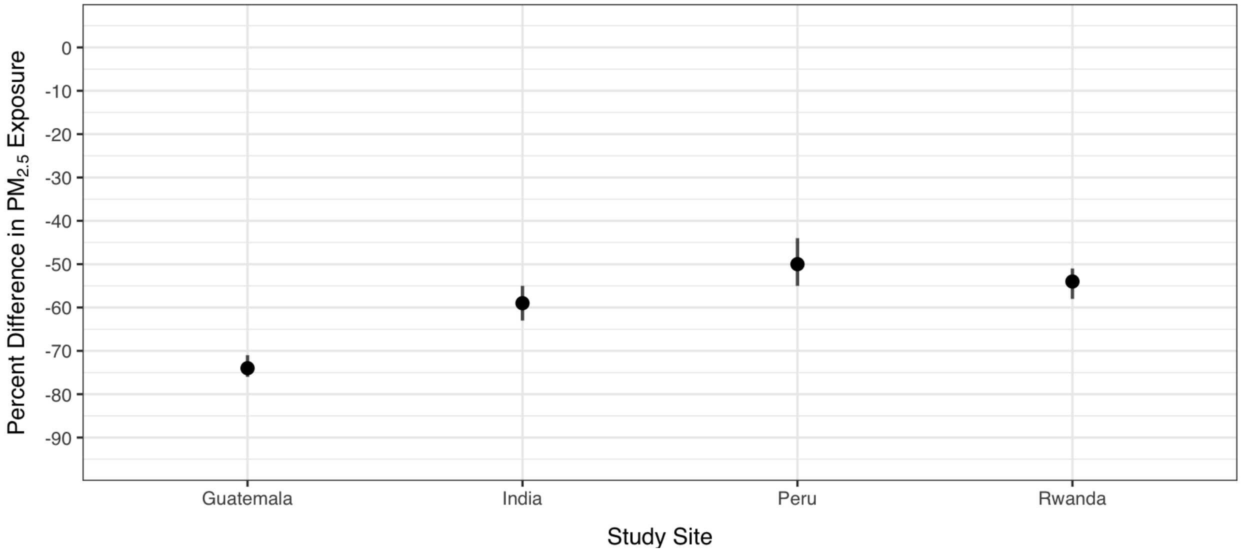

**B. Before and After**

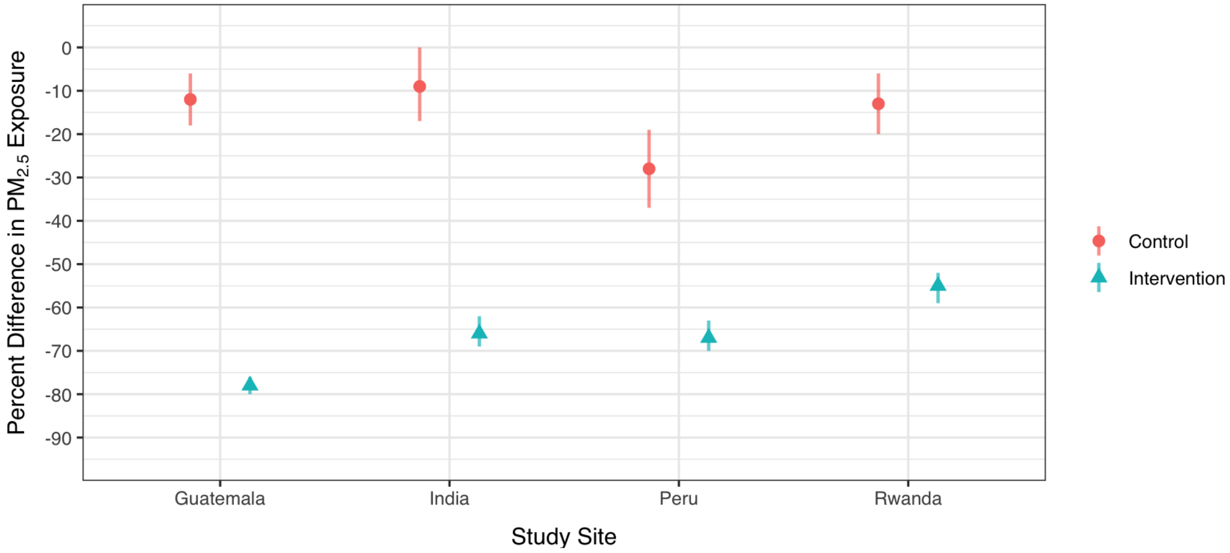

### C. Comparison of Changes

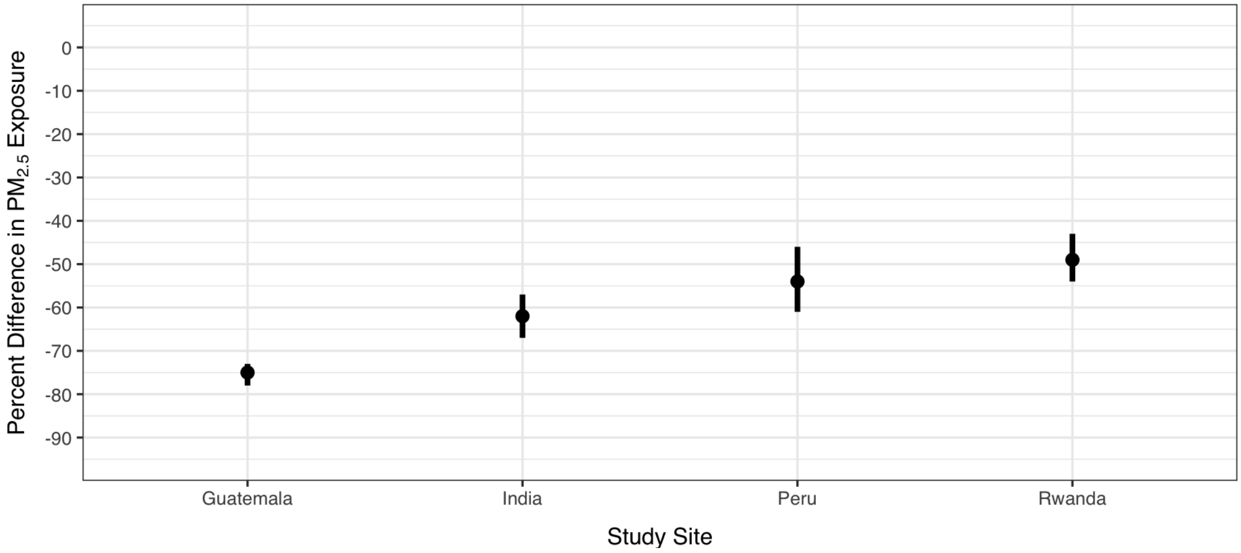

### D. Comparison of Changes by Visit

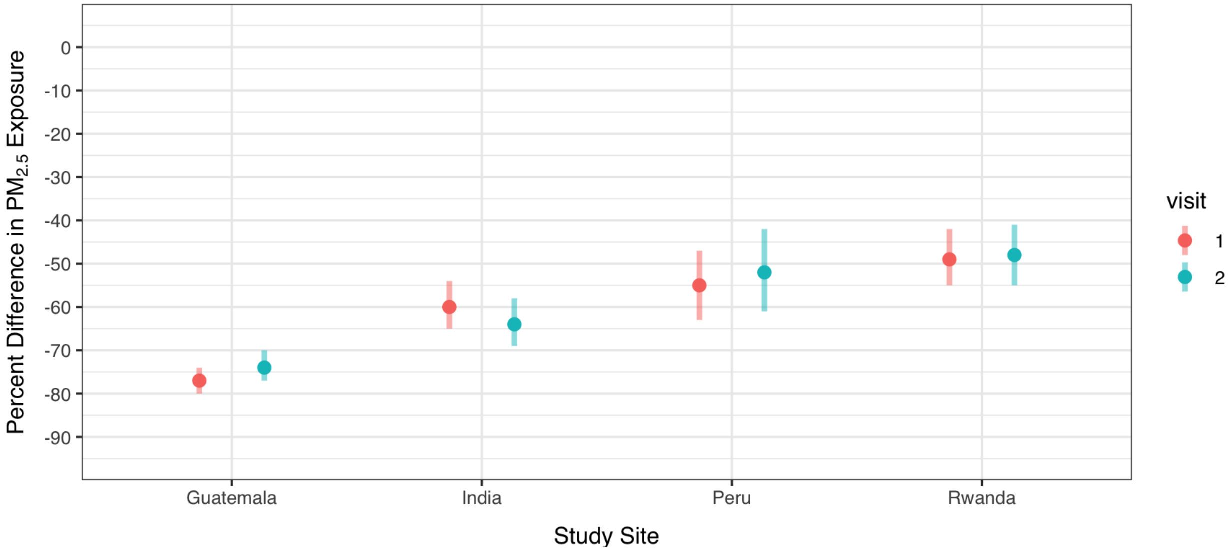

**Table S12: IRC-specific models of the impact of the intervention on Black Carbon**

| IRC | Model | Study Visit | Arm | Estimate | Upper CI | Lower CI |
|---|---|---|---|---|---|---|
| Guatemala | Between Groups | | | -62 | -60 | -65 |
| India | Between Groups | | | -69 | -66 | -73 |
| Peru | Between Groups | | | -62 | -57 | -66 |
| Rwanda | Between Groups | | | -53 | -50 | -56 |
| Guatemala | Before and After | | Intervention | -65 | -63 | -68 |
| Guatemala | Before and After | | Control | -8 | -3 | -13 |
| India | Before and After | | Intervention | -75 | -72 | -77 |
| India | Before and After | | Control | -16 | -8 | -23 |
| Peru | Before and After | | Intervention | -77 | -74 | -79 |
| Peru | Before and After | | Control | -35 | -25 | -43 |
| Rwanda | Before and After | | Intervention | -52 | -48 | -55 |
| Rwanda | Before and After | | Control | -11 | -5 | -17 |
| Guatemala | Comparison of Changes | | | -62 | -59 | -65 |
| India | Comparison of Changes | | | -70 | -66 | -74 |
| Peru | Comparison of Changes | | | -64 | -58 | -70 |
| Rwanda | Comparison of Changes | | | -45 | -40 | -51 |
| Guatemala | Comparison of Changes | 1 | | -64 | -60 | -67 |
| Guatemala | Comparison of Changes | 2 | | -61 | -57 | -65 |
| India | Comparison of Changes | 1 | | -73 | -68 | -77 |
| India | Comparison of Changes | 2 | | -67 | -62 | -72 |
| Peru | Comparison of Changes | 1 | | -66 | -59 | -72 |
| Peru | Comparison of Changes | 2 | | -63 | -55 | -69 |
| Rwanda | Comparison of Changes | 1 | | -46 | -40 | -52 |
| Rwanda | Comparison of Changes | 2 | | -45 | -38 | -51 |

**Figure S11: IRC-specific models of the impact of the intervention on Black Carbon**

**A. Between Groups**

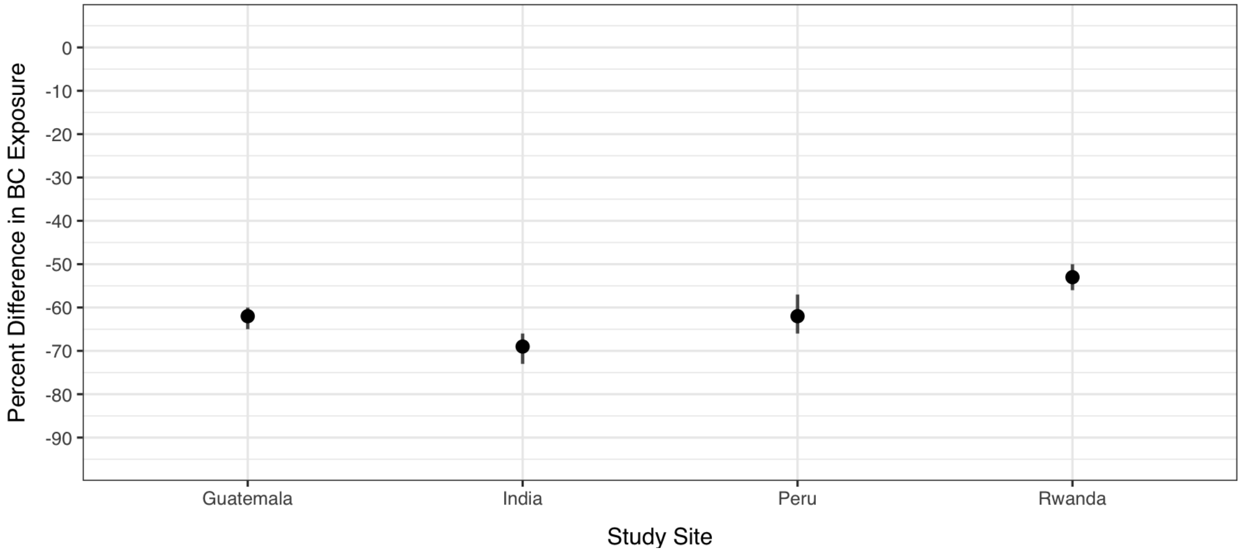

**B. Before and After**

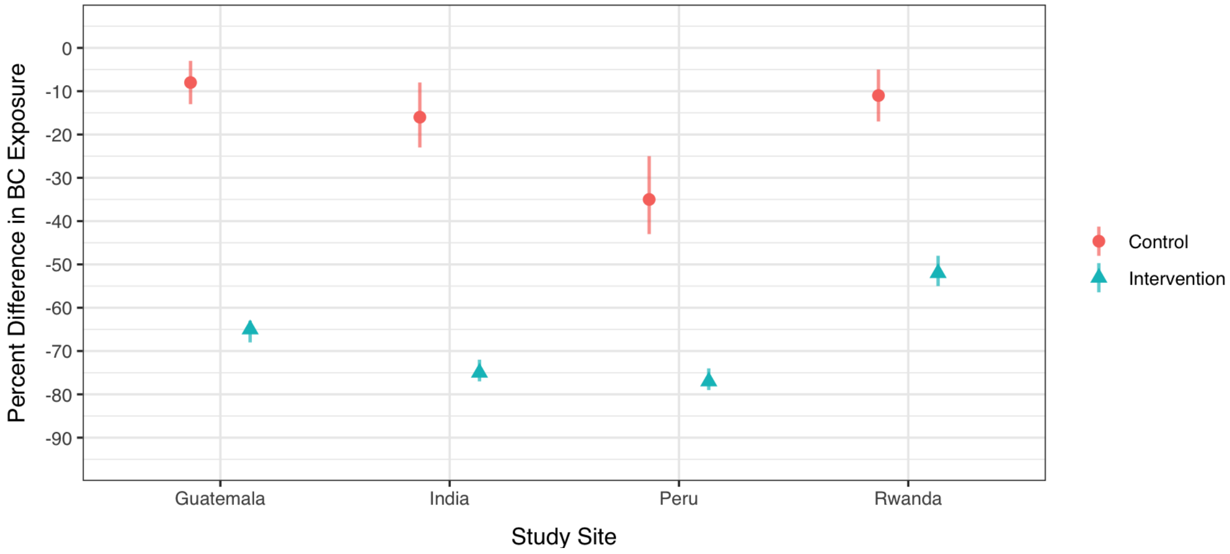

### C. Comparison of Changes

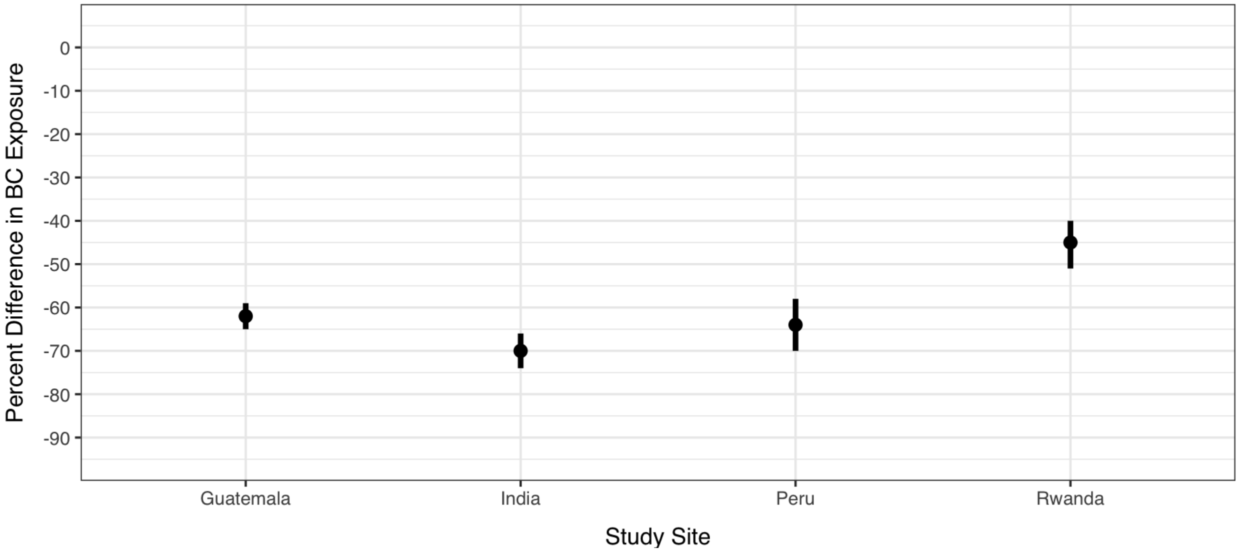

### D. Comparison of Changes by Visit

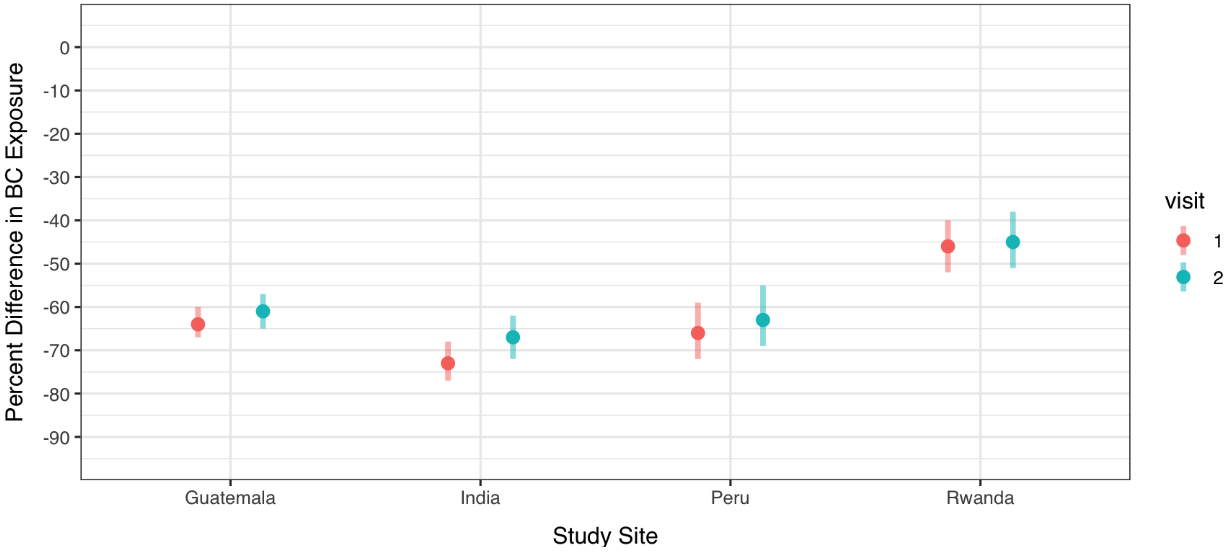

**Table S13: IRC-specific models of the impact of the intervention on Carbon Monoxide**

| IRC | Model | Arm | Study Visit | Estimate | Upper CI | Lower CI |
|---|---|---|---|---|---|---|
| Guatemala | Between Groups | | | -84 | -81 | -87 |
| India | Between Groups | | | -89 | -85 | -91 |
| Peru | Between Groups | | | -57 | -46 | -66 |
| Rwanda | Between Groups | | | -78 | -74 | -82 |
| Guatemala | Before and After | Intervention | | -88 | -85 | -90 |
| Guatemala | Before and After | Control | | -11 | 3 | -23 |
| India | Before and After | Intervention | | -91 | -88 | -93 |
| India | Before and After | Control | | -22 | -4 | -37 |
| Peru | Before and After | Intervention | | -74 | -68 | -80 |
| Peru | Before and After | Control | | -33 | -17 | -47 |
| Rwanda | Before and After | Intervention | | -83 | -80 | -86 |
| Rwanda | Before and After | Control | | -9 | 4 | -21 |
| Guatemala | Comparison of Changes | | | -86 | -82 | -89 |
| India | Comparison of Changes | | | -89 | -84 | -92 |
| Peru | Comparison of Changes | | | -62 | -48 | -72 |
| Rwanda | Comparison of Changes | | | -81 | -76 | -85 |
| Guatemala | Comparison of Changes | | 1 | -88 | -84 | -91 |
| Guatemala | Comparison of Changes | | 2 | -83 | -78 | -88 |
| India | Comparison of Changes | | 1 | -90 | -86 | -93 |
| India | Comparison of Changes | | 2 | -86 | -79 | -91 |
| Peru | Comparison of Changes | | 1 | -55 | -35 | -68 |
| Peru | Comparison of Changes | | 2 | -69 | -55 | -79 |
| Rwanda | Comparison of Changes | | 1 | -82 | -76 | -86 |
| Rwanda | Comparison of Changes | | 2 | -81 | -75 | -86 |

**Figure S12: IRC-specific models of the impact of the intervention on Carbon Monoxide**

**A. Between Groups**

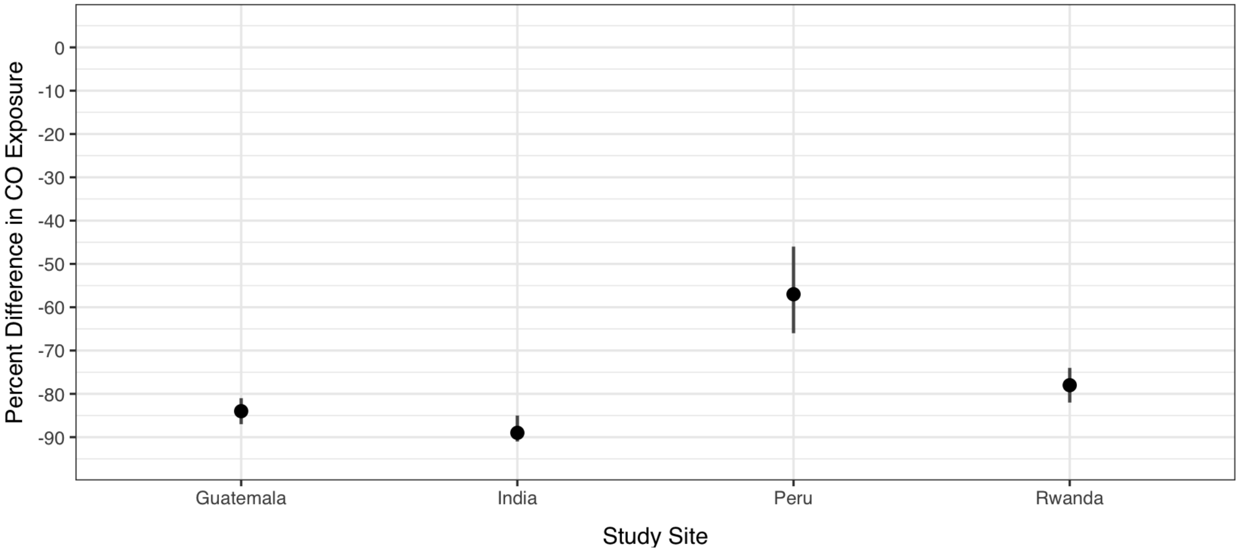

**B. Before and After**

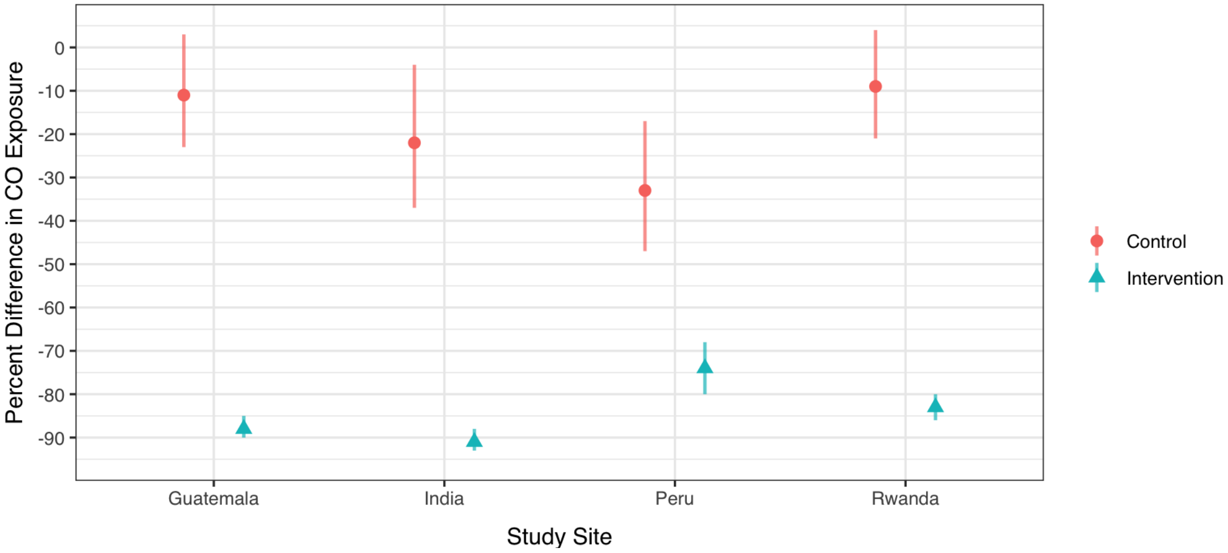

### C. Comparison of Changes

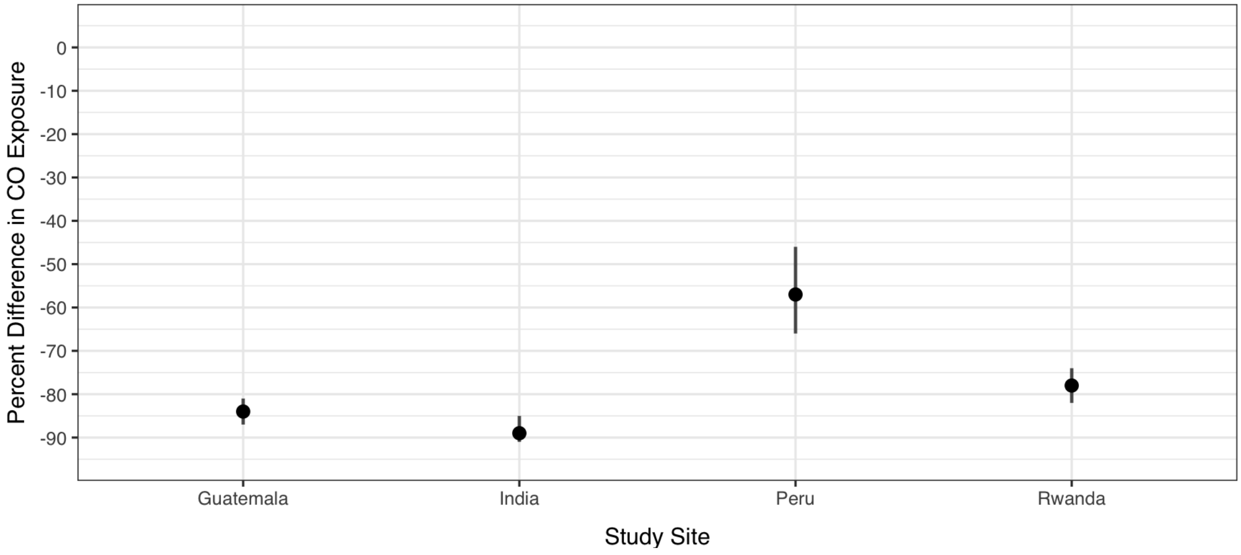

### D. Comparison of Changes by Visit

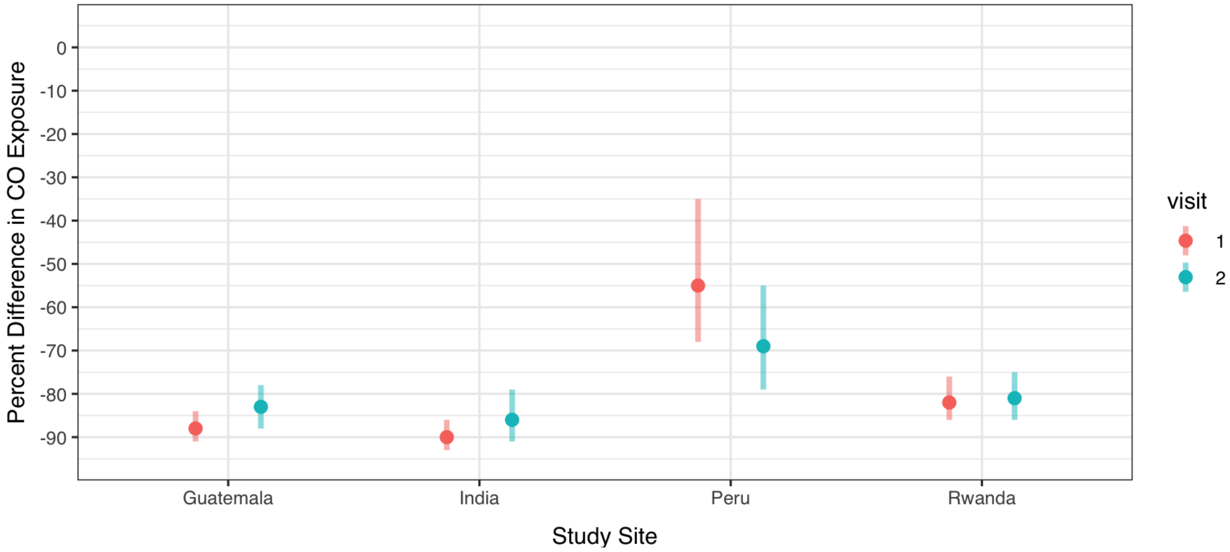
